# Single-Set Blood Culture Restriction During the 2024 National Blood Culture Bottle Shortage: An Interrupted Time Series Analysis of Patient Outcomes

**DOI:** 10.1101/2025.09.24.25335834

**Authors:** Joseph B. Ladines-Lim, Bailey Van, Leigh Cressman, Warren B. Bilker, Kyle Rodino, Laurel Glaser, Kathleen O. Degnan, Michael Z. David

## Abstract

**Importance:** The 2024 national blood culture bottle shortage led some hospitals to adopt single-set blood culture restrictions, conflicting with professional society guidance for 2–3 sets and risking underdiagnosis. Patient outcomes are not well studied.

**Objective:** To evaluate the impact of single-set blood culture restriction on patient outcomes, culture use, and antimicrobial therapy.

**Design, Setting, and Participants:** Interrupted time series analysis of 147,214 hospitalizations (36,909 with ≥1 blood culture) across 3 tertiary hospitals in an urban academic center, June 26, 2023–June 25, 2025. Periods were categorized as pre-restriction, restriction, and post-restriction. Analyses were conducted overall, among hospitalizations with ≥1 blood culture set (≥1-BC hospitalizations), and by hospital.

**Exposure:** Strict electronic health record order restriction limiting to 1 blood culture set per patient every 24 hours (June 26–December 23, 2024).

**Main Outcomes and Measures:** Primary outcomes included in-hospital mortality or hospice discharge, 30-day revisits, and length of stay (LOS). Secondary outcomes included blood culture metrics (positivity, number, timing, proportion with ≥1 culture) and receipt and days of antimicrobials. Odds or incidence rate ratios were reported.

**Results:** Among all hospitalizations, in-hospital mortality/hospice discharge declined pre-restriction (–1.3%/week, P<.001), plateaued during restriction (+0.6%/week, P=.33), and resumed decline post-restriction (–2.8%/week, P<.001). Among ≥1-BC hospitalizations, trends were similar, with additional 37.6% increase upon restriction onset (P=.005); LOS increased 14.9% upon restriction onset (P<.001) then decreased post-restriction (–0.9%/week, P<.001). 30-day revisits were unchanged. Overall culture positivity increased 37.8% upon restriction onset (P<.001) and decreased 27.1% upon restriction withdrawal (P<.001). The proportion of hospitalizations with ≥1 culture decreased 37.7% among all hospitalizations (P<.001) and mean number of cultures per hospitalization decreased 49.2% among ≥1-BC hospitalizations (P<.001) upon restriction onset, both partially rebounding afterward. Among ≥1-BC hospitalizations, time from admission to first culture collection and antimicrobial administration increased 72.2% (P<.001) and 21.5% (P=.001), respectively, upon restriction onset; antimicrobial use increased 24.9% upon restriction onset (P=.02) and decreased 14.7% upon post-restriction onset (P=.19).

**Conclusions and Relevance:** Single-set blood culture restriction was associated with decreased and delayed testing, delayed antimicrobial start, and increased in-hospital mortality/hospice discharge. Findings underscore the need for optimal diagnostic stewardship practices and supply-chain resiliency for critical diagnostic supplies.

**Key Points:** *Question:* What was the impact of restricting hospitals to 1 blood culture set per patient every 24 hours during the 2024 national shortage of blood culture bottles?

*Findings:* In this interrupted time series analysis of 147,214 hospitalizations (36,909 with ≥1 blood culture), single-set restriction was associated with decreased culture utilization, delays in time to obtaining cultures and antimicrobial administration, and increased in-hospital mortality/hospice discharge which interrupted otherwise declining trends.

*Meaning:* Single-set blood culture restrictions may impede detection of true bloodstream infections, delay antimicrobial prescribing, and worsen patient outcomes, underscoring the need for diagnostic stewardship and resilient supply chains for critical testing supplies.

## Introduction

Supply chain shocks and shortages of drugs or medical supplies have substantially affected healthcare delivery and patient outcomes, particularly during COVID-19 and recent weather-related manufacturing disruptions.^1–10^ In June 2024, a national shortage of BD BACTEC^™^ blood culture bottles manufactured by Becton Dickinson^11^ prompted conservation guidance from Centers for Disease Control and Prevention (CDC),^12^ aligning with ongoing blood culture diagnostic stewardship efforts.^13–20^ Hospitals implemented various measures, including stewardship education,^21–23^ clinical decision support tools,^23–26^ and blood culture ordering restrictions,^22,23,27–29^ with reports of decreased culture usage.^21,22,24–29^

However, the specific approach adopted by some hospitals to restrict to 1 blood culture set per patient^22,27^ conflicted with guidance from the Infectious Diseases Society of America and the American Society for Microbiology to collect 2–3 sets for suspected bloodstream infections to maximize sensitivity, adjudicate possible contaminants, and facilitate discontinuation of unnecessary antimicrobials.^30^ Some hospitals reported lower culture positivity during the shortage,^27,29^ suggesting underdiagnosis.^27^

At our institution, due to imminent supply depletion, an electronic health record (EHR) order restriction of 1 blood culture set per patient every 24 hours began June 26, 2024 and ended December 23, 2024, after sufficient blood culture bottle supplies were secured. While necessary due to the critical shortage, we hypothesized that the restriction may have impacted patient outcomes and tested this hypothesis in an interrupted time series analysis.

## Methods

### Study Setting

We studied 3 hospitals within a large, urban academic health system in Philadelphia, Pennsylvania: the Hospital of the University of Pennsylvania (HUP), Penn Presbyterian Medical Center (PPMC), and Pennsylvania Hospital (PAH), which have 1069, 347, and 462 beds, respectively.

### Data Source, Exposure, and Defined Time Periods

We extracted data from Epic Clarity for inpatient hospitalizations from June 26, 2023 to June 25, 2025. We divided these into the pre-restriction (June 26, 2023–June 25, 2024), restriction (June 26, 2024–December 23, 2024), and post-restriction (December 24, 2024–June 25, 2025) periods. During pre- and post-restriction, clinicians had no blood culture order limits, whereas during restriction, an EHR hard stop limited orders to 1 set per patient every 24 hours (<10 exceptions granted by the Clinical Microbiology Laboratory). Clinicians were notified of the restriction by email the day before, with a daily EHR reminder for approximately the first 3 months of the restriction period. Upon withdrawal on December 23, 2024, a new blood culture order panel simultaneously took effect to facilitate appropriate ordering (see **eMethods 1**).^15,31^ We assigned any encounters spanning cutoff dates to the period with the majority of encounter time.

We followed the Strengthening the Reporting of Observational Studies in Epidemiology reporting guideline.^32^ The University of Pennsylvania Institutional Review Board deemed the study exempt.

### Population and Outcomes

We aggregated hospitalizations into 1-week intervals. Baseline data included patient sociodemographic characteristics (sex, age, race, and ethnicity), Elixhauser Comorbidity Index based on International Classification of Diseases, 10^th^ Edition (ICD-10) codes at hospitalization,^33–35^ insurance coverage, and hospital.

We assessed outcomes per hospitalization, both all hospitalizations and hospitalizations with ≥1 blood culture obtained (≥1-BC hospitalizations), except overall culture positivity (total number of positive blood cultures divided by total number of cultures obtained for the entire health system or hospital). Primary outcomes included all-cause in-hospital mortality or hospice discharge; number of revisits within 30 days of discharge, including inpatient admissions, medical observation, and emergency department (ED) revisits; and length of stay (LOS).

Secondary outcomes included blood culture metrics. For all hospitalizations, we determined whether ≥1 culture (regardless of positivity) was obtained at any point during hospitalization. For ≥1-BC hospitalizations, we measured whether ≥1 positive culture was obtained; total number of cultures and positive cultures obtained; number of calendar days with ≥1 culture or positive culture obtained; and time from admission to first culture obtained. We also examined antimicrobial use outcomes (any systemic antimicrobial including broad-spectrum and non-broad-spectrum; see **eMethods 2**): whether ≥1 antimicrobial was administered at any point; number of days of antimicrobial therapy; and time from admission to first antimicrobial administered.

### Statistical Analysis

We used descriptive statistics to report baseline sociodemographic and clinical characteristics. We used interrupted time series analysis to assess slope and level changes before and after the cutoff dates of June 26 and December 23, 2024, fitting segmented logistic and Poisson regression for binary and count/continuous outcomes, respectively. We employed Newey-West robust standard errors,^36^ with a lag after each change point and maximum lag length guided by the Cumby-Huizinga test.^37^

To further quantify impact of the restriction on in-hospital mortality or hospice discharge, we modeled counterfactual projections. We extrapolated the pre-restriction slope into the restriction period, estimating the expected number of events if pre-restriction trends had continued unchanged by computing the difference between the observed model-based predictions and pre-restriction counterfactual, summed across all weeks of the restriction period. We performed the analogous computation comparing the restriction period counterfactual with the post-restriction period. We generated 95% confidence intervals using parametric bootstrap resampling of the coefficient covariance matrix.

We repeated analyses for each hospital, employed 2-sided hypothesis tests with α = 0.05, and conducted all analyses using R version 4.3.0 (R Foundation for Statistical Computing, Vienna, Austria).^38^

### Sensitivity Analyses

Although restriction implementation and withdrawal were immediate, some outcomes could have lagged. Therefore, we repeated analyses with 1- and 2-week washout periods before and after cutoff dates. Additionally, because hospitalizations spanning cutoff dates have inherent ambiguity regarding restriction exposure, we repeated analyses excluding these hospitalizations.

## Results

### Study Population

We identified 147,214 hospitalizations and 36,906 ≥1-BC hospitalizations during the study period. Baseline characteristics were similar across time periods, both overall (**Table 1**) and by hospital (**eTables 1–3**). Among all hospitalizations, in-hospital mortality or hospice discharge declined from 2.5% pre-restriction to 1.9% afterward (**eTable 4**) with similar hospital-specific trends (**eTables 5–7**). Overall culture positivity increased from 4.8% to 7.9% during restriction, decreasing to 6.8% post-restriction. The proportion of hospitalizations with ≥1 blood culture fell from 27.6% pre-restriction to 20.8% during restriction before increasing to 24.2% post-restriction.

**Table 1.**
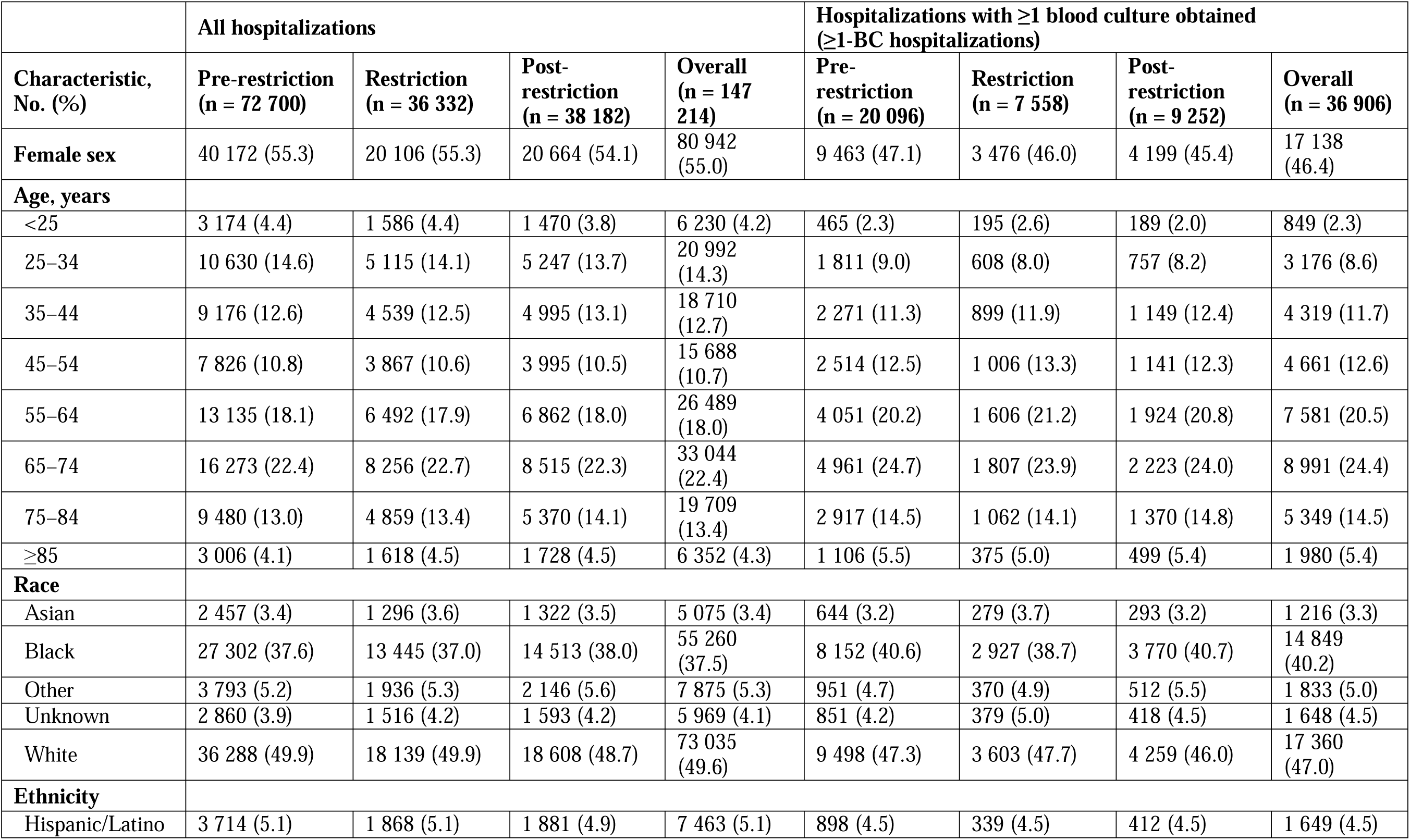
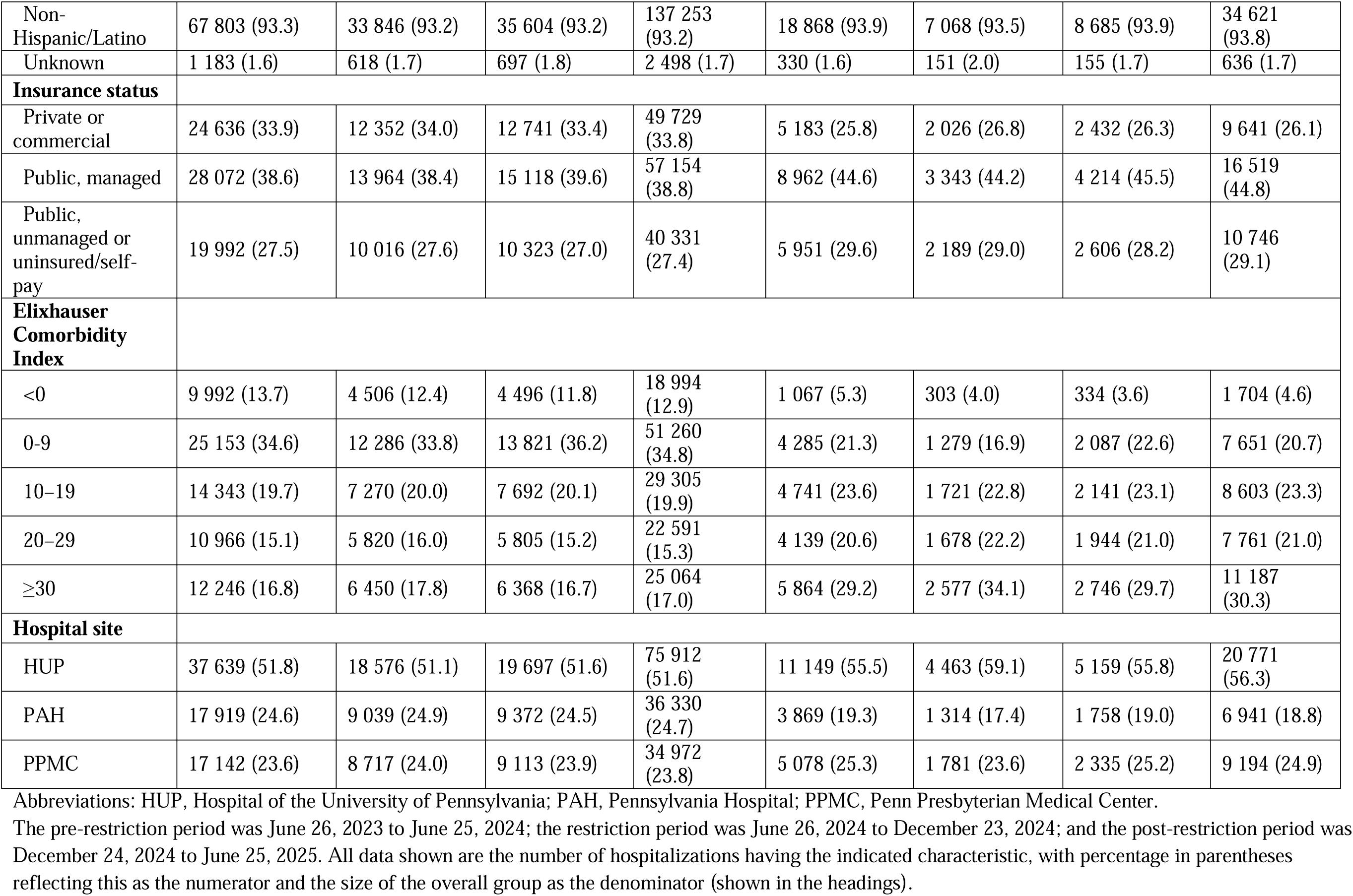
Sociodemographic and Clinical Characteristics of Patients Hospitalized at All 3 Hospital Sites Aggregated by Time Period.

### Primary Outcomes for All Hospitalizations

Across the 3 hospitals, the proportion of hospitalizations with in-hospital mortality or hospice discharge declined (**Table 2** and **Figure 1**) at –1.3% per week pre-restriction (P<.001) (**eTable 8**). We observed this trend across all hospitals except PAH, where the trend was nearly flat (**eTables 10–12** and **eFigures 3**, **6**, and **9**). During restriction, in-hospital mortality or hospice discharge flattened without significant change except for PPMC, where it had a –31.6% level decrease upon restriction onset (P=.048) and increased at +2.6% per week during restriction (P=.051) (**eTable 11**). After restriction, in-hospital mortality or hospice discharge decreased again at –2.8% per week (P<.001) and at all hospitals except PAH, which had a +91.8% increase upon post-restriction onset (P=.004). Thirty-day revisits were stable except for a post-restriction increase of +1.2% per week (P=.002) driven largely by PPMC with wide weekly variability. LOS increased and decreased at +0.2% and–0.7% per week during (P=.055) and post-restriction (P<.001), respectively; individual hospitals followed this trend although without statistical significance during restriction.

**Figure 1.**
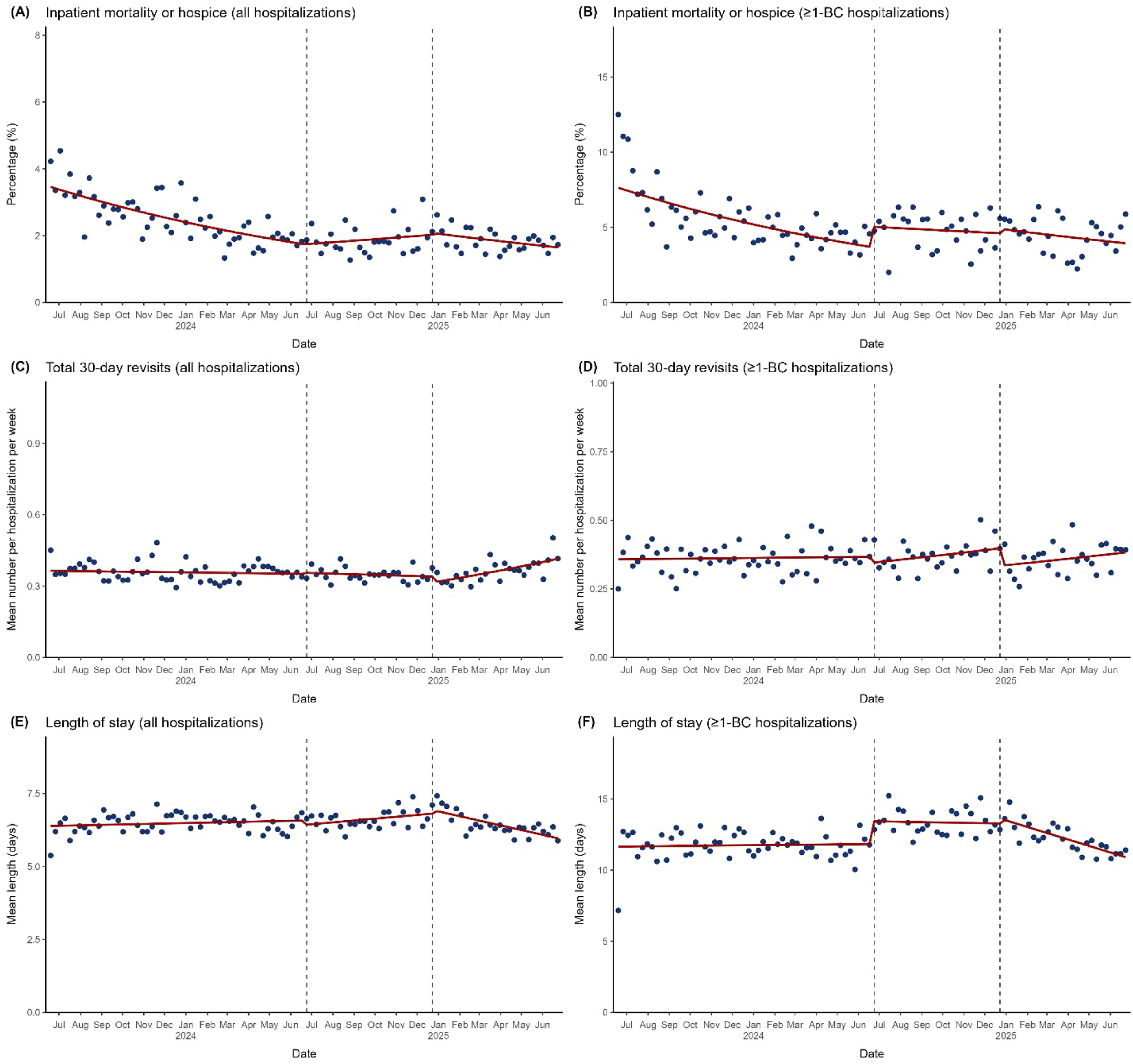
Changes in Primary Outcomes at All 3 Hospital Sites Combined. Blue dots represent weekly averages and the red solid line represents the fitted line from the segmented regression model assessing for abrupt level and slope changes at the start and stop dates of the blood culture restriction of 1 blood culture set per patient per 24 hours, which were June 26, 2024 and December 23, 2024, respectively, represented by dashed vertical lines. “≥1-BC hospitalizations” designates the hospitalizations with ≥1 blood culture obtained subgroup of all hospitalizations. The figure displays the following: (A) Percentage of inpatient mortality or hospice discharge for all hospitalizations (B) Percentage of inpatient mortality or hospice discharge for ≥1-BC hospitalizations (C) Mean number of total 30-day revisits for all hospitalizations (D) Mean number of total 30-day revisits for ≥1-BC hospitalizations (E) Mean length of stay for all hospitalizations (F) Mean length of stay for ≥1-BC hospitalizations

**Table 2.**
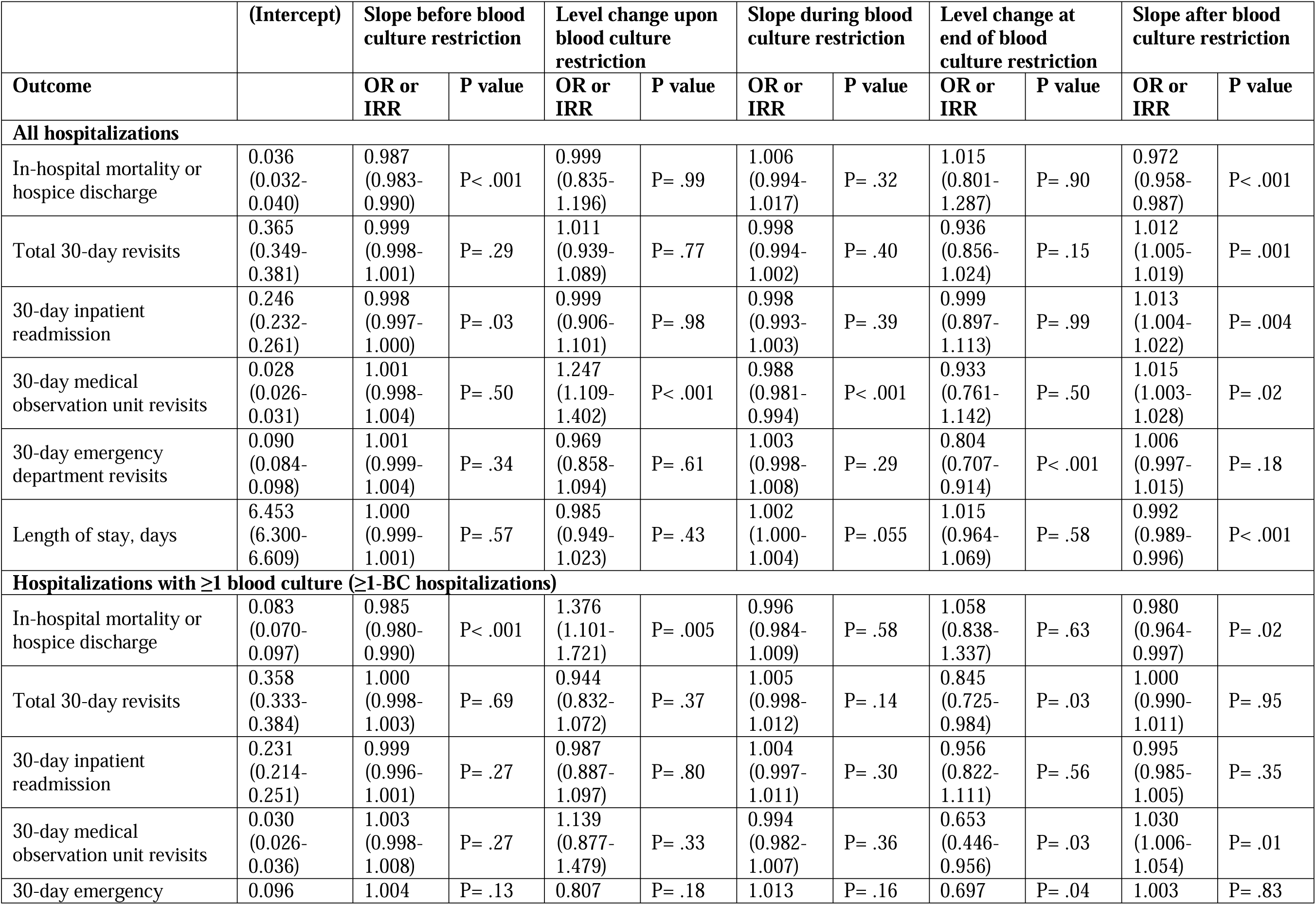
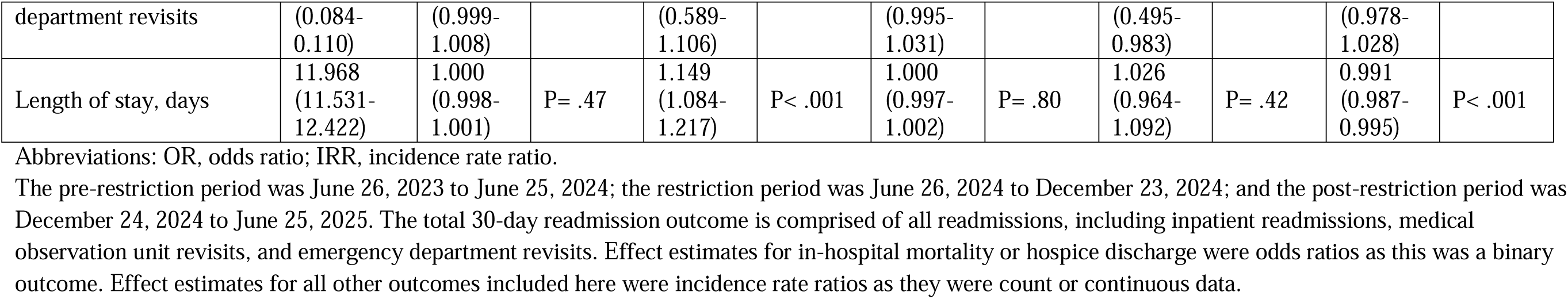
Effect Estimates of Blood Culture Restriction on Primary Outcomes for Hospitalizations at All 3 Hospital Sites Combined.

### Primary Outcomes for Hospitalizations with ≥1 Blood Culture Obtained

Among ≥1-BC hospitalizations, in-hospital mortality or hospice discharge followed trends similar to those for all hospitalizations, decreasing at –1.5% per week pre-restriction (P<.001), flattening during restriction, and decreasing at –2.0% per week post-restriction (P=.02) (**Table 2**, **eTable 9**, and **Figure 1**). There was a +37.6% increase upon restriction onset (P=.005), not observed for all hospitalizations. HUP and PPMC followed this trend to varying degrees of significance; at PAH, there were no significant changes (**eTables 10–12** and **eFigures 3**, **6**, and **9**). Frequency of 30-day revisits remained stable except for a –15.5% decrease upon post-restriction onset (P=.03). LOS increased +14.9% upon restriction onset (P<.001), remained stable during restriction, and declined at –0.9% per week post-restriction (P<.001). Individual hospitals followed similar trends to varying significance.

### Changes in Blood Culture Outcomes

Overall culture positivity was stable pre-restriction, immediately increased +36.2% upon restriction onset (P<.001), increased at +1.1% per week during restriction (P=.03), and decreased –27.0% upon post-restriction onset (P<.001) (**Table 3**, **eTable 8**, and **Figure 2**). Among all hospitalizations, the proportion of hospitalizations with ≥1 blood culture decreased –37.7% upon restriction onset (P<.001), increased at +0.9% per week during restriction (P=.002), and decreased at –1.3% per week post-restriction (P=.002).

**Figure 2.**
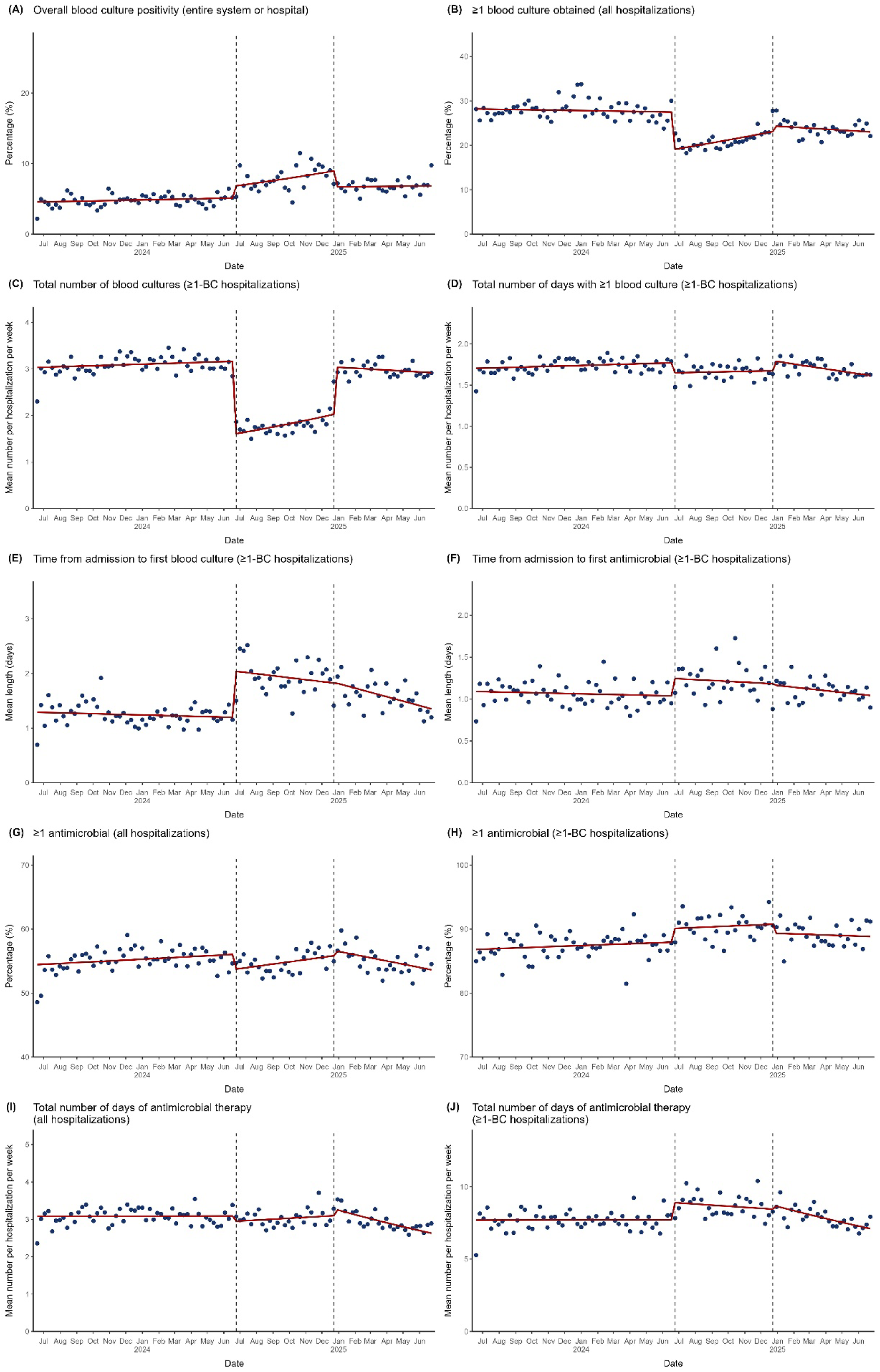
Changes in Selected Secondary Outcomes at All 3 Hospital Sites Combined. Blue dots represent weekly averages and the red solid line represents the fitted line from the segmented regression model assessing for abrupt level and slope changes at the start and stop dates of the blood culture restriction of 1 blood culture set per patient per 24 hours, which were June 26, 2024 and December 23, 2024, respectively, represented by dashed vertical lines. “≥1-BC hospitalizations” designates the hospitalizations with ≥1 blood culture obtained subgroup of all hospitalizations. The figure displays the following: (A) Percentage of overall blood culture positivity across all 3 hospital sites combined (B) Percentage of hospitalizations with ≥1 blood culture obtained for all hospitalizations (C) Mean number of blood cultures obtained for ≥1-BC hospitalizations (D) Mean number of days with ≥1 blood culture obtained for ≥1-BC hospitalizations (E) Mean length of time from admission to the first blood culture obtained for ≥1-BC hospitalizations (F) Mean length of time from admission to first antimicrobial administered for ≥1-BC hospitalizations (G) Percentage of hospitalizations with ≥1 antimicrobial administered for all hospitalizations (H) Percentage of hospitalizations with ≥1 antimicrobial administered for ≥1-BC hospitalizations (I) Mean number of days of non-broad-spectrum antimicrobial therapy for all hospitalizations (J) Mean number of days of non-broad-spectrum antimicrobial therapy for ≥1-BC hospitalizations

**Table 3.**
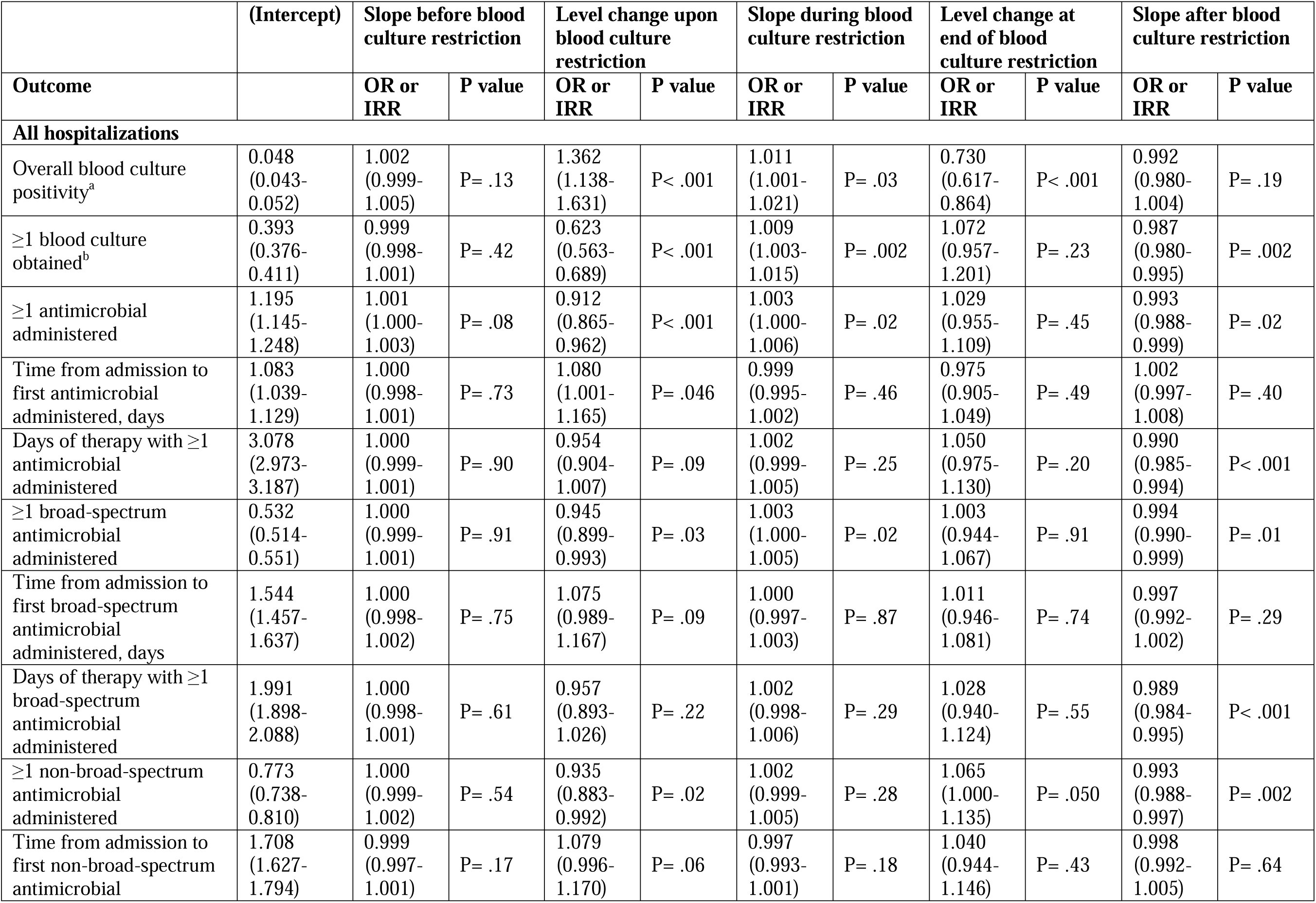
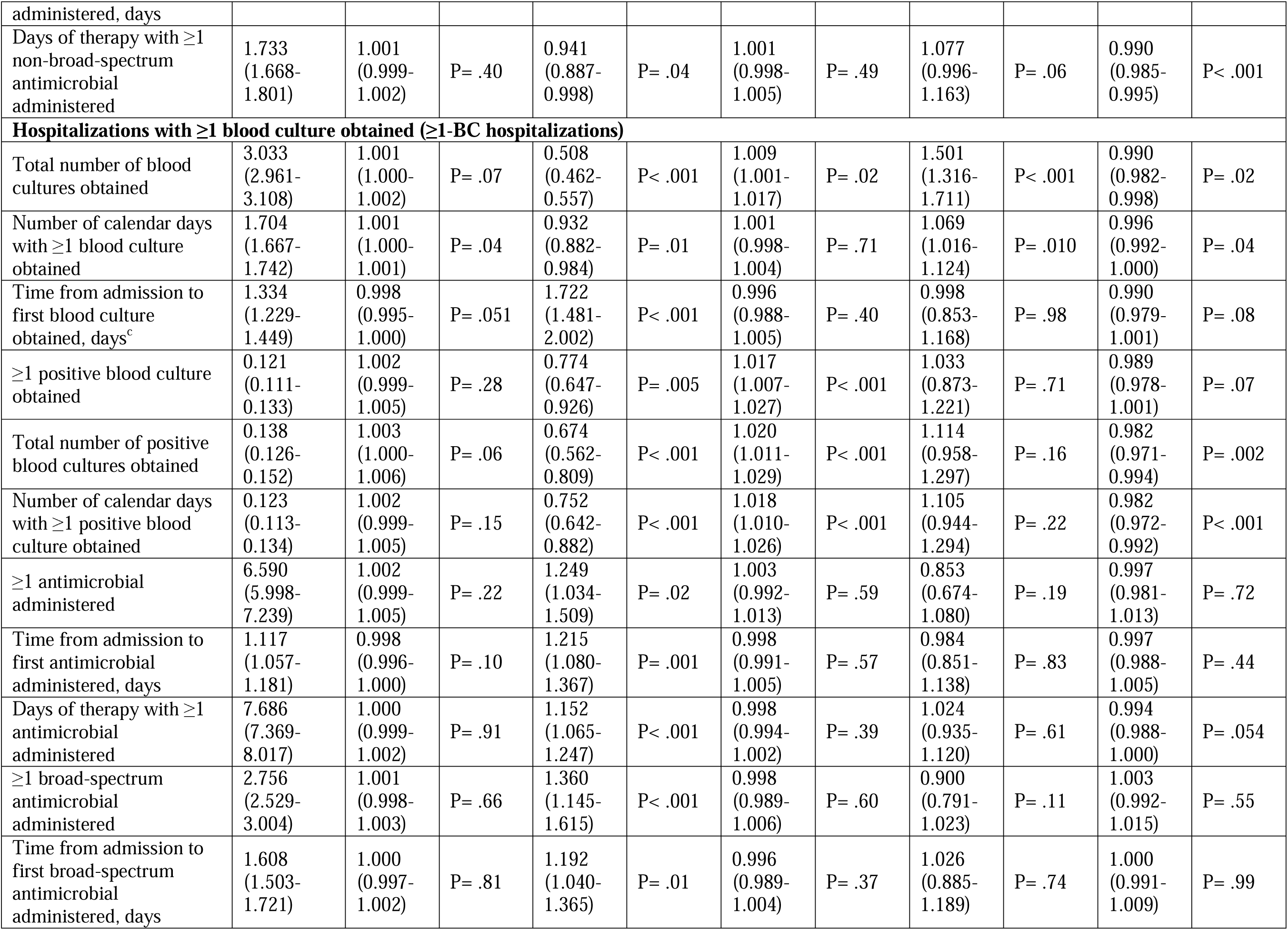
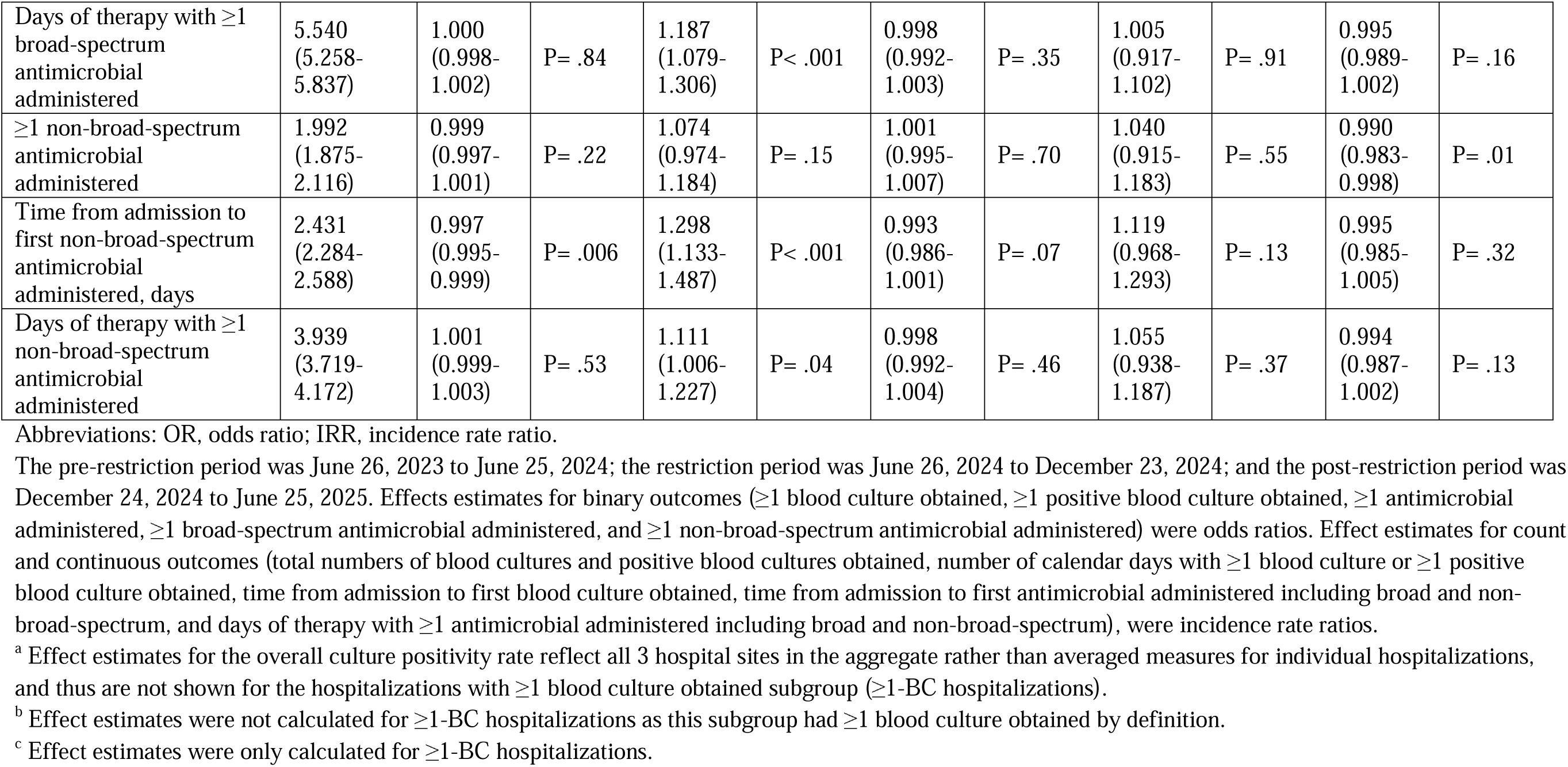
Effect Estimates of Blood Culture Restriction on Secondary Outcomes for Hospitalizations at All 3 Hospital Sites Combined.

Among ≥1-BC hospitalizations, the mean number of cultures per hospitalization per week decreased –49.2% upon restriction onset (P<.001), increased at +0.9% per week during restriction (P=.02) and +50.1% upon post-restriction onset (P<.001), and decreased at –1.0% per week post-restriction (P=.02) (**eTable 9**). Trends were similar for mean number of hospital days with ≥1 blood culture, proportion of hospitalizations with ≥1 positive blood culture, mean number of positive blood cultures, and mean number of hospital days with ≥1 positive blood culture obtained (**Table 3**, **eTable 9**, **Figure 2**, and **eFigure 1**). Mean length of time from admission to first blood culture obtained increased +72.2% upon restriction onset (P<.001), then decreased at –0.4% and –1.0% per week during and post-restriction, respectively, though estimates were underpowered (P=.40 and P=.08, respectively). Individual hospitals followed similar trends to varying significance (**eTables 10–12** and **eFigures 4**, **7**, and **10**).

### Changes in Antimicrobial Outcomes

Antimicrobial use was generally stable with modest shifts (**Table 3**, **eTables 8** and **9**, **Figure 2**, and **eFigure 2**). Among all hospitalizations, there were slight decreases in the proportion of hospitalizations with ≥1 antimicrobial administered upon restriction onset (–8.8% for any antimicrobial, –5.5% for broad-spectrum agents, and –6.5% for non-broad-spectrum agents, with P<.001, P=.03, and P=.02, respectively). Among ≥1-BC hospitalizations, there were corresponding increases (+24.9%, +36.0%, and +7.4% with P=.02, P<.001, and P=.15, respectively). Among all hospitalizations, the proportion of hospitalizations with ≥1 antimicrobial administered increased and decreased at +0.3% and –0.7% per week during and post-restriction, respectively (P=.02 for both) with similar trends for broad-spectrum and non-broad-spectrum agents; among ≥1-BC hospitalizations, the only significant change was a post-restriction decrease in non-broad-spectrum agent use at –1.0% per week (P=.01).

Among all hospitalizations, mean time from admission to first antimicrobial administered increased upon restriction onset (+8.0%, +7.5%, and +7.9% for any antimicrobial, broad-spectrum agents, and non-broad-spectrum agents with P=.048, P=.09, and P=.06, respectively). Among ≥1-BC hospitalizations, there were also corresponding increases (+21.5%, +19.2%, and +29.8% for any antimicrobial, broad-spectrum agents, and non-broad-spectrum agents with P=.001, P=.01, and P=.01, respectively).

Among all hospitalizations, the mean number of days of antimicrobial therapy decreased post-restriction (–1.0%, –1.1%, and –1.0% per week for any antimicrobial, broad-spectrum agents, and non-broad-spectrum agents with P=.001, P<.001, and P<.001, respectively), with a – 5.9% decrease in non-broad-spectrum agents upon restriction onset (P=.04). Among ≥1-BC hospitalizations, this metric increased upon restriction onset (+15.2%, +18.7%, and +11.1% for any antimicrobial, broad-spectrum agents, and non-broad-spectrum agents with P<.001, P<.001, and P=.04, respectively).

Individual hospitals followed similar trends for antimicrobial use outcomes to varying significance (**eTables 10–12** and **eFigures 5**, **8**, and **11**).

### Estimate of Excess In-Hospital Mortality or Hospice Discharge

We estimated excess in-hospital mortality or hospice discharge by extrapolating pre-restriction and restriction trends into the restriction and post-restriction periods, respectively (**eFigure 12**). Relative to pre-restriction, restriction was associated with an estimated 159 (4.4 per 1,000 hospitalizations; 95% CI, 2.0–6.7; P=.002) and 141 excess deaths or hospice discharges (18.7 per 1,000 hospitalizations; 95% CI, 10.1–26.2; P<.001) in all hospitalizations and ≥1-BC hospitalizations, respectively. Relative to restriction, post-restriction was associated with 122 (3.2 per 1,000 hospitalizations; 95% CI, –1.9 to 9.6; P=.23) and 1 excess death or hospice discharge (0.1 per 1,000 hospitalizations; 95% CI, –8.6 to 9.8; P=.97) in all hospitalizations and ≥1-BC hospitalizations, respectively.

### Sensitivity Analyses

Excluding hospitalizations during 1- and 2-week washout periods, we observed similar trends as in the main analysis with minimal change in effect estimates (**eTables 13 and 14**). We also observed similar trends when excluding hospitalizations with admission and discharge dates overlapping cutoff dates (**eTable 15** and **eFigures 13–15**). However, we observed a truncation effect at cutoff dates, likely due to selection bias affecting effect estimates, particularly for count or continuous outcomes. For example, while LOS in the main analysis slightly increased and decreased during restriction and post-restriction, respectively (**Table 2**), the sensitivity analysis showed markedly different estimates, with statistically significant slope decreases pre-restriction and during restriction and level increases upon restriction and post-restriction onset (**eTable 15**).

## Discussion

The single-set blood culture restriction for each patient every 24 hours had immediate significant effects, including increased overall culture positivity and decreased culture use. The decrease in hospitalizations with ≥1 blood culture obtained and increased length of time from admission to first culture collection suggest increased hesitancy by clinicians to order blood cultures. Subsequently, among ≥1-BC hospitalizations, we observed increased length of time from admission to first antimicrobial. Taken together, these suggest that increased hesitancy to order blood cultures led to delayed diagnosis and treatment, providing a likely mechanistic reason for the immediate increase in mortality and hospice discharge upon restriction onset.

Among all hospitalizations, in-hospital mortality and hospice discharge had been declining prior to restriction, a trend consistent with national patterns following the COVID-19 pandemic.^39–41^ That this decline flattened during the blood culture restriction period and resumed afterward also suggests worsened outcomes due to the restriction, possibly due to underdiagnosis, delayed treatment, or undertreatment of bloodstream infections, with estimated excess deaths or hospice discharges estimated at 4.4 and 18.7 per 1,000 hospitalizations among all hospitalizations and ≥1-BC hospitalizations, respectively. Antimicrobial use decreased overall but increased among ≥1-BC hospitalizations, possibly reflecting fewer perceived indications overall and higher acuity among those tested.

Many blood culture metrics returned to pre-restriction baseline levels, but not all did: for example, the proportion of hospitalizations with ≥1 blood culture obtained started at 28.2% pre-restriction and ended at 23.0% by end of the study period (calculated using effect estimates), while overall culture positivity started at 4.4% and ended at 6.4%. The incomplete return to baseline may have been due to residual effects of the restriction and the new blood culture order panel that simultaneously came into effect with restriction withdrawal. That in-hospital mortality or hospice discharge continued declining even with fewer blood cultures being ordered than during pre-restriction suggests that it is possible to lessen blood culture ordering through diagnostic stewardship interventions without worsening patient outcomes.

Notably, trends were not uniform across hospitals. For example, at HUP and PPMC, in-hospital mortality and hospice discharge decreased pre- and post-restriction while flattening during restriction, whereas at PAH there was no significant change. Possible explanations include smaller sample size or differences in case mix, baseline severity of illness, or staffing models (e.g., differing proportions of advanced practice providers or house officers).

Other hospitals have reported various responses to the shortage. Some, including Vanderbilt University Medical Center, Mount Sinai Health System (New York City), MaineHealth, Virginia Commonwealth University Health System, and Eskenazi Health, used some combination of educational efforts, EHR order-set changes, and clinical decision-support tools, generally reporting subsequent decreased culture use.^21–26^ At Nagoya City University East Medical Center during the shortage, clinicians were instructed, though not required, to collect only 1 blood culture set per patient, with exceptions for suspected catheter-related bloodstream infections or infective endocarditis. This significantly reduced not only blood culture usage but also overall culture positivity, raising concerns for possible underdiagnosis of bloodstream infections, though patient outcomes were not assessed.^27^ Vanderbilt and Fujita Health University Hospital also used single-set blood culture restrictions albeit to a more limited degree with exceptions, e.g., neutropenia^22,28^; a follow up study at Vanderbilt found no change in 30-day all-cause mortality, LOS, or 30-day readmissions, though this was limited to *Staphylococcus aureus* bacteremia, focusing primarily on culture metrics.^23^ Mount Sinai also reported no change in LOS or in-hospital mortality though this was limited to ED patients.^24^

More recently, a CDC-based team conducted a National Healthcare Safety Network questionnaire, finding that over 75% of responding facilities reported being affected by the shortage.^29^ Furthermore, they conducted a retrospective cohort study using administrative data at 11 facilities solely reliant on BD BACTEC^™^ blood culture bottles and 28 facilities not solely reliant, finding comparatively decreased culture usage and overall culture positivity in the former group. However, this study focused on blood culture metrics, not measuring patient outcomes such as mortality.

Our study is distinctive in that the single-set blood culture restriction applied to nearly all patients, regardless of setting and without condition-specific exceptions. We included a broad range of outcomes, including primary clinical outcomes, blood culture use, and antimicrobial administration. We demonstrated a significant adverse shift in primary outcomes, notably increased in-hospital mortality or hospice discharge, warranting heightened caution when considering similar restrictions for diagnostic stewardship. Future studies could explore context-specific restrictions and recommendations, as our own institution’s blood culture order panel attempts to encourage (e.g., recommending not obtaining blood cultures to document clearance of uncomplicated gram-negative bacteremia versus obtaining ≥2 sets for culture-proven *Staphylococcus aureus* bacteremia), in addition to broader, ongoing educational efforts on appropriate blood culture use.^19,20^ Our study also reinforces the need to build supply chain resiliency, not only for drugs and medications^5,9^ but also for critical diagnostic testing supplies^42^ such as blood culture bottles.^30^

### Limitations

This study had certain limitations. First, its single-center design, while conducted across 3 hospitals, limits generalizability, particularly without a simultaneous comparator group. Second, we did not evaluate additional outcomes of interest, such as blood culture appropriateness, sepsis, intensive care admission, post-discharge mortality, blood culture contamination rates, and antimicrobial appropriateness. Third, we did not account for changes at other nearby health systems that may have influenced outcomes, such as shared patient readmissions resulting in undercounted 30-day revisit rates. Fourth, while baseline characteristics were largely similar across the pre-restriction, restriction, and post-restriction periods, unmeasured factors such as geographic residence could have influenced outcomes. Fifth, some subgroup analyses were underpowered. Sixth, secular or seasonal trends not captured in our models could have also influenced results.

## Conclusions

The 2024 national blood culture bottle shortage compelled our health system to implement a near-universal single-set blood culture restriction, which was associated with significantly worse patient outcomes, including increased in-hospital mortality or hospice discharge. Future work is warranted both to determine optimal blood culture stewardship practices and to strengthen supply chain resiliency for blood culture bottles and other critical diagnostics.

## Data Availability

All data produced in the present study are available upon reasonable request to the authors.

## References

1. Pandey AK, Cohn J, Nampoothiri V, et al. A systematic review of antibiotic drug shortages and the strategies employed for managing these shortages. Clinical Microbiology and Infection. 2025;31(3):345–353. doi:10.1016/j.cmi.2024.09.023

2. Bartoo AS, Gilmer MA, Tichy EM. Antimicrobial Shortages: A Global Issue Impacting Infectious Diseases. Clinical Infectious Diseases. 2025;80(2):249–252. doi:10.1093/CID/CIAE498

3. Cahan E. IV Fluid Shortages Persist Months After Hurricane Helene Hit a Supplier— Hospitals Have Had to Adapt. JAMA. 2025;333(24):2127–2130. doi:10.1001/JAMA.2025.0075

4. Callaway Kim K, Rothenberger SD, Tadrous M, et al. Drug Shortages Prior to and During the COVID-19 Pandemic. JAMA Netw Open. 2024;7(4):e244246–e244246. doi:10.1001/JAMANETWORKOPEN.2024.4246

5. Serchen J, Hilden D, Silberger JR, et al. Bolstering the Medication Supply Chain and Ameliorating Medication Shortages: A Position Paper From the American College of Physicians. Ann Intern Med. Published online August 12, 2025. doi:10.7326/ANNALS-25-00607

6. Aronson JK, Heneghan C, Ferner RE. Drug shortages. Part 1. Definitions and harms. Br J Clin Pharmacol. 2023;89(10):2950–2956. doi:10.1111/BCP.15842

7. Aronson JK, Heneghan C, Ferner RE. Drug shortages. Part 2: Trends, causes and solutions. Br J Clin Pharmacol. 2023;89(10):2957–2963. doi:10.1111/BCP.15853

8. Park M, Conti RM, Wosińska ME, Ozlem E, Hopp WJ, Fox ER. Building Resilience Into US Prescription Drug Supply Chains. Health Affairs Forefront. Published online January 30, 2023. doi:10.1377/FOREFRONT.20230126.864137

9. Shahzad M, Nogueira LM, Wagner A. Threats of Weather Disasters for Drug Manufacturing Facilities in the US. JAMA. Published online August 20, 2025. doi:10.1001/JAMA.2025.13843

10. Pineda-Moncusí M, Rekkas A, Pérez ÁM, et al. Changes in use and utilisation patterns of drugs with reported shortages between 2010 and 2024 in Europe and North America: a network cohort study. Lancet Public Health. 2025;10(10):e835–e847. doi:10.1016/S2468-2667(25)00194-X

11. U.S. Food & Drug Administration. FDA Roundup: July 12, 2024.; 2024. Accessed August 21, 2025. https://www.fda.gov/news-events/press-announcements/fda-roundup-july-12-2024

12. Centers for Disease Control and Prevention. Disruptions in Availability of BD BACTEC Blood Culture Bottles: Current Situation.; 2024. Accessed August 21, 2025. https://www.cdc.gov/healthcare-associated-infections/bd-bactec-availability/index.html

13. Morgan DJ, Malani P, Diekema DJ. Diagnostic Stewardship—Leveraging the Laboratory to Improve Antimicrobial Use. JAMA. 2017;318(7):607–608. doi:10.1001/JAMA.2017.8531

14. Sullivan K V. Diagnostic Stewardship in Clinical Microbiology, Essential Partner to Antimicrobial Stewardship. Clin Chem. 2021;68(1):75–82. doi:10.1093/CLINCHEM/HVAB206

15. Fabre V, Klein E, Salinas AB, et al. A diagnostic stewardship intervention to improve blood culture use among adult nonneutropenic inpatients: The DISTRIBUTE study. J Clin Microbiol. 2020;58(10):1053–1073. doi:10.1128/JCM.01053-20

16. Woods-Hill CZ, Colantuoni EA, Koontz DW, et al. Association of Diagnostic Stewardship for Blood Cultures in Critically Ill Children With Culture Rates, Antibiotic Use, and Patient Outcomes: Results of the Bright STAR Collaborative. JAMA Pediatr. 2022;176(7):690–698. doi:10.1001/JAMAPEDIATRICS.2022.1024

17. Fabre V, Davis A, Diekema DJ, et al. Principles of diagnostic stewardship: A practical guide from the Society for Healthcare Epidemiology of America Diagnostic Stewardship Task Force. Infect Control Hosp Epidemiol. 2023;44(2):178–185. doi:10.1017/ICE.2023.5

18. Dräger S, Giehl C, Søgaard KK, et al. Do we need blood culture stewardship programs? A quality control study and survey to assess the appropriateness of blood culture collection and the knowledge and attitudes among physicians in Swiss hospitals. Eur J Intern Med. 2022;103:50–56. doi:10.1016/J.EJIM.2022.04.028

19. Fabre V, Carroll KC, Cosgrove SE. Blood Culture Utilization in the Hospital Setting: a Call for Diagnostic Stewardship. J Clin Microbiol. 2022;60(3). doi:10.1128/JCM.01005-21

20. Ryder JH, Van Schooneveld TC, Diekema DJ, Fabre V. Every Crisis Is an Opportunity: Advancing Blood Culture Stewardship During a Blood Culture Bottle Shortage. Open Forum Infect Dis. 2024;11(9). doi:10.1093/OFID/OFAE479

21. Ezran C, Herrle E, Yen CF, Mercuro NJ, Diekema DJ, Gordon LB. Shortage as a catalyst for high-value care: Evaluation of a blood culture stewardship intervention driven by supply chain disruption. J Hosp Med. Published online August 16, 2025. doi:10.1002/JHM.70158

22. Humphries RM, Wright PW, Banerjee R, et al. Rapid Implementation of Blood Culture Stewardship: Institutional Response to an Acute National Blood Culture Bottle Shortage. Clin Infect Dis. 2025;80(2). doi:10.1093/CID/CIAE402

23. Humphries RM, Banerjee R, Dupont WD, et al. Association Between Blood Culture Bottle Shortage and Ordering Restrictions and Clinical Outcomes for Patients With Staphylococcus aureus Bacteremia. Open Forum Infect Dis. 2025;12(9). doi:10.1093/OFID/OFAF546

24. Takkavatakarn K, Patel G, Oh W, et al. Electronic clinical decision support system guided blood culture stewardship in emergency departments: response to the national blood culture media shortage. Infect Control Hosp Epidemiol. 2025;46(6):650–653. doi:10.1017/ICE.2025.83

25. Doern CD, Whitman M, Doll M, et al. Blood culture bottle shortage mitigation efforts: analysis of impact on ordering and patient impact. Antimicrobial stewardship & healthcare epidemiology : ASHE. 2025;5(1). doi:10.1017/ASH.2024.474

26. Butt S, Kressel AB, Haines BL, et al. Rapid implementation of a clinical decision-support workflow during the national blood culture bottle shortage. Infection Prevention in Practice. 2024;6(4). doi:10.1016/j.infpip.2024.100417

27. Itoh N, Akazawa-Kai N, Okumura N, Kuriki S, Wachino C, Kawabata T. Impact of BD BACTEC blood culture bottle shortage on performance metrics: An interrupted time-series analysis at a Japanese university-affiliated hospital. J Infect Chemother. 2025;31(4). doi:10.1016/J.JIAC.2025.102664

28. Hanai S, Shintani C, Higashimoto Y, Uehara Y, Doi Y, Honda H. Restoring the 2-set blood culture practice after the resolution of supply shortage. Infect Control Hosp Epidemiol. Published online 2025. doi:10.1017/ICE.2025.60

29. Lutgring JD, Maillis A, Bryant GC, et al. The Impact of a Nationwide Blood Culture Bottle Shortage in 2024 on Healthcare Facilities in the United States. Clin Infect Dis. Published online September 10, 2025. doi:10.1093/CID/CIAF498

30. Miller JM, Binnicker MJ, Campbell S, et al. Guide to Utilization of the Microbiology Laboratory for Diagnosis of Infectious Diseases: 2024 Update by the Infectious Diseases Society of America (IDSA) and the American Society for Microbiology (ASM). Clin Infect Dis. Published online March 5, 2024. doi:10.1093/CID/CIAE104

31. Fabre V, Sharara SL, Salinas AB, Carroll KC, Desai S, Cosgrove SE. Does This Patient Need Blood Cultures? A Scoping Review of Indications for Blood Cultures in Adult Nonneutropenic Inpatients. Clin Infect Dis. 2020;71(5):1339–1347. doi:10.1093/CID/CIAA039

32. von Elm E, Altman DG, Egger M, Pocock SJ, Gøtzsche PC, Vandenbroucke JP. The Strengthening the Reporting of Observational Studies in Epidemiology (STROBE) statement: guidelines for reporting observational studies. J Clin Epidemiol. 2008;61(4):344–349. doi:10.1016/j.jclinepi.2007.11.008

33. Elixhauser A, Steiner C, Harris DR, Coffey RM. Comorbidity Measures for Use with Administrative Data. Med Care. 1998;36(1):8–27. doi:10.1097/00005650-199801000-00004

34. Gasparini A. comorbidity: An R package for computing comorbidity scores. J Open Source Softw. 2018;3(23):648. doi:10.21105/JOSS.00648

35. Quan H, Sundararajan V, Halfon P, et al. Coding algorithms for defining comorbidities in ICD-9-CM and ICD-10 administrative data. Med Care. 2005;43(11):1130–1139. doi:10.1097/01.MLR.0000182534.19832.83

36. Newey W, West K. A Simple, Positive Semi-Definite, Heteroskedasticity and Autocorrelation Consistent Covariance Matrix. Econometrica. 1987;55(3):708. doi:10.2307/1913610

37. Cumby RE, Huizinga J. Testing the Autocorrelation Structure of Disturbances in Ordinary Least Squares and Instrumental Variables Regressions. Econometrica. 1992;60(1):195. doi:10.2307/2951684

38. R Core Team. R: A Language and Environment for Statistical Computing.; 2023. https://www.R-project.org/

39. Teasdale B, Narayan A, Harman S, Schulman KA. Place of Death Before and During the COVID-19 Pandemic. JAMA Netw Open. 2024;7(1):e2350821–e2350821. doi:10.1001/JAMANETWORKOPEN.2023.50821

40. Chen W. COVID-19 Surges and Hospital Outcomes in the United States. American Journal of Managed Care. 2022;28(11):E399–E404. doi:10.37765/AJMC.2022.89264,

41. Minhas AMK, Fudim M, Michos ED, Abramov D. Has mortality in the United States returned to pre-pandemic levels? An analysis of provisional 2023 data. J Intern Med. 2024;296(2):168–176. doi:10.1111/JOIM.13811

42. Weber DJ, Malani AN, Shenoy ES, et al. Society for Healthcare Epidemiology of America position statement on pandemic preparedness for policymakers: Mitigating supply shortages. Infect Control Hosp Epidemiol. 2024;45(7):813–817. doi:10.1017/ICE.2024.67

