## Supplement for "Single-Set Blood Culture Restriction During the 2024 National Blood Culture Bottle Shortage: An Interrupted Time Series Analysis of Patient Outcomes"

**Date:** September 23, 2025

### **Table of Contents**

**eMethods 1.** Blood Culture Order Panel Implemented in the Post-Restriction Period.

**eMethods 2.** Categorization of Antimicrobials.

**eFigure 1.** Changes in Other Blood Culture Outcomes at All 3 Hospital Sites Combined.

**eFigure 2.** Changes in Other Antimicrobial Outcomes at All 3 Hospital Sites Combined.

**eFigure 3.** Changes in Primary Outcomes at the Hospital of the University of Pennsylvania.

**eFigure 4.** Changes in Blood Culture Outcomes at the Hospital of the University of Pennsylvania.

**eFigure 5.** Changes in Antimicrobial Outcomes at the Hospital of the University of Pennsylvania.

**eFigure 6.** Changes in Primary Outcomes at Penn Presbyterian Medical Center.

**eFigure 7.** Changes in Blood Culture Outcomes at Penn Presbyterian Medical Center.

**eFigure 8.** Changes in Antimicrobial Outcomes at Penn Presbyterian Medical Center.

**eFigure 9.** Changes in Primary Outcomes at Pennsylvania Hospital.

**eFigure 10.** Changes in Blood Culture Outcomes at Pennsylvania Hospital.

**eFigure 11.** Changes in Antimicrobial Outcomes at Pennsylvania Hospital.

**eFigure 12.** Excess In-Hospital Mortality or Hospice Discharge at All 3 Hospital Sites Combined.

**eFigure 13.** Changes in Primary Outcomes at All 3 Hospital Sites Combined, Excluding Hospitalizations Overlapping Restriction Cutoff Dates.

**eFigure 14.** Changes in Blood Culture Outcomes at All 3 Hospital Sites Combined, Excluding Hospitalizations Overlapping Restriction Cutoff Dates.

**eFigure 15.** Changes in Antimicrobial Outcomes at All 3 Hospital Sites Combined, Excluding Hospitalizations Overlapping Restriction Cutoff Dates.

**eTable 1.** Sociodemographic and Clinical Characteristics of Patients Hospitalized at the Hospital of the University of Pennsylvania.

**eTable 2.** Sociodemographic and Clinical Characteristics of Patients Hospitalized at Penn Presbyterian Medical Center.

**eTable 3.** Sociodemographic and Clinical Characteristics of Patients Hospitalized at Pennsylvania Hospital.

**eTable 4.** Primary and Secondary Outcomes at All 3 Hospital Sites Combined Aggregated by Time Period.

**eTable 5.** Primary and Secondary Outcomes at the Hospital of the University of Pennsylvania Aggregated by Time Period.

**eTable 6.** Primary and Secondary Outcomes at Penn Presbyterian Medical Center Aggregated by Time Period.

**eTable 7.** Primary and Secondary at Pennsylvania Hospital Aggregated by Time Period.

**eTable 8.** Interpretation of Effect Estimates of Blood Culture Restriction on Primary and Secondary Outcomes During All Hospitalizations at All 3 Hospital Sites Combined.

**eTable 9.** Interpretation of Effect Estimates of Blood Culture Restriction on Primary and Secondary Outcomes During All Hospitalizations with ≥1 Blood Culture Obtained at All 3 Hospital Sites Combined.

**eTable 10.** Effect Estimates of Blood Culture Restriction on Primary and Secondary Outcomes for Hospitalizations at the Hospital of the University of Pennsylvania.

**eTable 11.** Effect Estimates of Blood Culture Restriction on Primary and Secondary Outcomes for Hospitalizations at Penn Presbyterian Medical Center.

**eTable 12.** Effect Estimates of Blood Culture Restriction on Primary and Secondary Outcomes for Hospitalizations at Pennsylvania Hospital.

**eTable 13.** Effect Estimates of Blood Culture Restriction on Primary and Secondary Outcomes for Hospitalizations at All 3 Hospital Sites Combined with Washout Period of 1 Week.

**eTable 14.** Effect Estimates of Blood Culture Restriction on Primary and Secondary Outcomes for Hospitalizations at All 3 Hospital Sites Combined with Washout Period of 2 Weeks.

**eTable 15.** Effect Estimates of Blood Culture Restriction on Primary and Secondary Outcomes for Hospitalizations at All 3 Hospital Sites Combined, Excluding Hospitalizations Overlapping Restriction Cutoff Dates.

### **eMethods 1.** Blood Culture Order Panel Implemented in the Post-Restriction Period.

On December 23, 2024, when the single-set blood culture restriction was withdrawn, a new blood culture order panel went into effect in the electronic health record. This order panel provides different options depending on whether the patient has 1. neutropenic fever or active liquid malignancy, 2. history of solid organ transplant, or 3. none of the above. It is based on findings from a multicenter study through the CDC Prevention Epicenter Program and the Society for Healthcare Epidemiology of America Research Network that provided a framework for evidence-based blood culture order panels (Fabre et al, *J Clin Microbiol,* 2020 and Fabre et al, *Clin Infect Dis*, 2020).

Below are example screenshots of the order panel for a patient without neutropenic fever, active liquid malignancy, or history of solid organ transplant, encouraging 2 sets of peripheral blood cultures rather than from central lines, and discouraging collection if the indication is not one of the listed options, which include new onset sepsis or septic shock or evaluation for a suspected new bloodstream infection for a multiple possible reasons (e.g. endocarditis, meningitis). It discourages blood culture collection for a variety of what our institution considered low-yield indications (e.g. cellulitis, aspiration events). For documented clearance of a bloodstream infection, it encourages two sets for organisms such as *Staphylococcus aureus* but discourages collecting blood cultures for other organisms such as *Streptococcus pyogenes*.

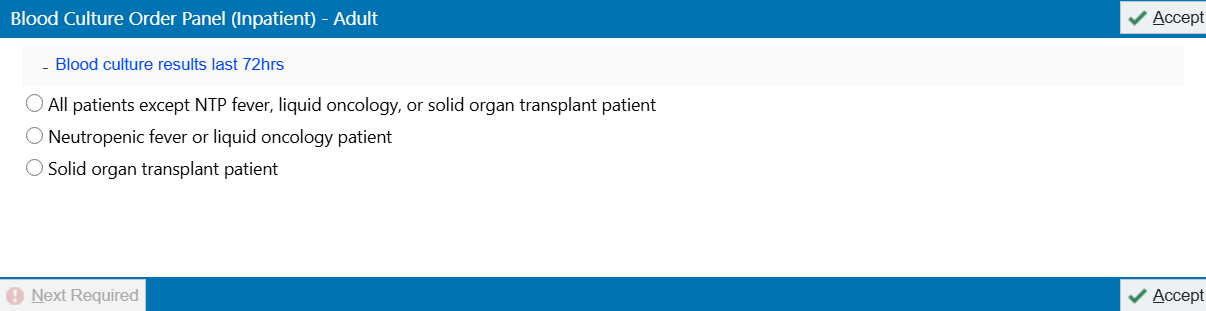

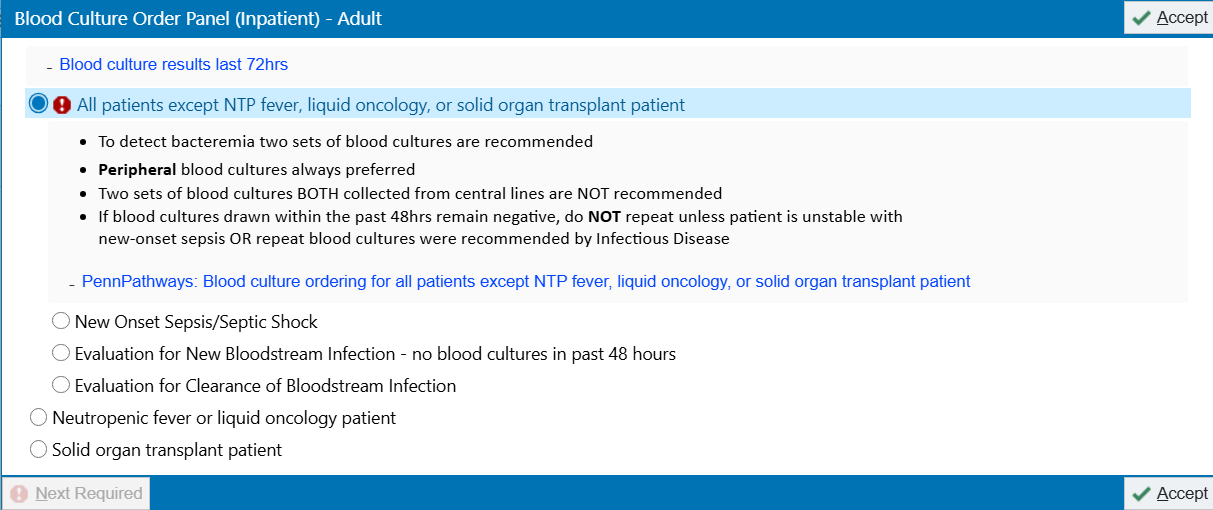

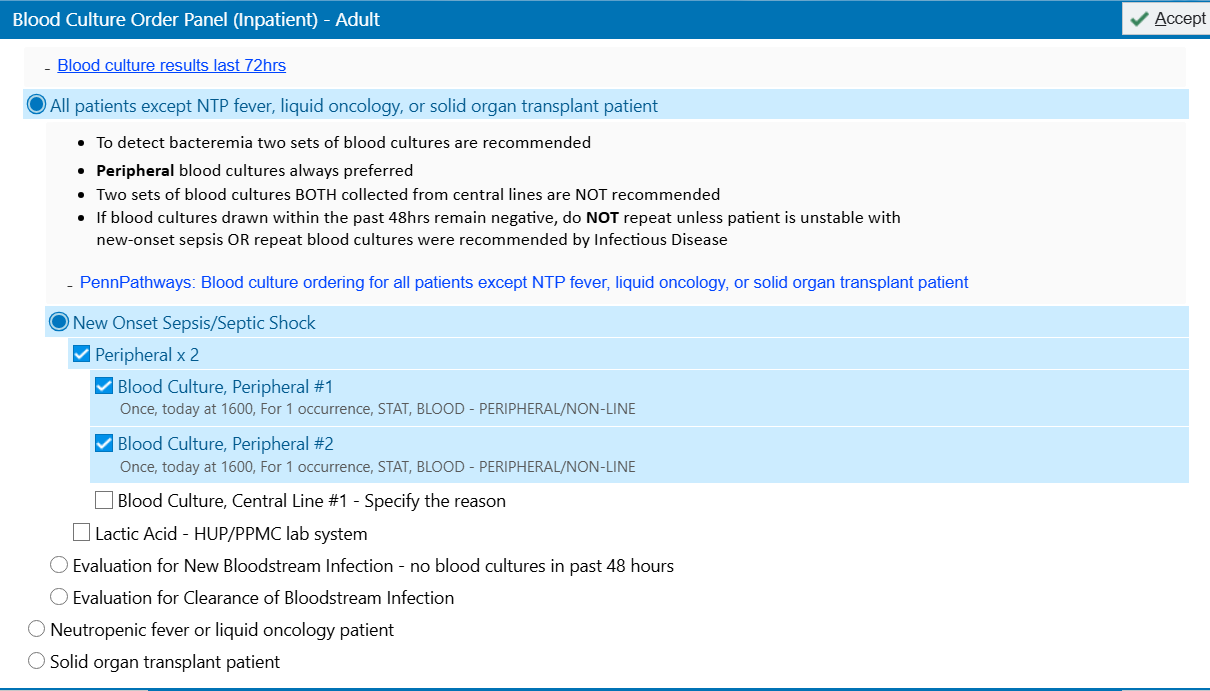

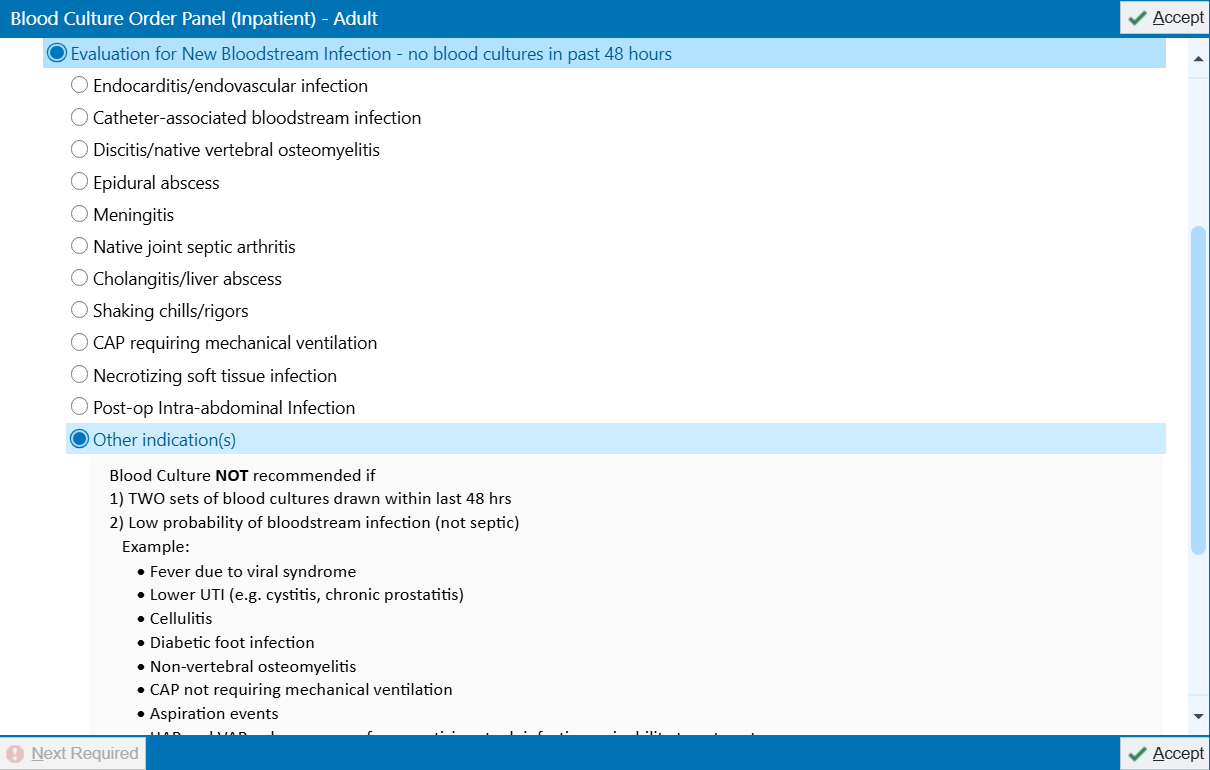

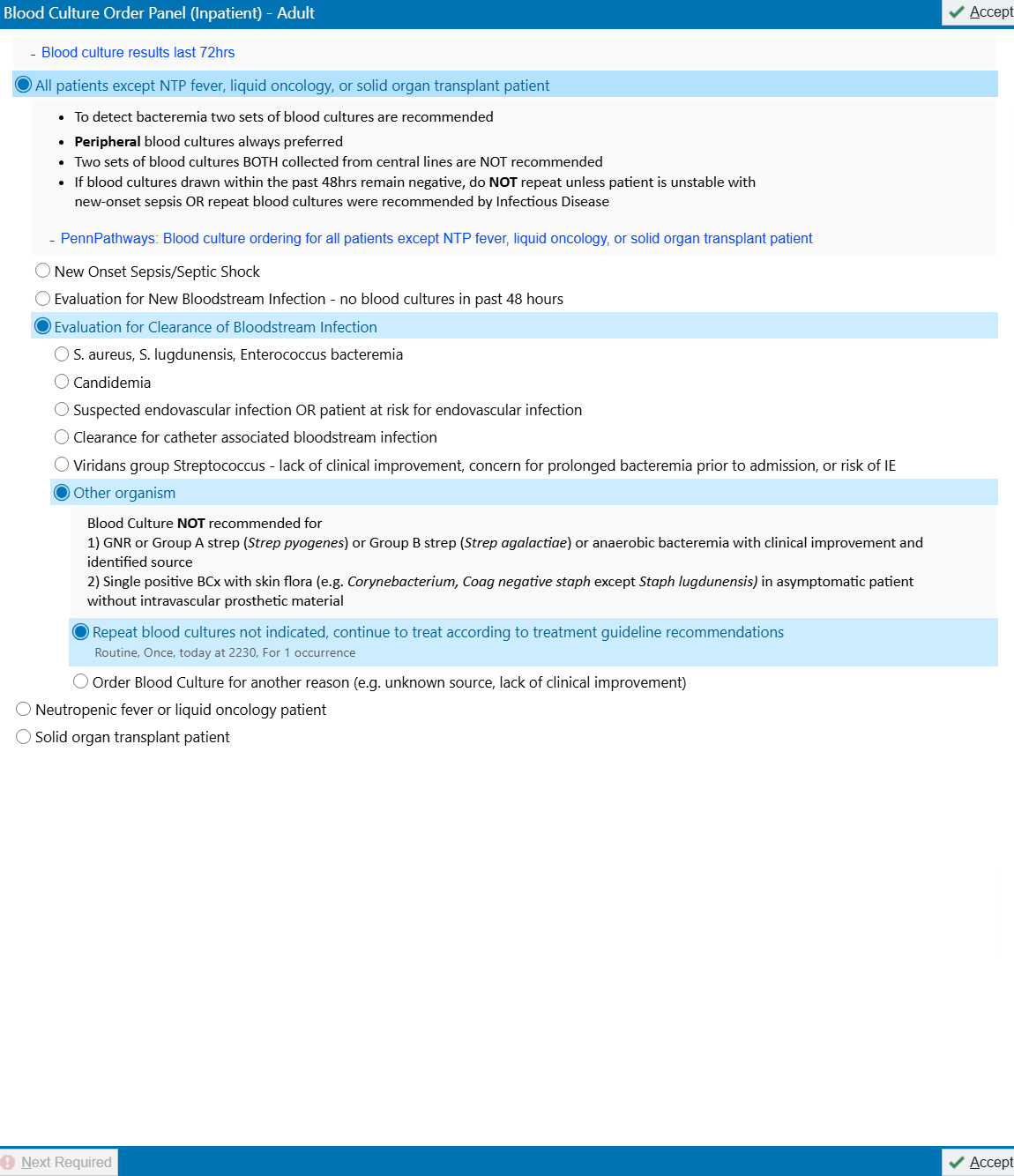

Below is an example screenshot of the order panel for a patient with neutropenic fever or active liquid malignancy, recommending 2 sets of cultures, 1 peripheral and 1 central if a central line is present, or 2 peripheral cultures if there is no central line.

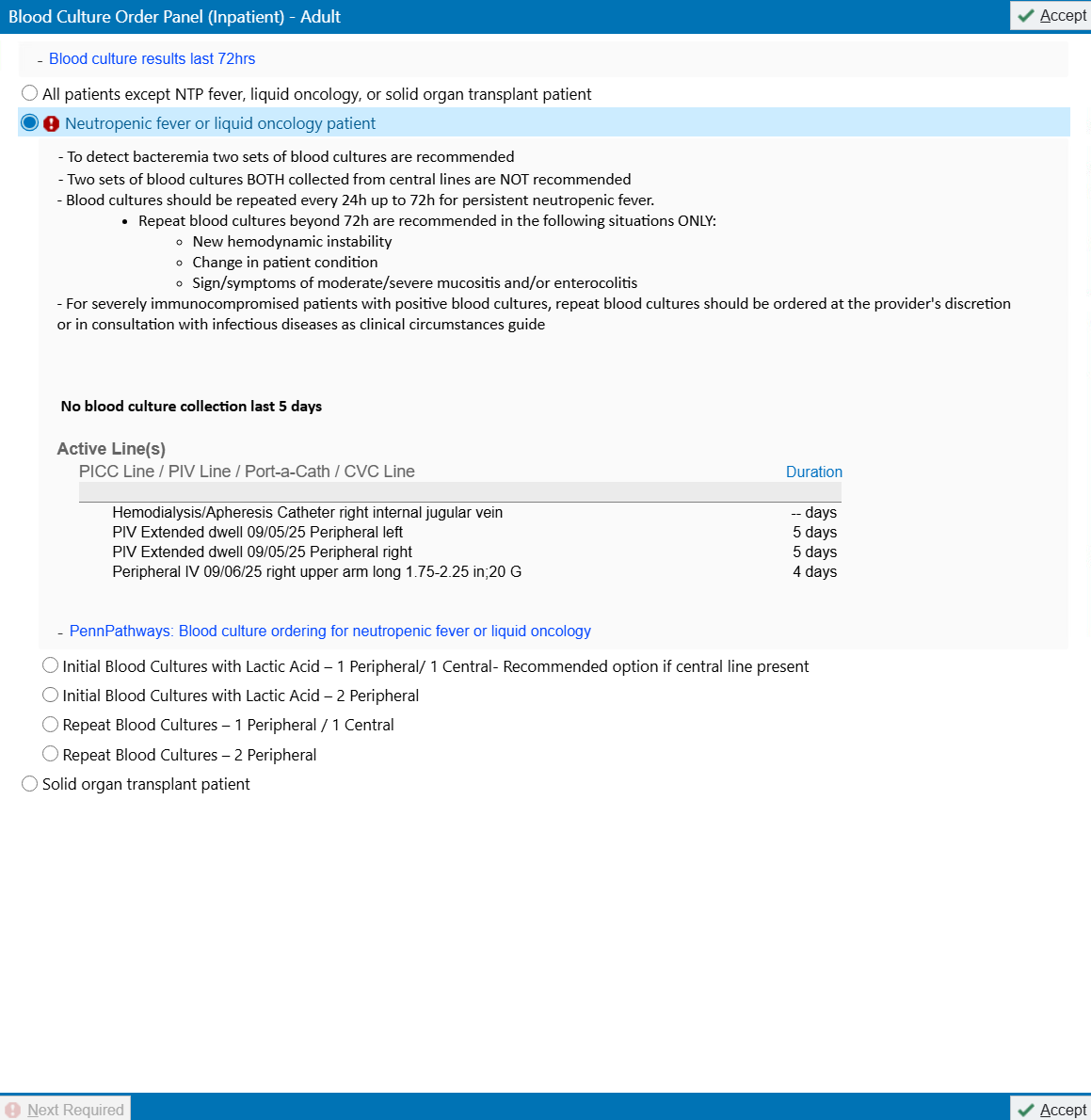

Below is an example screenshot of the order panel for a patient with history of solid organ transplant, recommending 2 sets of peripheral blood cultures rather than from central lines and prompting the clinician to provide a reason if a blood culture from a central line is ordered.

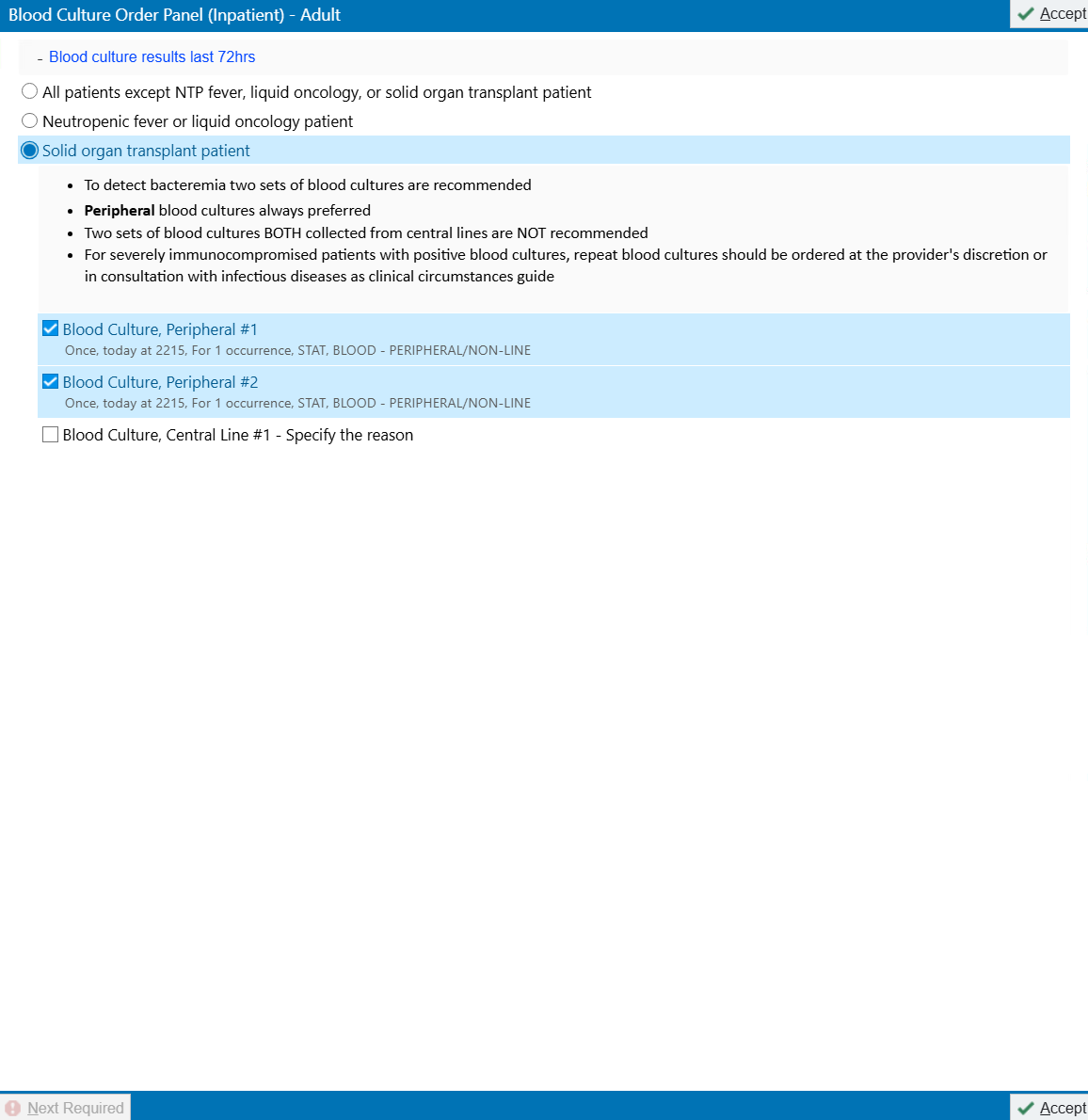

### **eMethods 2.** Categorization of Antimicrobials.

Antimicrobials administered during hospitalizations were obtained from our institution’s electronic health record using Epic Clarity. Topical antimicrobials and antivirals were not included. Antimicrobials were categorized as broad-spectrum or non-broad-spectrum based on their ability to target a wide range of pathogens, including potentially multidrug-resistant organisms, methicillin-resistant *Staphylococcus aureus*, and *Pseudomonas aeruginosa*.

| **Antimicrobial** | **Broad-spectrum versus non-broad-spectrum** | **Drug class** |
| --- | --- | --- |
| Albendazole | Non-broad-spectrum | Anthelmintic |
| Amikacin | Broad-spectrum | Aminoglycoside |
| Amoxicillin | Non-broad-spectrum | Aminopenicillin |
| Amoxicillin-clavulanate | Broad-spectrum | β-lactam and β-lactamase inhibitor |
| Amphotericin B liposomal | Broad-spectrum | Polyene antifungal |
| Ampicillin | Non-broad-spectrum | Aminopenicillin |
| Ampicillin-sulbactam | Broad-spectrum | β-lactam and β-lactamase inhibitor |
| Artemether-lumefantrine | Non-broad-spectrum | Anti-malarial |
| Artesunate | Non-broad-spectrum | Anti-malarial |
| Atovaquone | Non-broad-spectrum | Antiprotozoal |
| Atovaquone-proguanil | Non-broad-spectrum | Antiprotozoal |
| Azithromycin | Non-broad-spectrum | Macrolide |
| Aztreonam | Broad-spectrum | Monobactam |
| Caspofungin | Non-broad-spectrum | Echinocandin antifungal |
| Cefadroxil | Non-broad-spectrum | Cephalosporin, 1^st^ generation |
| Cefazolin | Non-broad-spectrum | Cephalosporin, 1^st^ generation |
| Cefdinir | Non-broad-spectrum | Cephalosporin, 3^rd^ generation |
| Cefepime | Broad-spectrum | Cephalosporin, 4^th^ generation |
| Cefiderocol | Broad-spectrum | Cephalosporin, siderophore |
| Cefoxitin | Broad-spectrum | Cephalosporin, 2^nd^ generation |
| Cefpodoxime | Non-broad-spectrum | Cephalosporin, 3^rd^ generation |
| Cefprozil | Non-broad-spectrum | Cephalosporin, 2^nd^ generation |
| Ceftaroline | Broad-spectrum | Cephalosporin, 5^th^ generation |
| Ceftazidime | Broad-spectrum | Cephalosporin, 3^rd^ generation |
| Ceftazidime-avibactam | Broad-spectrum | Cephalosporin and β-lactamase inhibitor |
| Ceftolazone-tazobactam | Broad-spectrum | Cephalosporin and β-lactamase inhibitor |
| Ceftriaxone | Non-broad-spectrum | Cephalosporin, 3^rd^ generation |
| Cefuroxime | Non-broad-spectrum | Cephalosporin, 2^nd^ generation |
| Cephalexin | Non-broad-spectrum | Cephalosporin, 1^st^ generation |
| Ciprofloxacin | Broad-spectrum | Fluoroquinolone |
| Clarithromycin | Non-broad-spectrum | Macrolide |
| Clindamycin | Non-broad-spectrum | Lincosamide |
| Clofazimine | Non-broad-spectrum | Antimycobacterial |
| Dalbavancin | Broad-spectrum | Lipoglycopeptide |
| Dapsone | Non-broad-spectrum | Sulfone |
| Daptomycin | Broad-spectrum | Lipopeptide |
| Dicloxacillin | Non-broad-spectrum | β-lactam |
| Doxycycline | Non-broad-spectrum | Tetracycline |
| Eravacycline | Broad-spectrum | Fluorocycline |
| Ertapenem | Non-broad-spectrum | Carbapenem |
| Erythromycin | Non-broad-spectrum | Macrolide |
| Fluconazole | Non-broad-spectrum | Triazole antifungal |
| Flucytosine | Non-broad-spectrum | Pyrimidine antifungal |
| Fosfomycin | Broad-spectrum | Phosphonic acid derivative |
| Gentamicin | Broad-spectrum | Aminoglycoside |
| Imipenem-cilastatin | Broad-spectrum | Carbapenem |
| Imipenem-cilastatin-relebactam | Broad-spectrum | Carbapenem and β-lactamase inhibitor |
| Isavuconazonium | Broad-spectrum | Triazole antifungal |
| Isoniazid | Non-broad-spectrum | Antimycobacterial |
| Itraconazole | Non-broad-spectrum | Triazole antifungal |
| Ivermectin | Non-broad-spectrum | Anti-parasite |
| Ketoconazole | Non-broad-spectrum | Imidazole antifungal |
| Levofloxacin | Broad-spectrum | Fluoroquinolone |
| Linezolid | Broad-spectrum | Oxazolidinone |
| Meropenem | Broad-spectrum | Carbapenem |
| Meropenem-vaborbactam | Broad-spectrum | Carbapenem and β-lactamase inhibitor |
| Metronidazole | Non-broad-spectrum | Nitroimidazole |
| Micafungin | Non-broad-spectrum | Echinocandin antifungal |
| Minocycline | Non-broad-spectrum | Tetracycline |
| Moxifloxacin | Broad-spectrum | Fluoroquinolone |
| Nafcillin | Non-broad-spectrum | β-lactam, penicillinase-resistant |
| Neomycin | Non-broad-spectrum | Aminoglycoside |
| Nitazoxanide | Non-broad-spectrum | Antiprotozoal |
| Nitrofurantoin | Non-broad-spectrum | Nitrofuran |
| Nystatin | Non-broad-spectrum | Polyene antifungal |
| Omadacycline | Broad-spectrum | Tetracycline |
| Oxacillin | Non-broad-spectrum | β-lactam, penicillinase-resistant |
| Penicillin G | Non-broad-spectrum | Penicillin |
| Penicillin V | Non-broad-spectrum | Penicillin |
| Piperacillin-tazobactam | Broad-spectrum | β-lactam and β-lactamase inhibitor |
| Posaconazole | Broad-spectrum | Triazole antifungal |
| Praziquantel | Non-broad-spectrum | Anthelmintic |
| Pretomanid | Non-broad-spectrum | Antimycobacterial |
| Primaquine | Non-broad-spectrum | Antimalarial |
| Pyrazinamide | Non-broad-spectrum | Antimycobacterial |
| Rifabutin | Non-broad-spectrum | Antimycobacterial |
| Rifampin | Non-broad-spectrum | Antimycobacterial |
| Rifapentine | Non-broad-spectrum | Antimycobacterial |
| Rifaximin | Non-broad-spectrum | Rifamycin derivative |
| Sulbactam-durlobactam | Broad-spectrum | β-lactam and β-lactamase inhibitor |
| Sulfadiazine | Non-broad-spectrum | Sulfonamide |
| Sulfamethoxazole-trimethoprim | Broad-spectrum | Sulfonamide and anti-folate |
| Tafenoquine | Non-broad-spectrum | Antimalarial |
| Tedizolid | Broad-spectrum | Oxazolidinone |
| Terbinafine | Non-broad-spectrum | Allylamine antifungal |
| Tetracycline | Non-broad-spectrum | Tetracycline |
| Tinidazole | Non-broad-spectrum | Nitroimidazole |
| Tobramycin | Broad-spectrum | Aminoglycoside |
| Trimethoprim | Non-broad-spectrum | Anti-folate |
| Vancomycin | Broad-spectrum | Glycopeptide |
| Voriconazole | Broad-spectrum | Triazole antifungal |

### **eFigures.**

#### **eFigure 1.** Changes in Other Blood Culture Outcomes at All 3 Hospital Sites Combined.

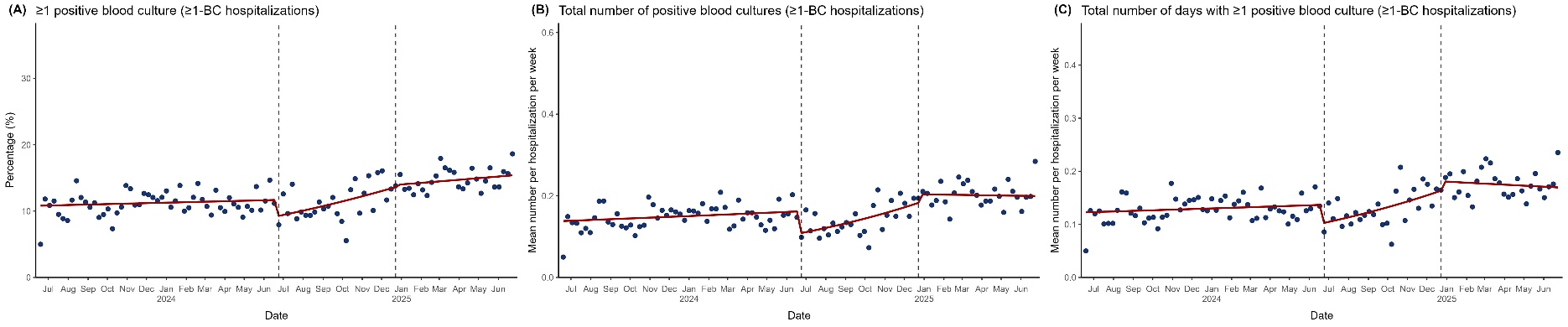

**eFigure 1.** Changes in Other Secondary Blood Culture Outcomes at All 3 Hospital Sites Combined. Blue dots represent weekly averages and the red solid line represents the fitted line from the segmented regression model assessing for abrupt level and slope changes at the start and stop dates of the blood culture restriction of 1 blood culture set per patient per 24 hours, which were June 26, 2024 and December 23, 2024, respectively, represented by dashed vertical lines. “≥1-BC hospitalizations” designates the hospitalizations with ≥1 blood culture obtained subgroup of all hospitalizations. The figure displays the following:

(A) Weekly percentage of hospitalizations with ≥1 positive blood culture for ≥1-BC hospitalizations

(B) Mean number of positive blood cultures obtained for ≥1-BC hospitalizations

(C) Mean number of days with ≥1 positive blood culture for ≥1-BC hospitalizations

#### **eFigure 2.** Changes in Other Antimicrobial Outcomes at All 3 Hospital Sites Combined.

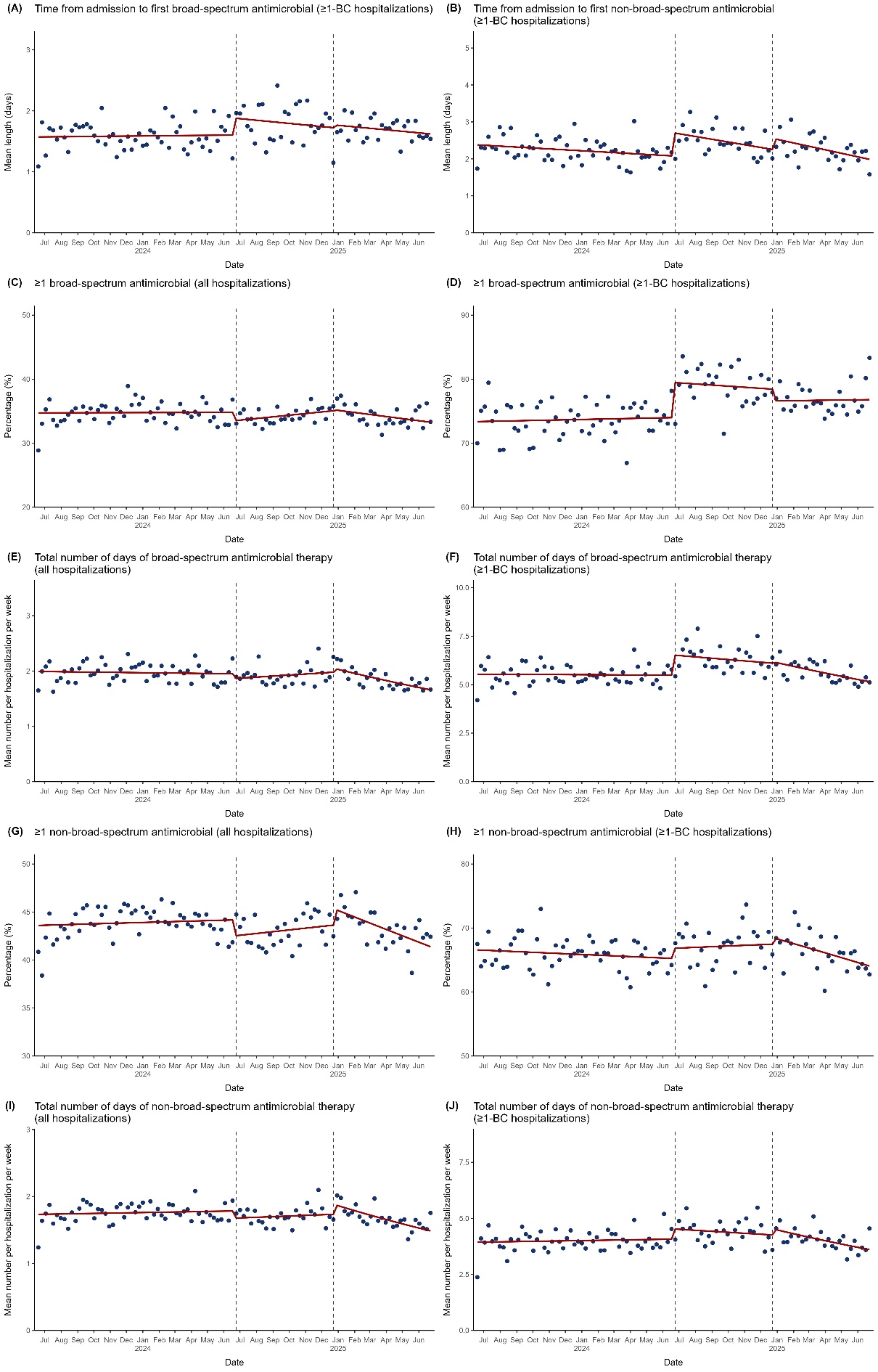

**eFigure 2.** Changes in Other Antimicrobial Outcomes at All 3 Hospital Sites Combined. Blue dots represent weekly averages and the red solid line represents the fitted line from the segmented regression model assessing for abrupt level and slope changes at the start and stop dates of the blood culture restriction of 1 blood culture set per patient per 24 hours, which were June 26, 2024 and December 23, 2024, respectively, represented by dashed vertical lines. “≥1-BC hospitalizations” designates the hospitalizations with ≥1 blood culture obtained subgroup of all hospitalizations. The figure displays the following:
(A) Mean length of time from admission to the first broad-spectrum antimicrobial administered for ≥1-BC hospitalizations
(B) Mean length of time from admission to the first non-broad-spectrum antimicrobial administered for ≥1-BC hospitalizations
(C) Percentage of hospitalizations with ≥1 broad-spectrum antimicrobial administered for all hospitalizations
(D) Percentage of hospitalizations with ≥1 broad-spectrum antimicrobial administered for ≥1-BC hospitalizations
(E) Mean number of days of antimicrobial therapy for all hospitalizations
(F) Mean number of days of antimicrobial therapy for ≥1-BC hospitalizations
(G) Percentage of hospitalizations with ≥1 non-broad-spectrum antimicrobial administered for all hospitalizations
(H) Percentage of hospitalizations with ≥1 non-broad-spectrum antimicrobial administered for ≥1-BC hospitalizations
(I) Mean number of days of non-broad-spectrum antimicrobial therapy for all hospitalizations
(J) Mean number of days of non-broad-spectrum antimicrobial therapy for ≥1-BC hospitalizations

#### **eFigure 3.** Changes in Primary Outcomes at the Hospital of the University of Pennsylvania.

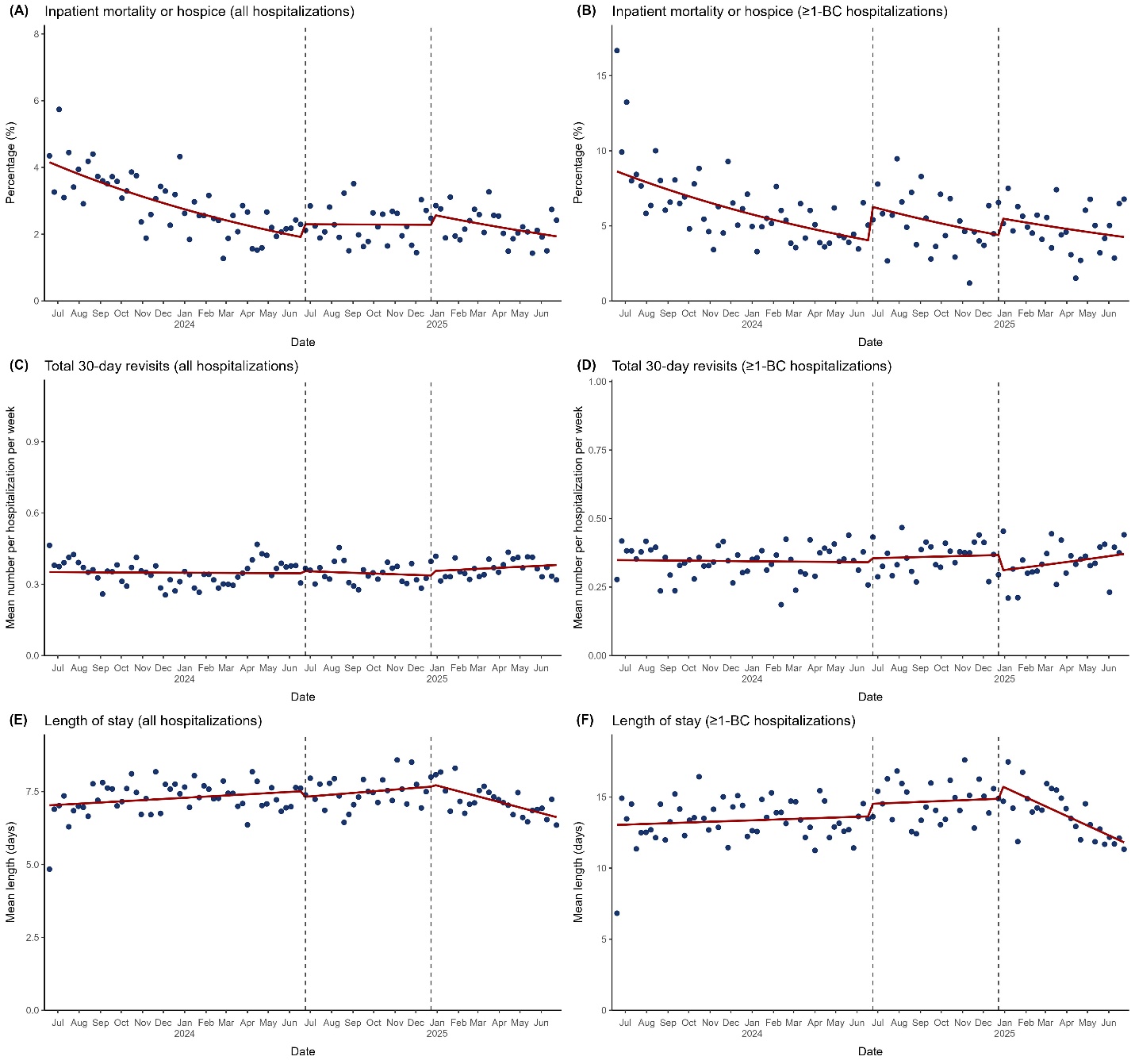

**eFigure 3.** Changes in Primary Outcomes at the Hospital of the University of Pennsylvania. Blue dots represent weekly averages and the red solid line represents the fitted line from the segmented regression model assessing for abrupt level and slope changes at the start and stop dates of the blood culture restriction of 1 blood culture set per patient per 24 hours, which were June 26, 2024 and December 23, 2024, respectively, represented by dashed vertical lines. “≥1-BC hospitalizations” designates the hospitalizations with ≥1 blood culture obtained subgroup of all hospitalizations. The figure displays the following:

(A) Percentage of inpatient mortality or hospice discharge for all hospitalizations

(B) Percentage of inpatient mortality or hospice discharge for ≥1-BC hospitalizations

(C) Mean number of total 30-day revisits for all hospitalizations

(D) Mean number of total 30-day revisits for ≥1-BC hospitalizations

(E) Mean length of stay for all hospitalizations

(F) Mean length of stay for ≥1-BC hospitalizations

#### **eFigure 4.** Changes in Blood Culture Outcomes at the Hospital of the University of Pennsylvania.

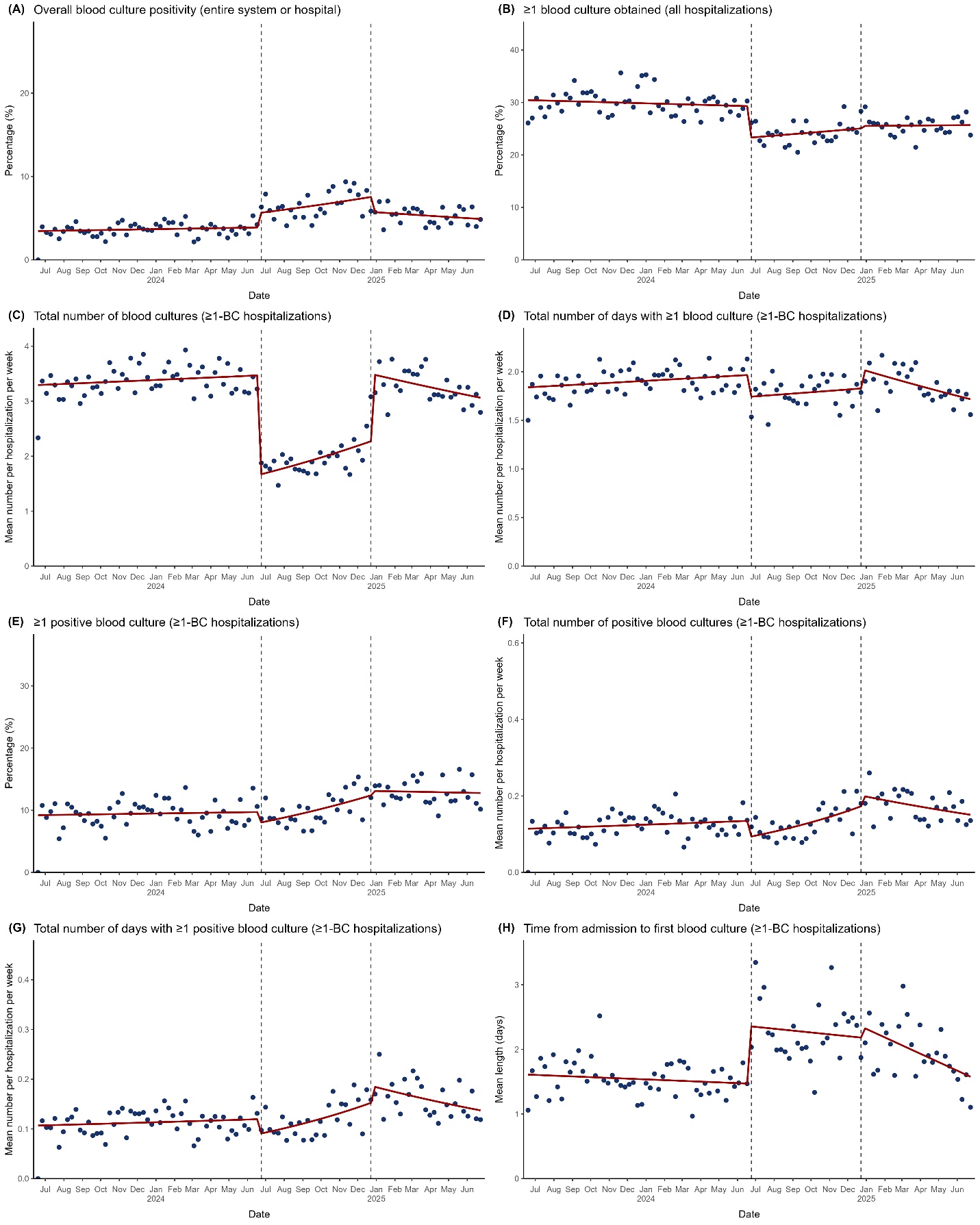

**eFigure 4.** Changes in Blood Culture Outcomes at the Hospital of the University of Pennsylvania. Blue dots represent weekly averages and the red solid line represents the fitted line from the segmented regression model assessing for abrupt level and slope changes at the start and stop dates of the blood culture restriction of 1 blood culture set per patient per 24 hours, which were June 26, 2024 and December 23, 2024, respectively, represented by dashed vertical lines. “≥1-BC hospitalizations” designates the hospitalizations with ≥1 blood culture obtained subgroup of all hospitalizations. The figure displays the following:
(A) Percentage of overall blood culture positivity across the entire hospital
(B) Percentage of hospitalizations with ≥1 blood culture obtained for all hospitalizations
(C) Mean number of blood cultures obtained for ≥1-BC hospitalizations
(D) Mean number of days with ≥1 blood culture obtained for ≥1-BC hospitalizations
(E) Percentage of hospitalizations with ≥1 positive blood culture for ≥1-BC hospitalizations
(F) Mean number of positive blood cultures obtained for ≥1-BC hospitalizations
(G) Mean number of days with ≥1 positive blood culture for ≥1-BC hospitalizations
(H) Mean length of time from admission to the first blood culture obtained for ≥1-BC hospitalizations

#### **eFigure 5.** Changes in Antimicrobial Outcomes at the Hospital of the University of Pennsylvania.

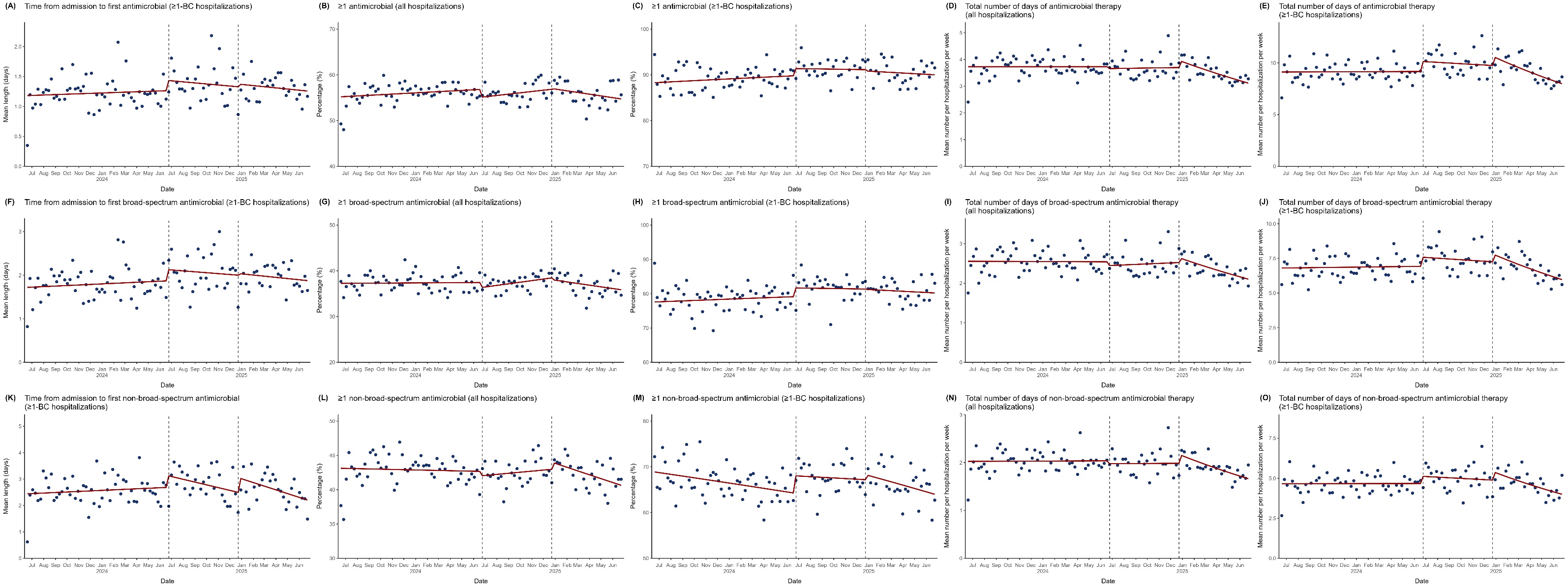

**eFigure 5.** Changes in Antimicrobial Outcomes at the Hospital of the University of Pennsylvania. Blue dots represent weekly averages and the red solid line represents the fitted line from the segmented regression model assessing for abrupt level and slope changes at the start and stop dates of the blood culture restriction of 1 blood culture set per patient per 24 hours, which were June 26, 2024 and December 23, 2024, respectively, represented by dashed vertical lines. “≥1-BC hospitalizations” designates the hospitalizations with ≥1 blood culture obtained subgroup of all hospitalizations. The figure displays the following:
(A) Mean length of time from admission to the first antimicrobial administered for ≥1-BC hospitalizations
(B) Weekly percentage of hospitalizations with ≥1 antimicrobial administered for all hospitalizations
(C) Weekly percentage of hospitalizations with ≥1 antimicrobial administered for ≥1-BC hospitalizations
(D) Mean number of days of antimicrobial therapy for all hospitalizations
(E) Mean number of days of antimicrobial therapy for ≥1-BC hospitalizations
(F) Mean length of time from admission to the first broad-spectrum antimicrobial administered for ≥1-BC hospitalizations
(G) Weekly percentage of hospitalizations with ≥1 broad-spectrum antimicrobial administered for all hospitalizations
(H) Weekly percentage of hospitalizations with ≥1 broad-spectrum antimicrobial administered for ≥1-BC hospitalizations
(I) Mean number of days of broad-spectrum antimicrobial therapy for all hospitalizations
(J) Mean number of days of broad-spectrum antimicrobial therapy for ≥1-BC hospitalizations
(K) Mean length of time from admission to the first non-broad-spectrum antimicrobial administered for ≥1-BC hospitalizations
(L) Weekly percentage of hospitalizations with ≥1 non-broad-spectrum antimicrobial administered for all hospitalizations
(M) Weekly percentage of hospitalizations with ≥1 non-broad-spectrum antimicrobial administered for ≥1-BC hospitalizations
(N) Mean number of days of non-broad-spectrum antimicrobial therapy for all hospitalizations
(O) Mean number of days of non-broad-spectrum antimicrobial therapy for ≥1-BC hospitalizations

#### **eFigure 6.** Changes in Primary Outcomes at Penn Presbyterian Medical Center.

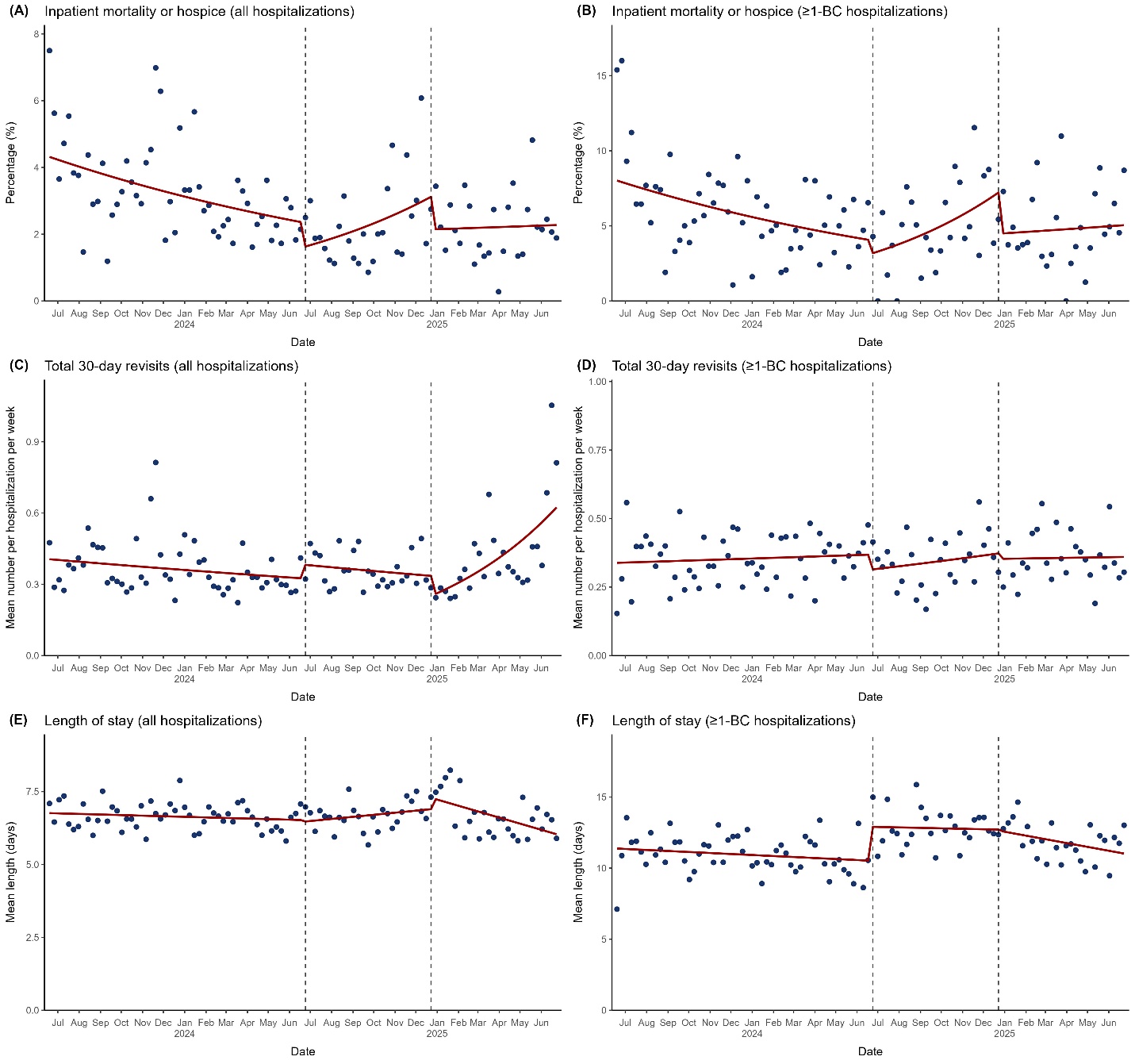

**eFigure 6.** Changes in Primary Outcomes at Penn Presbyterian Medical Center. Blue dots represent weekly averages and the red solid line represents the fitted line from the segmented regression model assessing for abrupt level and slope changes at the start and stop dates of the blood culture restriction of 1 blood culture set per patient per 24 hours, which were June 26, 2024 and December 23, 2024, respectively, represented by dashed vertical lines. “≥1-BC hospitalizations” designates the hospitalizations with ≥1 blood culture obtained subgroup of all hospitalizations. The figure displays the following:

(A) Percentage of inpatient mortality or hospice discharge for all hospitalizations

(B) Percentage of inpatient mortality or hospice discharge for ≥1-BC hospitalizations

(C) Mean number of total 30-day revisits for all hospitalizations

(D) Mean number of total 30-day revisits for ≥1-BC hospitalizations

(E) Mean length of stay for all hospitalizations

(F) Mean length of stay for ≥1-BC hospitalizations

#### **eFigure 7.** Changes in Blood Culture Outcomes at Penn Presbyterian Medical Center.

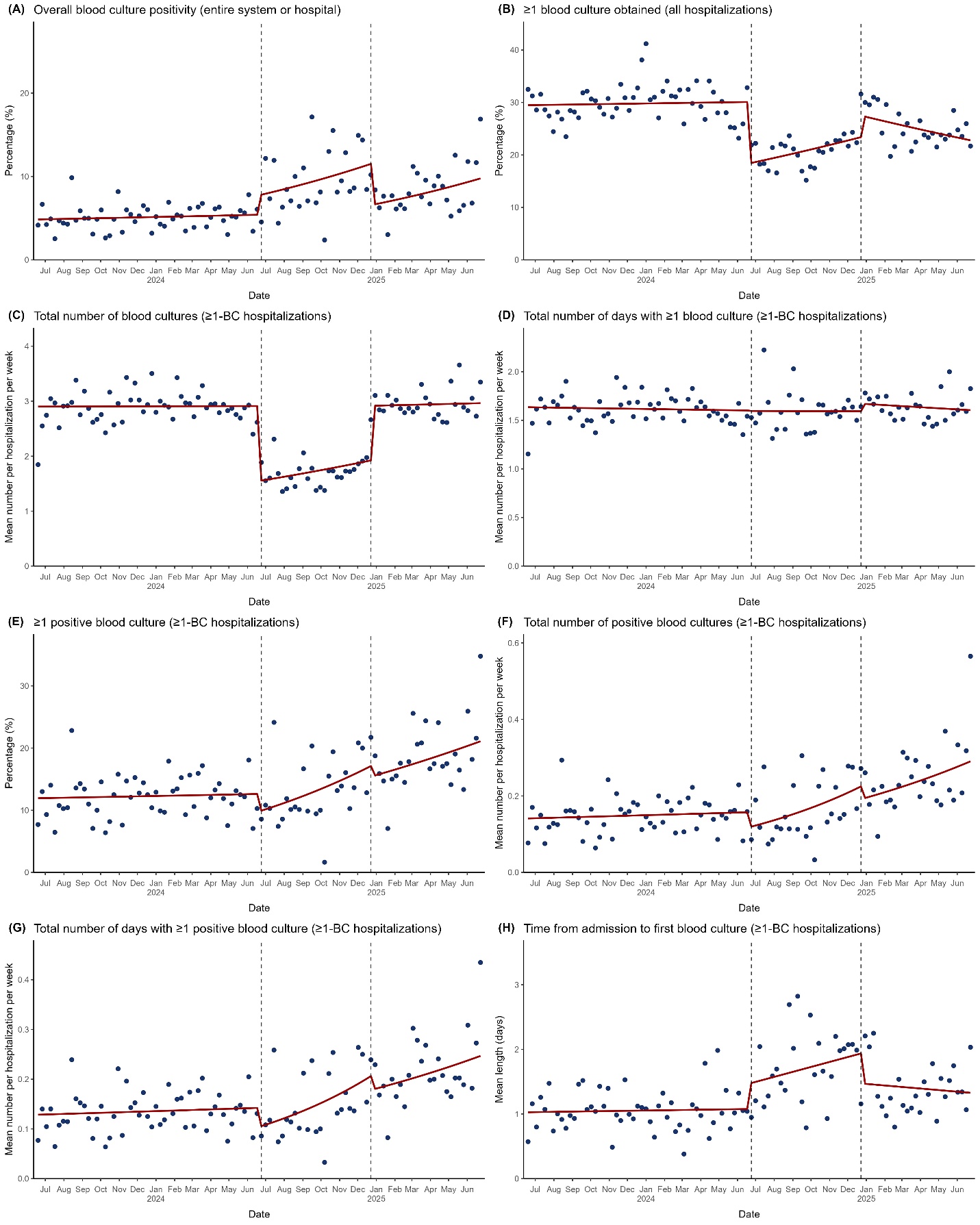

**eFigure 7.** Changes in Blood Culture Outcomes at Penn Presbyterian Medical Center. Blue dots represent weekly averages and the red solid line represents the fitted line from the segmented regression model assessing for abrupt level and slope changes at the start and stop dates of the blood culture restriction of 1 blood culture set per patient per 24 hours, which were June 26, 2024 and December 23, 2024, respectively, represented by dashed vertical lines. “≥1-BC hospitalizations” designates the hospitalizations with ≥1 blood culture obtained subgroup of all hospitalizations. The figure displays the following:
(A) Percentage of overall blood culture positivity across the entire hospital
(B) Percentage of hospitalizations with ≥1 blood culture obtained for all hospitalizations
(C) Mean number of blood cultures obtained for ≥1-BC hospitalizations
(D) Mean number of days with ≥1 blood culture obtained for ≥1-BC hospitalizations
(E) Percentage of hospitalizations with ≥1 positive blood culture for ≥1-BC hospitalizations
(F) Mean number of positive blood cultures obtained for ≥1-BC hospitalizations
(G) Mean number of days with ≥1 positive blood culture for ≥1-BC hospitalizations
(H) Mean length of time from admission to the first blood culture obtained for ≥1-BC hospitalizations

#### **eFigure 8.** Changes in Antimicrobial Outcomes at Penn Presbyterian Medical Center.

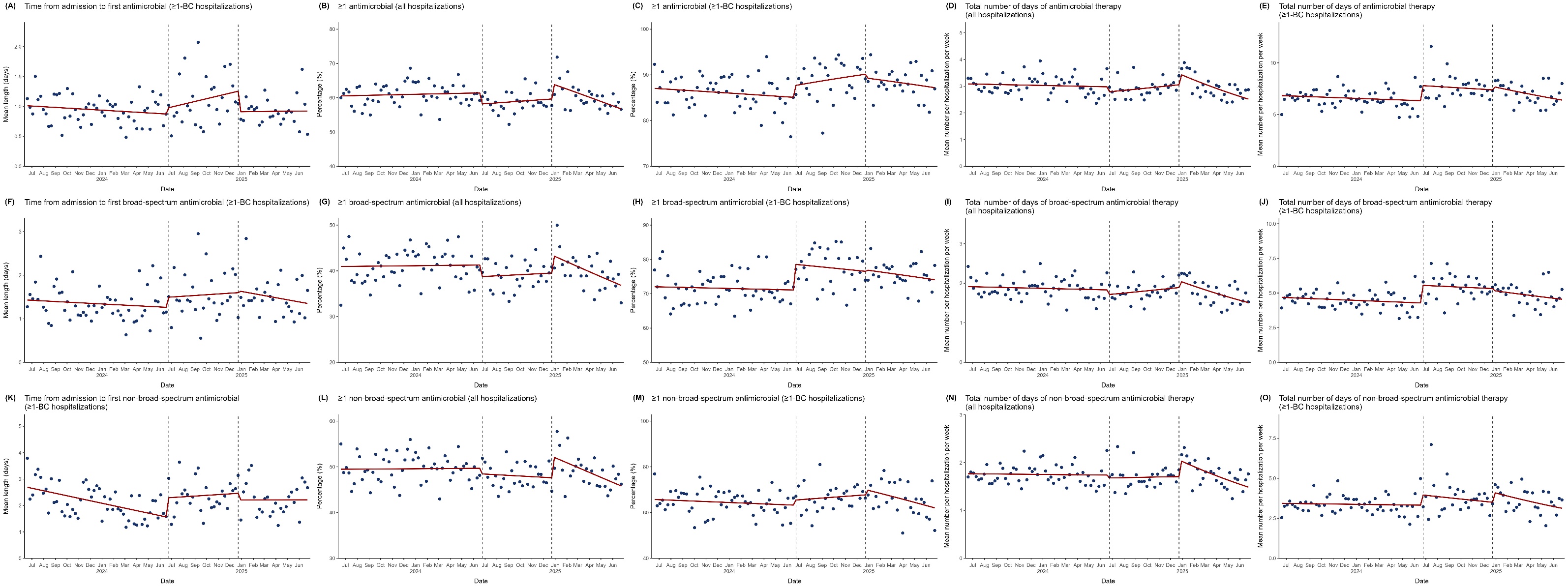

**eFigure 8.** Changes in Antimicrobial Outcomes at Penn Presbyterian Medical Center. Blue dots represent weekly averages and the red solid line represents the fitted line from the segmented regression model assessing for abrupt level and slope changes at the start and stop dates of the blood culture restriction of 1 blood culture set per patient per 24 hours, which were June 26, 2024 and December 23, 2024, respectively, represented by dashed vertical lines. “≥1-BC hospitalizations” designates the hospitalizations with ≥1 blood culture obtained subgroup of all hospitalizations. The figure displays the following:
(A) Mean length of time from admission to the first antimicrobial administered for ≥1-BC hospitalizations
(B) Weekly percentage of hospitalizations with ≥1 antimicrobial administered for all hospitalizations
(C) Weekly percentage of hospitalizations with ≥1 antimicrobial administered for ≥1-BC hospitalizations
(D) Mean number of days of antimicrobial therapy for all hospitalizations
(E) Mean number of days of antimicrobial therapy for ≥1-BC hospitalizations
(F) Mean length of time from admission to the first broad-spectrum antimicrobial administered for ≥1-BC hospitalizations
(G) Weekly percentage of hospitalizations with ≥1 broad-spectrum antimicrobial administered for all hospitalizations
(H) Weekly percentage of hospitalizations with ≥1 broad-spectrum antimicrobial administered for ≥1-BC hospitalizations
(I) Mean number of days of broad-spectrum antimicrobial therapy for all hospitalizations
(J) Mean number of days of broad-spectrum antimicrobial therapy for ≥1-BC hospitalizations
(K) Mean length of time from admission to the first non-broad-spectrum antimicrobial administered for ≥1-BC hospitalizations
(L) Weekly percentage of hospitalizations with ≥1 non-broad-spectrum antimicrobial administered for all hospitalizations
(M) Weekly percentage of hospitalizations with ≥1 non-broad-spectrum antimicrobial administered for ≥1-BC hospitalizations
(N) Mean number of days of non-broad-spectrum antimicrobial therapy for all hospitalizations
(O) Mean number of days of non-broad-spectrum antimicrobial therapy for ≥1-BC hospitalizations

#### **eFigure 9.** Changes in Primary Outcomes at Pennsylvania Hospital.

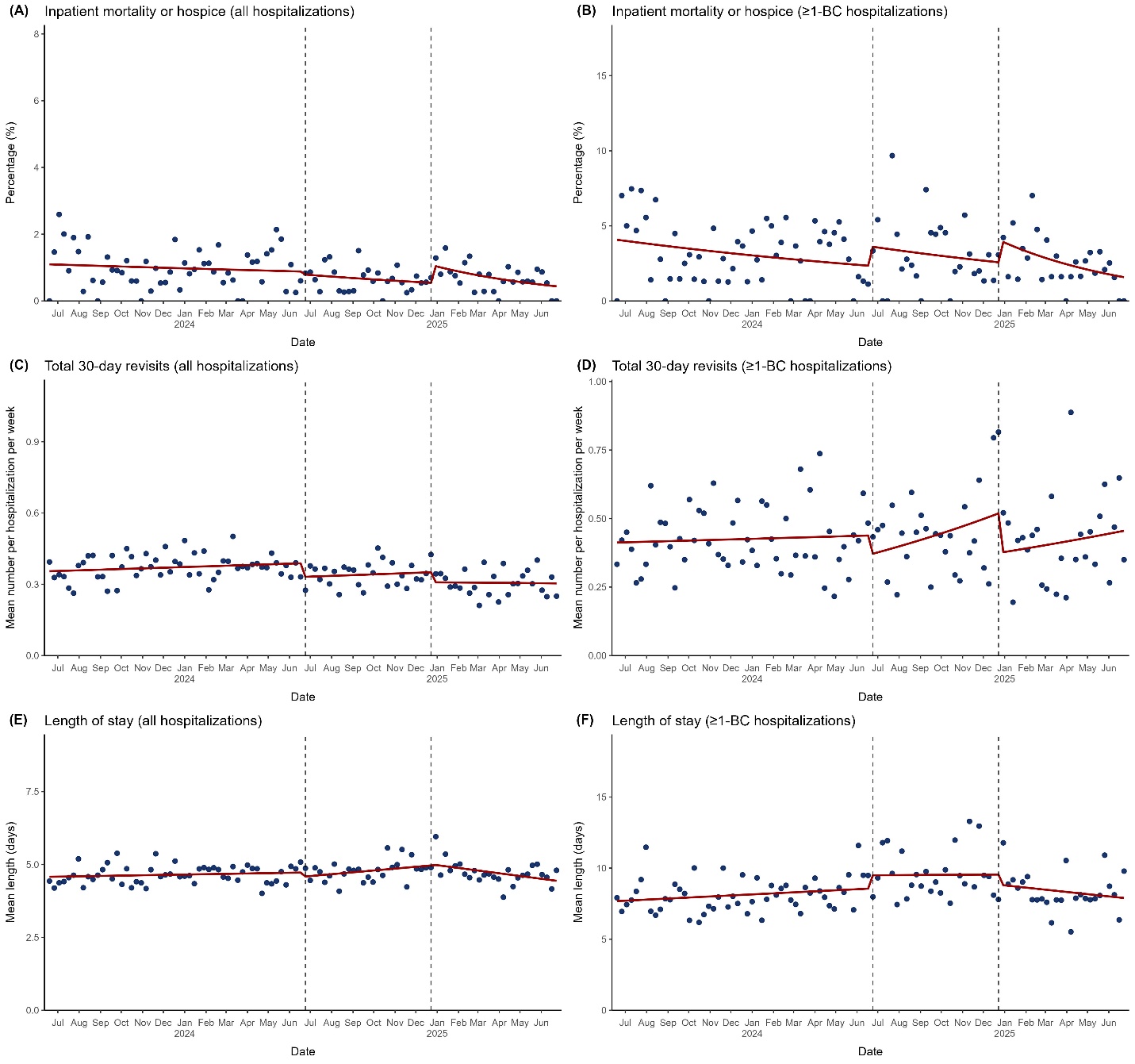

**eFigure 9.** Changes in Primary Outcomes at Pennsylvania Hospital. Blue dots represent weekly averages and the red solid line represents the fitted line from the segmented regression model assessing for abrupt level and slope changes at the start and stop dates of the blood culture restriction of 1 blood culture set per patient per 24 hours, which were June 26, 2024 and December 23, 2024, respectively, represented by dashed vertical lines. “≥1-BC hospitalizations” designates the hospitalizations with ≥1 blood culture obtained subgroup of all hospitalizations. The figure displays the following:

(A) Percentage of inpatient mortality or hospice discharge for all hospitalizations

(B) Percentage of inpatient mortality or hospice discharge for ≥1-BC hospitalizations

(C) Mean number of total 30-day revisits for all hospitalizations

(D) Mean number of total 30-day revisits for ≥1-BC hospitalizations

(E) Mean length of stay for all hospitalizations

(F) Mean length of stay for ≥1-BC hospitalizations

#### **eFigure 10.** Changes in Blood Culture Outcomes at Pennsylvania Hospital.

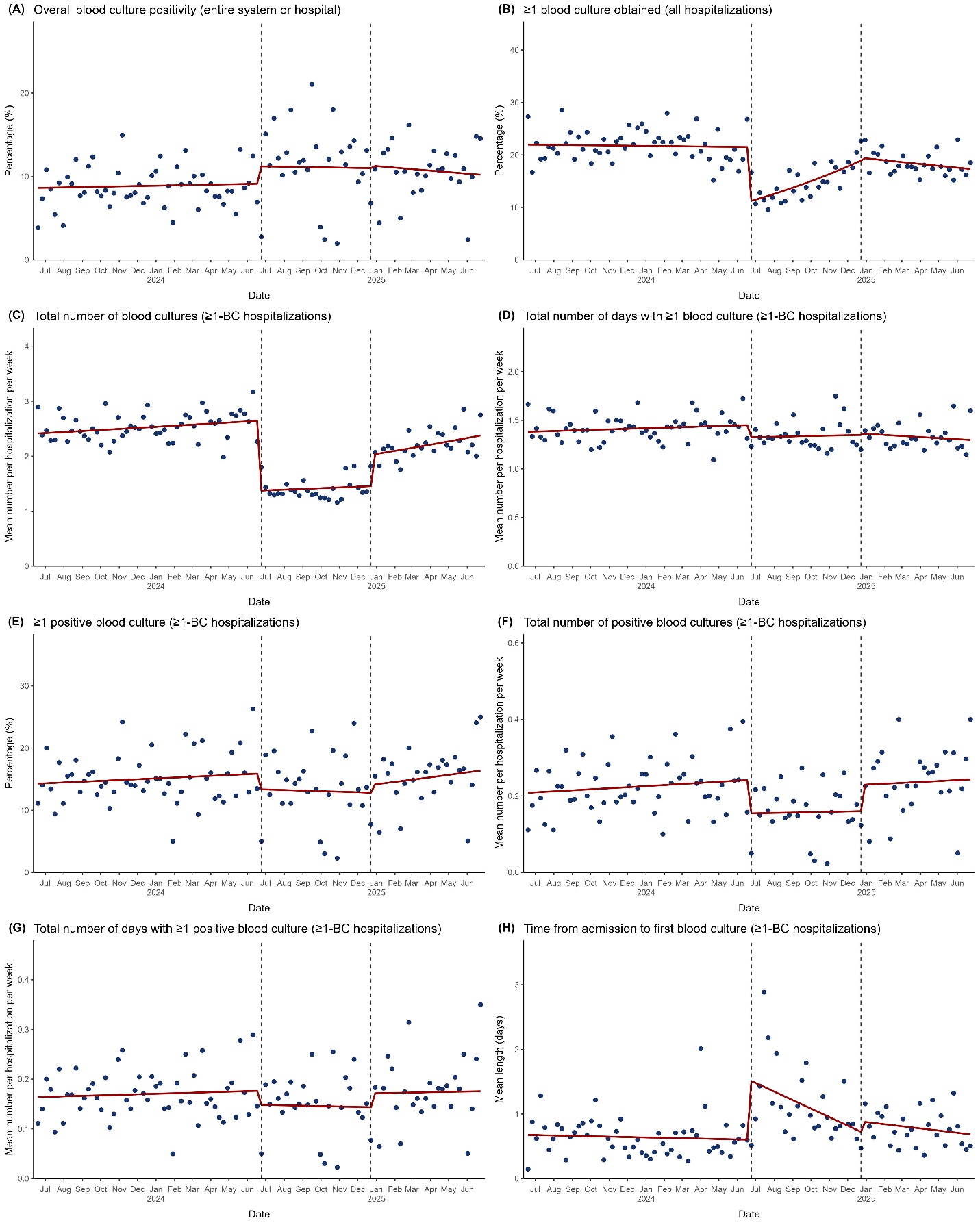

**eFigure 10.** Changes in Blood Culture Outcomes at Pennsylvania Hospital. Blue dots represent weekly averages and the red solid line represents the fitted line from the segmented regression model assessing for abrupt level and slope changes at the start and stop dates of the blood culture restriction of 1 blood culture set per patient per 24 hours, which were June 26, 2024 and December 23, 2024, respectively, represented by dashed vertical lines. “≥1-BC hospitalizations” designates the hospitalizations with ≥1 blood culture obtained subgroup of all hospitalizations. The figure displays the following:
(A) Percentage of overall blood culture positivity across the entire hospital
(B) Percentage of hospitalizations with ≥1 blood culture obtained for all hospitalizations
(C) Mean number of blood cultures obtained for ≥1-BC hospitalizations
(D) Mean number of days with ≥1 blood culture obtained for ≥1-BC hospitalizations
(E) Percentage of hospitalizations with ≥1 positive blood culture for ≥1-BC hospitalizations
(F) Mean number of positive blood cultures obtained for ≥1-BC hospitalizations
(G) Mean number of days with ≥1 positive blood culture for ≥1-BC hospitalizations
(H) Mean length of time from admission to the first blood culture obtained for ≥1-BC hospitalizations

#### **eFigure 11.** Changes in Antimicrobial Outcomes at Pennsylvania Hospital.

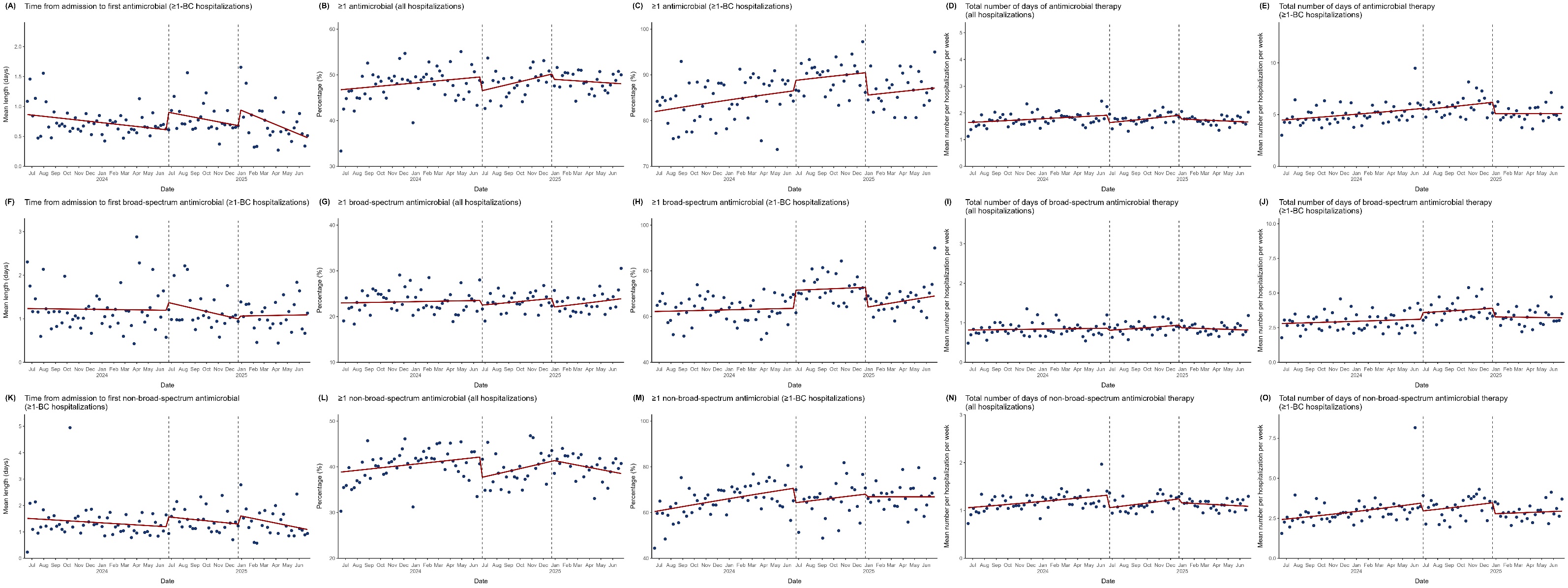

**eFigure 11.** Changes in Antimicrobial Outcomes at Pennsylvania Hospital. Blue dots represent weekly averages and the red solid line represents the fitted line from the segmented regression model assessing for abrupt level and slope changes at the start and stop dates of the blood culture restriction of 1 blood culture set per patient per 24 hours, which were June 26, 2024 and December 23, 2024, respectively, represented by dashed vertical lines. “≥1-BC hospitalizations” designates the hospitalizations with ≥1 blood culture obtained subgroup of all hospitalizations. The figure displays the following:
(A) Mean length of time from admission to the first antimicrobial administered for ≥1-BC hospitalizations
(B) Weekly percentage of hospitalizations with ≥1 antimicrobial administered for all hospitalizations
(C) Weekly percentage of hospitalizations with ≥1 antimicrobial administered for ≥1-BC hospitalizations
(D) Mean number of days of antimicrobial therapy for all hospitalizations
(E) Mean number of days of antimicrobial therapy for ≥1-BC hospitalizations
(F) Mean length of time from admission to the first broad-spectrum antimicrobial administered for ≥1-BC hospitalizations
(G) Weekly percentage of hospitalizations with ≥1 broad-spectrum antimicrobial administered for all hospitalizations
(H) Weekly percentage of hospitalizations with ≥1 broad-spectrum antimicrobial administered for ≥1-BC hospitalizations
(I) Mean number of days of broad-spectrum antimicrobial therapy for all hospitalizations
(J) Mean number of days of broad-spectrum antimicrobial therapy for ≥1-BC hospitalizations
(K) Mean length of time from admission to the first non-broad-spectrum antimicrobial administered for ≥1-BC hospitalizations
(L) Weekly percentage of hospitalizations with ≥1 non-broad-spectrum antimicrobial administered for all hospitalizations
(M) Weekly percentage of hospitalizations with ≥1 non-broad-spectrum antimicrobial administered for ≥1-BC hospitalizations
(N) Mean number of days of non-broad-spectrum antimicrobial therapy for all hospitalizations
(O) Mean number of days of non-broad-spectrum antimicrobial therapy for ≥1-BC hospitalizations

#### **eFigure 12.** Excess In-Hospital Mortality or Hospice Discharge at All 3 Hospital Sites Combined.

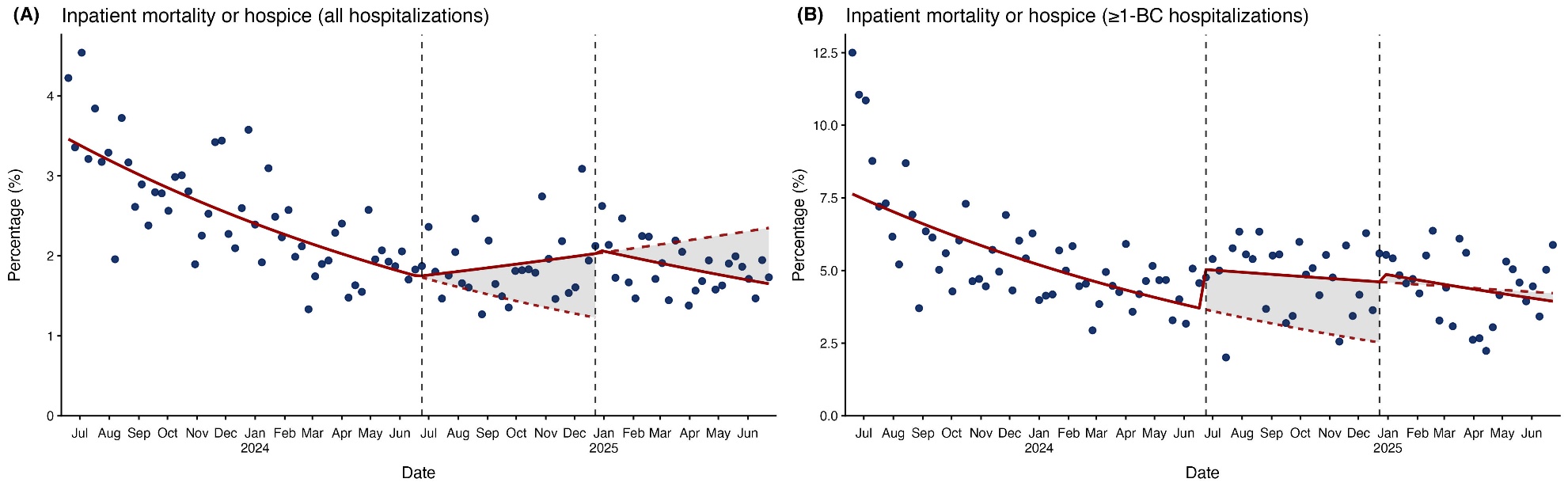

**eFigure 12.** Excess In-Hospital Mortality or Hospice Discharge at All 3 Hospital Sites Combined. Blue dots represent weekly averages and the red solid line represents the fitted line from the segmented regression model assessing for abrupt level and slope changes at the start and stop dates of the blood culture restriction of 1 blood culture set per patient per 24 hours, which were June 26, 2024 and December 23, 2024, respectively (represented by dashed vertical lines). The figure displays the weekly percentage of inpatient mortality or hospice discharge for all hospitalizations (A) and hospitalizations with ≥1 blood culture obtained (B). The fitted solid red lines represent the models for the pre-restriction, restriction, and post-restriction periods, with the dashed red lines representing the pre-restriction versus restriction and restriction versus post-restriction counterfactuals and the gray shaded areas representing the excess in in-hospital mortality or hospice discharge. Excess deaths or hospice discharges in the restriction period when extrapolating pre-restriction trends were 159 (4.4 per 1000 hospitalizations; 95% CI, 2.0–6.7, P=.002) and 141 (18.7 per 1000 hospitalizations; 95% CI, 10.1–26.2, P<.001) in hospitalizations and hospitalizations with ≥1 blood culture obtained, respectively. Excess deaths or hospice discharges in the post-restriction period when extrapolating restriction period trends were 122 (3.2 per hospitalizations; 95% CI –1.9 to 9.6, P=.23) and 1 (0.1 per 1000 hospitalizations; 95% CI –8.6 to 9.8, P=.97) in all hospitalizations and hospitalizations with ≥1 blood culture obtained, respectively, though these estimates were not statistically significant.

#### **eFigure 13.** Changes in Primary Outcomes at All 3 Hospital Sites Combined, Excluding Hospitalizations Overlapping Restriction Cutoff Dates.

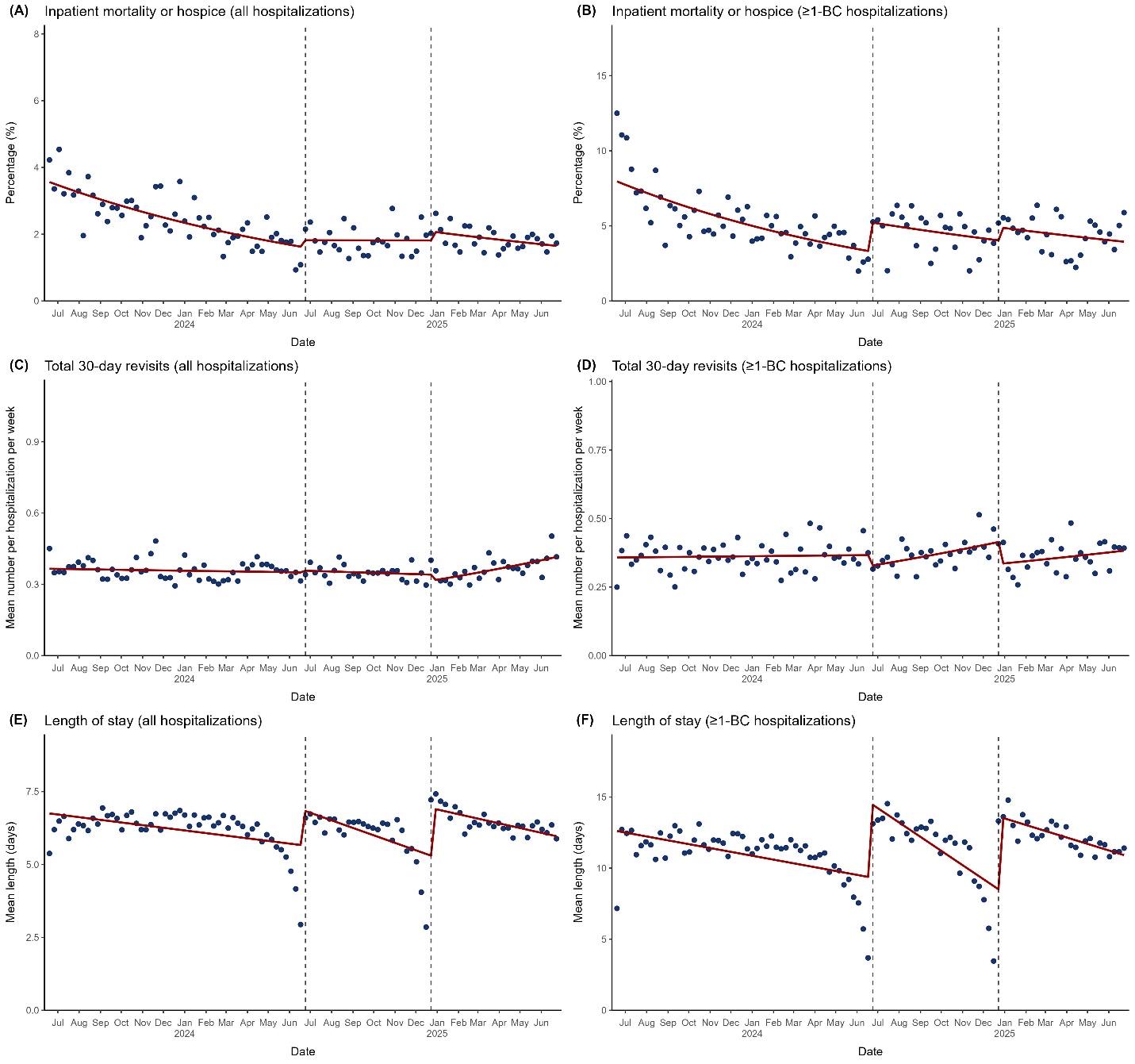

**eFigure 13**. Changes in Primary Outcomes at All 3 Hospital Sites Combined, Excluding Hospitalizations Overlapping Restriction Cutoff Dates. Blue dots represent weekly averages and the red solid line represents the fitted line from the segmented regression model assessing for abrupt level and slope changes at the start and stop dates of the blood culture restriction of 1 blood culture set per patient per 24 hours, which were June 26, 2024 and December 23, 2024, respectively, represented by dashed vertical lines. Any hospitalizations whose admission and discharge dates overlapped the cutoff dates were excluded as a sensitivity analysis. “≥1-BC hospitalizations” designates the hospitalizations with ≥1 blood culture obtained subgroup of all hospitalizations. The figure displays the following:

(A) Percentage of inpatient mortality or hospice discharge for all hospitalizations

(B) Percentage of inpatient mortality or hospice discharge for ≥1-BC hospitalizations

(C) Mean number of total 30-day revisits for all hospitalizations

(D) Mean number of total 30-day revisits for ≥1-BC hospitalizations

(E) Mean length of stay for all hospitalizations

(F) Mean length of stay for ≥1-BC hospitalizations (F) Mean length of stay for ≥1-BC hospitalizations

#### **eFigure 14.** Changes in Blood Culture Outcomes at All 3 Hospital Sites Combined, Excluding Hospitalizations Overlapping Restriction Cutoff Dates.

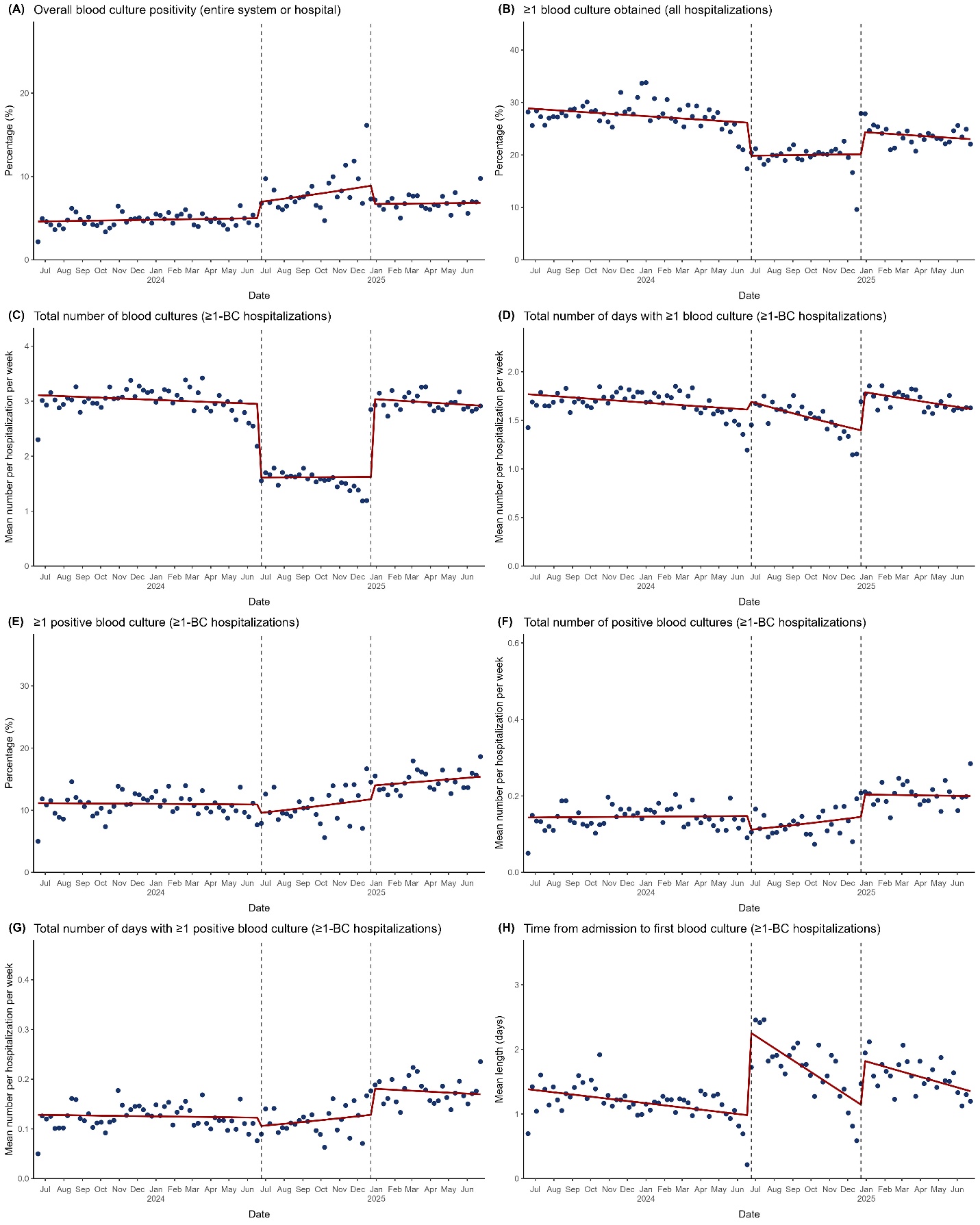

**eFigure 14.** Changes in Blood Culture Outcomes at All 3 Hospital Sites Combined, Excluding Hospitalizations Overlapping Restriction Cutoff Dates. Blue dots represent weekly averages and the red solid line represents the fitted line from the segmented regression model assessing for abrupt level and slope changes at the start and stop dates of the blood culture restriction of 1 blood culture set per patient per 24 hours, which were June 26, 2024 and December 23, 2024, respectively, represented by dashed vertical lines. Any hospitalizations whose admission and discharge dates overlapped the cutoff dates were excluded as a sensitivity analysis. “≥1-BC hospitalizations” designates the hospitalizations with ≥1 blood culture obtained subgroup of all hospitalizations. The figure displays the following:
(A) Percentage of overall blood culture positivity across all 3 hospitals combined
(B) Percentage of hospitalizations with ≥1 blood culture obtained for all hospitalizations
(C) Mean number of blood cultures obtained for ≥1-BC hospitalizations
(D) Mean number of days with ≥1 blood culture obtained for ≥1-BC hospitalizations
(E) Percentage of hospitalizations with ≥1 positive blood culture for ≥1-BC hospitalizations
(F) Mean number of positive blood cultures obtained for ≥1-BC hospitalizations
(G) Mean number of days with ≥1 positive blood culture for ≥1-BC hospitalizations
(H) Mean length of time from admission to the first blood culture obtained for ≥1-BC hospitalizations

#### **eFigure 15.** Changes in Antimicrobial Outcomes Over Time at All 3 Hospital Sites Combined, Excluding Hospitalizations Overlapping Restriction Cutoff Dates.

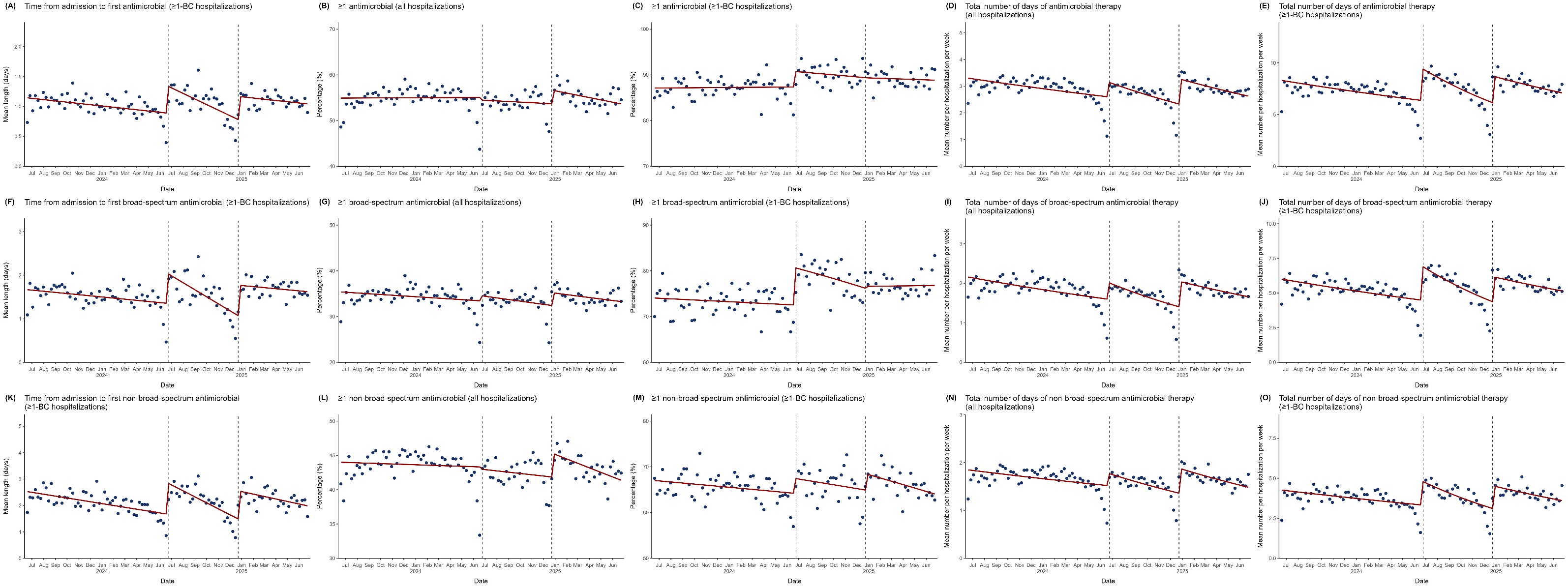

**eFigure 15.** Changes in Antimicrobial Outcomes Over Time at All 3 Hospital Sites Combined, Excluding Hospitalizations Overlapping Restriction Cutoff Dates. Blue dots represent weekly averages and the red solid line represents the fitted line from the segmented regression model assessing for abrupt level and slope changes at the start and stop dates of the blood culture restriction of 1 blood culture set per patient per 24 hours, which were June 26, 2024 and December 23, 2024, respectively, represented by dashed vertical lines. Any hospitalizations whose admission and discharge dates overlapped the cutoff dates were excluded as a sensitivity analysis. “≥1-BC hospitalizations” designates the hospitalizations with ≥1 blood culture obtained subgroup of all hospitalizations. The figure displays the following:
(A) Mean length of time from admission to the first antimicrobial administered for ≥1-BC hospitalizations
(B) Weekly percentage of hospitalizations with ≥1 antimicrobial administered for all hospitalizations
(C) Weekly percentage of hospitalizations with ≥1 antimicrobial administered for ≥1-BC hospitalizations
(D) Mean number of days of antimicrobial therapy for all hospitalizations
(E) Mean number of days of antimicrobial therapy for ≥1-BC hospitalizations
(F) Mean length of time from admission to the first broad-spectrum antimicrobial administered for ≥1-BC hospitalizations
(G) Weekly percentage of hospitalizations with ≥1 broad-spectrum antimicrobial administered for all hospitalizations
(H) Weekly percentage of hospitalizations with ≥1 broad-spectrum antimicrobial administered for ≥1-BC hospitalizations
(I) Mean number of days of broad-spectrum antimicrobial therapy for all hospitalizations
(J) Mean number of days of broad-spectrum antimicrobial therapy for ≥1-BC hospitalizations
(K) Mean length of time from admission to the first non-broad-spectrum antimicrobial administered for ≥1-BC hospitalizations
(L) Weekly percentage of hospitalizations with ≥1 non-broad-spectrum antimicrobial administered for all hospitalizations
(M) Weekly percentage of hospitalizations with ≥1 non-broad-spectrum antimicrobial administered for ≥1-BC hospitalizations
(N) Mean number of days of non-broad-spectrum antimicrobial therapy for all hospitalizations
(O) Mean number of days of non-broad-spectrum antimicrobial therapy for ≥1-BC hospitalizations

### **eTables.**

#### **eTable 1.** Sociodemographic and Clinical Characteristics of Patients Hospitalized at the Hospital of the University of Pennsylvania Aggregated by Time Period.

|  | **All hospitalizations** | | | | **Hospitalizations with ≥1 blood culture obtained** | | | |
| --- | --- | --- | --- | --- | --- | --- | --- | --- |
| **Characteristic** | **Pre-restriction (n = 37 639)** | **Restriction (n = 18 576)** | **Post- restriction (n = 19 697)** | **Overall (n = 75 912)** | **Pre-restriction (n = 11 149)** | **Restriction (n = 4 463)** | **Post-restriction (n = 5 159)** | **Overall (n = 20 771)** |
| **Female sex, No. (%)** | 20 146 (53.5) | 9 840 (53.0) | 10 366 (52.6) | 40 352 (53.2) | 5 211 (46.7) | 2 062 (46.2) | 2 331 (45.2) | 9 604 (46.2) |
| **Age, years, No. (%)** |  | | | | | | | |
| <25 | 1 789 (4.8) | 895 (4.8) | 815 (4.1) | 3 499 (4.6) | 271 (2.4) | 120 (2.7) | 115 (2.2) | 506 (2.4) |
| 25–34 | 5 045 (13.4) | 2 354 (12.7) | 2 578 (13.1) | 9 977 (13.1) | 956 (8.6) | 349 (7.8) | 435 (8.4) | 1 740 (8.4) |
| 35–44 | 4 507 (12.0) | 2 175 (11.7) | 2 409 (12.2) | 9 091 (12.0) | 1 213 (10.9) | 470 (10.5) | 564 (10.9) | 2 247 (10.8) |
| 45–54 | 4 488 (11.9) | 2 197 (11.8) | 2 264 (11.5) | 8 949 (11.8) | 1 465 (13.1) | 618 (13.8) | 670 (13.0) | 2 753 (13.3) |
| 55–64 | 7 324 (19.5) | 3 593 (19.3) | 3 847 (19.5) | 14 764 (19.4) | 2 392 (21.5) | 1 001 (22.4) | 1 167 (22.6) | 4 560 (22.0) |
| 65–74 | 8 676 (23.1) | 4 344 (23.4) | 4 577 (23.2) | 17 597 (23.2) | 2 899 (26.0) | 1 114 (25.0) | 1 291 (25.0) | 5 304 (25.5) |
| 75–84 | 4 748 (12.6) | 2 412 (13.0) | 2 578 (13.1) | 9 738 (12.8) | 1 568 (14.1) | 636 (14.3) | 730 (14.2) | 2 934 (14.1) |
| ≥85 | 1 062 (2.8) | 606 (3.3) | 629 (3.2) | 2 297 (3.0) | 385 (3.5) | 155 (3.5) | 187 (3.6) | 727 (3.5) |
| **Race, No. (%)** |  | | | | | | | |
| Asian | 1 346 (3.6) | 709 (3.8) | 772 (3.9) | 2 827 (3.7) | 403 (3.6) | 179 (4.0) | 188 (3.6) | 770 (3.7) |
| Black | 12 466 (33.1) | 6 024 (32.4) | 6 624 (33.6) | 25 114 (33.1) | 3 721 (33.4) | 1 393 (31.2) | 1 756 (34.0) | 6 870 (33.1) |
| Other | 1 729 (4.6) | 920 (5.0) | 1 058 (5.4) | 3 707 (4.9) | 481 (4.3) | 222 (5.0) | 296 (5.7) | 999 (4.8) |
| Unknown | 1 873 (5.0) | 1 007 (5.4) | 1 039 (5.3) | 3 919 (5.2) | 585 (5.2) | 267 (6.0) | 289 (5.6) | 1 141 (5.5) |
| White | 20 225 (53.7) | 9 916 (53.4) | 10 204 (51.8) | 40 345 (53.1) | 5 959 (53.4) | 2 402 (53.8) | 2 630 (51.0) | 10 991 (52.9) |
| **Ethnicity, No. (%)** |  | | | | | | | |
| Hispanic/Latino | 1 651 (4.4) | 823 (4.4) | 859 (4.4) | 3 333 (4.4) | 463 (4.2) | 197 (4.4) | 225 (4.4) | 885 (4.3) |
| Non-Hispanic/Latino | 35 330 (93.9) | 17 392 (93.6) | 18 410 (93.5) | 71 132 (93.7) | 10 500 (94.2) | 4 164 (93.3) | 4 828 (93.6) | 19 492 (93.8) |
| Unknown | 658 (1.7) | 361 (1.9) | 428 (2.2) | 1 447 (1.9) | 186 (1.7) | 102 (2.3) | 106 (2.1) | 394 (1.9) |
| **Insurance status, No. (%)** |  | | | | | | | |
| Private or commercial | 14 032 (37.3) | 6 928 (37.3) | 7 452 (37.8) | 28 412 (37.4) | 3 572 (32.0) | 1 435 (32.2) | 1 715 (33.2) | 6 722 (32.4) |
| Public, managed | 13 245 (35.2) | 6 533 (35.2) | 6 851 (34.8) | 26 629 (35.1) | 4 213 (37.8) | 1 693 (37.9) | 1 925 (37.3) | 7 831 (37.7) |
| Public, unmanaged or uninsured/self-pay | 10 362 (27.5) | 5 115 (27.5) | 5 394 (27.4) | 20 871 (27.5) | 3 364 (30.2) | 1 335 (29.9) | 1 519 (29.4) | 6 218 (29.9) |
| **Elixhauser Comorbidity Index, No. (%)** |  | | | | | | | |
| <0 | 4 231 (11.2) | 1 840 (9.9) | 1 853 (9.4) | 7 924 (10.4) | 447 (4.0) | 153 (3.4) | 137 (2.7) | 737 (3.5) |
| 0-9 | 11 445 (30.4) | 5 374 (28.9) | 6 397 (32.5) | 23 216 (30.6) | 1 974 (17.7) | 624 (14.0) | 994 (19.3) | 3 592 (17.3) |
| 10–19 | 7 665 (20.4) | 3 847 (20.7) | 4 040 (20.5) | 15 552 (20.5) | 2 456 (22.0) | 896 (20.1) | 1 120 (21.7) | 4 472 (21.5) |
| 20–29 | 6 218 (16.5) | 3 241 (17.4) | 3 262 (16.6) | 12 721 (16.8) | 2 453 (22.0) | 1 033 (23.1) | 1 136 (22.0) | 4 622 (22.3) |
| ≥30 | 8 080 (21.5) | 4 274 (23.0) | 4 145 (21.0) | 16 499 (21.7) | 3 819 (34.3) | 1 757 (39.4) | 1 772 (34.3) | 7 348 (35.4) |

The pre-restriction period was June 26, 2023 to June 25, 2024; the restriction period was June 26, 2024 to December 23, 2024; and the post-restriction period was December 24, 2024 to June 25, 2025. All data shown are the number of hospitalizations having the indicated characteristic, with percentage in parentheses reflecting this as the numerator and the size of the overall group as the denominator (shown in the headings).

#### **eTable 2.** Sociodemographic and Clinical Characteristics of Patients Hospitalized at Penn Presbyterian Medical Center Aggregated by Time Period.

|  | **All hospitalizations** | | | | **Hospitalizations with ≥1 blood culture obtained** | | | |
| --- | --- | --- | --- | --- | --- | --- | --- | --- |
| **Characteristic** | **Pre-restriction (n = 17 142)** | **Restriction (n = 8 717)** | **Post-restriction (n = 9 113)** | **Overall (n = 34 972)** | **Pre-restriction (n = 5 078)** | **Restriction (n = 1 781)** | **Post-restriction (n = 2 335)** | **Overall (n = 9 194)** |
| **Female sex, No. (%)** | 7 985 (46.6) | 4 186 (48.0) | 4 274 (46.9) | 16 445 (47.0) | 2 297 (45.2) | 769 (43.2) | 1 036 (44.4) | 4 102 (44.6) |
| **Age, years, No. (%)** |  | | | | | | | |
| <25 | 427 (2.5) | 207 (2.4) | 186 (2.0) | 820 (2.3) | 109 (2.1) | 40 (2.2) | 43 (1.8) | 192 (2.1) |
| 25–34 | 1 227 (7.2) | 562 (6.4) | 565 (6.2) | 2 354 (6.7) | 431 (8.5) | 124 (7.0) | 146 (6.3) | 701 (7.6) |
| 35–44 | 1 577 (9.2) | 742 (8.5) | 925 (10.2) | 3 244 (9.3) | 578 (11.4) | 245 (13.8) | 326 (14.0) | 1 149 (12.5) |
| 45–54 | 1 912 (11.2) | 971 (11.1) | 994 (10.9) | 3 877 (11.1) | 613 (12.1) | 232 (13.0) | 278 (11.9) | 1 123 (12.2) |
| 55–64 | 3 539 (20.6) | 1 806 (20.7) | 1 841 (20.2) | 7 186 (20.5) | 1 005 (19.8) | 360 (20.2) | 464 (19.9) | 1 829 (19.9) |
| 65–74 | 4 531 (26.4) | 2 307 (26.5) | 2 322 (25.5) | 9 160 (26.2) | 1 199 (23.6) | 413 (23.2) | 552 (23.6) | 2 164 (23.5) |
| 75–84 | 2 773 (16.2) | 1 503 (17.2) | 1 667 (18.3) | 5 943 (17.0) | 750 (14.8) | 254 (14.3) | 371 (15.9) | 1 375 (15.0) |
| ≥85 | 1 156 (6.7) | 619 (7.1) | 613 (6.7) | 2 388 (6.8) | 393 (7.7) | 113 (6.3) | 155 (6.6) | 661 (7.2) |
| **Race, No. (%)** |  | | | | | | | |
| Asian | 352 (2.1) | 169 (1.9) | 161 (1.8) | 682 (2.0) | 104 (2.0) | 40 (2.2) | 40 (1.7) | 184 (2.0) |
| Black | 8 722 (50.9) | 4 392 (50.4) | 4 658 (51.1) | 17 772 (50.8) | 3 029 (59.6) | 1 062 (59.6) | 1 379 (59.1) | 5 470 (59.5) |
| Other | 654 (3.8) | 301 (3.5) | 382 (4.2) | 1 337 (3.8) | 202 (4.0) | 54 (3.0) | 94 (4.0) | 350 (3.8) |
| Unknown | 612 (3.6) | 319 (3.7) | 381 (4.2) | 1 312 (3.8) | 184 (3.6) | 75 (4.2) | 95 (4.1) | 354 (3.9) |
| White | 6 802 (39.7) | 3 536 (40.6) | 3 531 (38.7) | 13 869 (39.7) | 1 559 (30.7) | 550 (30.9) | 727 (31.1) | 2 836 (30.8) |
| **Ethnicity, No. (%)** |  |  |  |  |  |  |  |  |
| Hispanic/Latino | 583 (3.4) | 255 (2.9) | 278 (3.1) | 1 116 (3.2) | 172 (3.4) | 52 (2.9) | 68 (2.9) | 292 (3.2) |
| Non-Hispanic/Latino | 16 272 (94.9) | 8 312 (95.4) | 8 684 (95.3) | 33 268 (95.1) | 4 816 (94.8) | 1 698 (95.3) | 2 230 (95.5) | 8 744 (95.1) |
| Unknown | 287 (1.7) | 150 (1.7) | 151 (1.7) | 588 (1.7) | 90 (1.8) | 31 (1.7) | 37 (1.6) | 158 (1.7) |
| **Insurance status, No. (%)** |  | | | | | | | |
| Private or commercial | 3 979 (23.2) | 1 946 (22.3) | 1 952 (21.4) | 7 877 (22.5) | 857 (16.9) | 310 (17.4) | 381 (16.3) | 1 548 (16.8) |
| Public, managed | 8 095 (47.2) | 4 151 (47.6) | 4 598 (50.5) | 16 844 (48.2) | 2 849 (56.1) | 1 001 (56.2) | 1 367 (58.5) | 5 217 (56.7) |
| Public, unmanaged or uninsured/self-pay | 5 068 (29.6) | 2 620 (30.1) | 2 563 (28.1) | 10 251 (29.3) | 1 372 (27.0) | 470 (26.4) | 587 (25.1) | 2 429 (26.4) |
| **Elixhauser Comorbidity Index, No. (%)** |  | | | | | | | |
| <0 | 2 355 (13.7) | 1 040 (11.9) | 1 041 (11.4) | 4 436 (12.7) | 331 (6.5) | 70 (3.9) | 86 (3.7) | 487 (5.3) |
| 0-9 | 5 233 (30.5) | 2 557 (29.3) | 2 866 (31.4) | 10 656 (30.5) | 1 149 (22.6) | 313 (17.6) | 563 (24.1) | 2 025 (22.0) |
| 10–19 | 3 943 (23.0) | 2 047 (23.5) | 2 120 (23.3) | 8 110 (23.2) | 1 303 (25.7) | 474 (26.6) | 567 (24.3) | 2 344 (25.5) |
| 20–29 | 2 924 (17.1) | 1 656 (19.0) | 1 645 (18.1) | 6 225 (17.8) | 1 020 (20.1) | 403 (22.6) | 502 (21.5) | 1 925 (20.9) |
| ≥30 | 2 687 (15.7) | 1 417 (16.3) | 1 441 (15.8) | 5 545 (15.9) | 1 275 (25.1) | 521 (29.3) | 617 (26.4) | 2 413 (26.2) |

The pre-restriction period was June 26, 2023 to June 25, 2024; the restriction period was June 26, 2024 to December 23, 2024; and the post-restriction period was December 24, 2024 to June 25, 2025. All data shown are the number of hospitalizations having the indicated characteristic, with percentage in parentheses reflecting this as the numerator and the size of the overall group as the denominator (shown in the headings).

#### **eTable 3.** Sociodemographic and Clinical Characteristics of Patients Hospitalized at Pennsylvania Hospital Aggregated by Time Period.

|  | **All hospitalizations** | | | | **Hospitalizations with ≥1 blood culture obtained** | | | |
| --- | --- | --- | --- | --- | --- | --- | --- | --- |
| **Characteristic** | **Pre-restriction (n = 17 919)** | **Restriction (n = 9 039)** | **Post-restriction (n = 9 372)** | **Overall (n = 36 330)** | **Pre-restriction (n = 3 869)** | **Restriction (n = 1 314)** | **Post-restriction (n = 1 758)** | **Overall (n = 6 941)** |
| **Female sex, No. (%)** | 12 041 (67.2) | 6 080 (67.3) | 6 024 (64.3) | 24 145 (66.5) | 1 955 (50.5) | 645 (49.1) | 832 (47.3) | 3 432 (49.4) |
| **Age, years, No. (%)** |  | | | | | | | |
| <25 | 958 (5.3) | 484 (5.4) | 469 (5.0) | 1 911 (5.3) | 85 (2.2) | 35 (2.7) | 31 (1.8) | 151 (2.2) |
| 25–34 | 4 358 (24.3) | 2 199 (24.3) | 2 104 (22.4) | 8 661 (23.8) | 424 (11.0) | 135 (10.3) | 176 (10.0) | 735 (10.6) |
| 35–44 | 3 092 (17.3) | 1 622 (17.9) | 1 661 (17.7) | 6 375 (17.5) | 480 (12.4) | 184 (14.0) | 259 (14.7) | 923 (13.3) |
| 45–54 | 1 426 (8.0) | 699 (7.7) | 737 (7.9) | 2 862 (7.9) | 436 (11.3) | 156 (11.9) | 193 (11.0) | 785 (11.3) |
| 55–64 | 2 272 (12.7) | 1 093 (12.1) | 1 174 (12.5) | 4 539 (12.5) | 654 (16.9) | 245 (18.6) | 293 (16.7) | 1 192 (17.2) |
| 65–74 | 3 066 (17.1) | 1 605 (17.8) | 1 616 (17.2) | 6 287 (17.3) | 863 (22.3) | 280 (21.3) | 380 (21.6) | 1 523 (21.9) |
| 75–84 | 1 959 (10.9) | 944 (10.4) | 1 125 (12.0) | 4 028 (11.1) | 599 (15.5) | 172 (13.1) | 269 (15.3) | 1 040 (15.0) |
| ≥85 | 788 (4.4) | 393 (4.3) | 486 (5.2) | 1 667 (4.6) | 328 (8.5) | 107 (8.1) | 157 (8.9) | 592 (8.5) |
| **Race, No. (%)** |  | | | | | | | |
| Asian | 759 (4.2) | 418 (4.6) | 389 (4.2) | 1 566 (4.3) | 137 (3.5) | 60 (4.6) | 65 (3.7) | 262 (3.8) |
| Black | 6 114 (34.1) | 3 029 (33.5) | 3 231 (34.5) | 12 374 (34.1) | 1 402 (36.2) | 472 (35.9) | 635 (36.1) | 2 509 (36.1) |
| Other | 1 410 (7.9) | 715 (7.9) | 706 (7.5) | 2 831 (7.8) | 268 (6.9) | 94 (7.2) | 122 (6.9) | 484 (7.0) |
| Unknown | 375 (2.1) | 190 (2.1) | 173 (1.8) | 738 (2.0) | 82 (2.1) | 37 (2.8) | 34 (1.9) | 153 (2.2) |
| White | 9 261 (51.7) | 4 687 (51.9) | 4 873 (52.0) | 18 821 (51.8) | 1 980 (51.2) | 651 (49.5) | 902 (51.3) | 3 533 (50.9) |
| **Ethnicity, No. (%)** |  | | | | | | | |
| Hispanic/Latino | 1 480 (8.3) | 790 (8.7) | 744 (7.9) | 3 014 (8.3) | 263 (6.8) | 90 (6.8) | 119 (6.8) | 472 (6.8) |
| Non-Hispanic/Latino | 16 201 (90.4) | 8 142 (90.1) | 8 510 (90.8) | 32 853 (90.4) | 3 552 (91.8) | 1 206 (91.8) | 1 627 (92.5) | 6 385 (92.0) |
| Unknown | 238 (1.3) | 107 (1.2) | 118 (1.3) | 463 (1.3) | 54 (1.4) | 18 (1.4) | 12 (0.7) | 84 (1.2) |
| **Insurance status, No. (%)** |  | | | | | | | |
| Private or commercial | 6 625 (37.0) | 3 478 (38.5) | 3 337 (35.6) | 13 440 (37.0) | 754 (19.5) | 281 (21.4) | 336 (19.1) | 1 371 (19.8) |
| Public, managed | 6 732 (37.6) | 3 280 (36.3) | 3 669 (39.1) | 13 681 (37.7) | 1 900 (49.1) | 649 (49.4) | 922 (52.4) | 3 471 (50.0) |
| Public, unmanaged or uninsured/self-pay | 4 562 (25.5) | 2 281 (25.2) | 2 366 (25.2) | 9 209 (25.3) | 1 215 (31.4) | 384 (29.2) | 500 (28.4) | 2 099 (30.2) |
| **Elixhauser Comorbidity Index, No. (%)** |  | | | | | | | |
| <0 | 3 406 (19.0) | 1 626 (18.0) | 1 602 (17.1) | 6 634 (18.3) | 289 (7.5) | 80 (6.1) | 111 (6.3) | 480 (6.9) |
| 0-9 | 8 475 (47.3) | 4 355 (48.2) | 4 558 (48.6) | 17 388 (47.9) | 1 162 (30.0) | 342 (26.0) | 530 (30.1) | 2 034 (29.3) |
| 10–19 | 2 735 (15.3) | 1 376 (15.2) | 1 532 (16.3) | 5 643 (15.5) | 982 (25.4) | 351 (26.7) | 454 (25.8) | 1 787 (25.7) |
| 20–29 | 1 824 (10.2) | 923 (10.2) | 898 (9.6) | 3 645 (10.0) | 666 (17.2) | 242 (18.4) | 306 (17.4) | 1 214 (17.5) |
| ≥30 | 1 479 (8.3) | 759 (8.4) | 782 (8.3) | 3 020 (8.3) | 770 (19.9) | 299 (22.8) | 357 (20.3) | 1 426 (20.5) |

The pre-restriction period was June 26, 2023 to June 25, 2024; the restriction period was June 26, 2024 to December 23, 2024; and the post-restriction period was December 24, 2024 to June 25, 2025. All data shown are the number of hospitalizations having the indicated characteristic, with percentage in parentheses reflecting this as the numerator and the size of the overall group as the denominator (shown in the headings).

#### **eTable 4.** Primary and Secondary Outcomes at All 3 Hospital Sites Combined Aggregated by Time Period.

|  | **All hospitalizations** | | | | **Hospitalizations with ≥1 blood culture obtained (≥1-BC hospitalizations)** | | | |
| --- | --- | --- | --- | --- | --- | --- | --- | --- |
| **Outcome** | **Pre-restriction (n = 72 700)** | **Restriction (n = 36 332)** | **Post-restriction (n = 38 182)** | **Overall (n = 147 214)** | **Pre-restriction (n = 20 096)** | **Restriction (n = 7 558)** | **Post-restriction (n = 9 252)** | **Overall (n = 36 906)** |
| Overall blood culture positivity, No./total (%)^a^ | 2 964/61 664 (4.8) | 1 055/13 377 (7.9) | 1 913/28 047 (6.8) | 5 932/103 088 (5.8) | NA | NA | NA | NA |
| In-hospital mortality or hospice discharge, No. (%) | 1 802 (2.5) | 683 (1.9) | 724 (1.9) | 3 209 (2.2) | 1 082 (5.4) | 361 (4.8) | 420 (4.5) | 1 863 (5.0) |
| Total 30-day revisits, mean (SD) | 0.36 (1.00) | 0.35 (0.95) | 0.36 (1.09) | 0.36 (1.01) | 0.36 (0.82) | 0.37 (0.93) | 0.36 (0.87) | 0.36 (0.86) |
| 30-day inpatient readmission, mean (SD) | 0.23 (0.75) | 0.22 (0.66) | 0.25 (0.88) | 0.23 (0.77) | 0.22 (0.52) | 0.23 (0.54) | 0.22 (0.52) | 0.22 (0.52) |
| 30-day medical observation unit revisits, mean (SD) | 0.03 (0.19) | 0.03 (0.19) | 0.03 (0.18) | 0.03 (0.19) | 0.03 (0.20) | 0.04 (0.21) | 0.03 (0.18) | 0.03 (0.20) |
| 30-day emergency department revisits, mean (SD) | 0.09 (0.53) | 0.10 (0.57) | 0.09 (0.52) | 0.09 (0.54) | 0.11 (0.52) | 0.11 (0.64) | 0.11 (0.57) | 0.11 (0.56) |
| Length of stay, days, mean (SD) | 6.35 (9.09) | 6.62 (9.87) | 6.73 (9.62) | 6.51 (9.43) | 11.46 (14.38) | 13.39 (17.19) | 13.03 (15.75) | 12.25 (15.36) |
| ≥1 blood culture obtained, No. (%) | 20 096 (27.6) | 7 558 (20.8) | 9 252 (24.2) | 36 906 (25.1) | 20 096 (100.0) | 7 558 (100.0) | 9 252 (100.0) | 36 906 (100.0) |
| Total number of blood cultures obtained, mean (SD)^b^ | NA | NA | NA | NA | 3.07 (3.14) | 1.77 (2.03) | 3.03 (3.01) | 2.79 (2.96) |
| Number of calendar days with ≥1 blood culture obtained, mean (SD)^b^ | NA | NA | NA | NA | 1.71 (1.73) | 1.67 (1.70) | 1.76 (1.74) | 1.71 (1.73) |
| Time from admission to first blood culture obtained, days, mean (SD)^b^ | NA | NA | NA | NA | 1.23 (3.42) | 1.89 (4.43) | 1.68 (4.22) | 1.48 (3.86) |
| ≥1 positive blood culture obtained, No. (%)^b^ | NA | NA | NA | NA | 2 235 (11.1) | 853 (11.3) | 1 373 (14.8) | 4 461 (12.1) |
| Total number of positive blood cultures obtained, mean (SD)^b^ | NA | NA | NA | NA | 0.15 (0.48) | 0.14 (0.45) | 0.21 (0.59) | 0.16 (0.51) |
| Number of calendar days with ≥1 positive blood culture obtained, mean (SD)^b^ | NA | NA | NA | NA | 0.13 (0.40) | 0.13 (0.40) | 0.18 (0.49) | 0.14 (0.43) |
| ≥1 antimicrobial administered, No. (%) | 40 058 (55.1) | 19 899 (54.8) | 21 172 (55.5) | 81 129 (55.1) | 17 544 (87.3) | 6 835 (90.4) | 8 268 (89.4) | 32 647 (88.5) |
| Days of therapy with ≥1 antimicrobial administered, mean (SD) | 2.99 (6.27) | 3.04 (7.17) | 3.11 (6.92) | 3.03 (6.67) | 7.44 (9.89) | 8.80 (13.14) | 8.36 (11.68) | 7.95 (11.10) |
| Time from admission to first antimicrobial administered, days, mean (SD) | 1.06 (2.31) | 1.13 (2.58) | 1.12 (2.43) | 1.10 (2.41) | 1.06 (2.70) | 1.20 (3.32) | 1.16 (2.89) | 1.11 (2.89) |
| ≥1 broad-spectrum antimicrobial administered, No. (%) | 25 122 (34.6) | 12 456 (34.3) | 13 246 (34.7) | 50 824 (34.5) | 14 765 (73.5) | 5 976 (79.1) | 7 135 (77.1) | 27 876 (75.5) |
| Days of therapy with ≥1 broad-spectrum antimicrobial administered, mean (SD) | 1.90 (5.09) | 1.92 (5.64) | 1.98 (5.63) | 1.93 (5.38) | 5.33 (8.28) | 6.36 (10.45) | 6.03 (9.73) | 5.72 (9.15) |
| Time from admission to first broad-spectrum antimicrobial administered, days, mean (SD) | 1.51 (3.27) | 1.60 (3.60) | 1.64 (3.63) | 1.57 (3.45) | 1.57 (3.71) | 1.77 (4.41) | 1.77 (4.15) | 1.67 (3.98) |
| ≥1 non-broad-spectrum antimicrobial administered, No. (%) | 31 820 (43.8) | 15 649 (43.1) | 16 636 (43.6) | 64 105 (43.5) | 13 215 (65.8) | 5 075 (67.1) | 6 167 (66.7) | 24 457 (66.3) |
| Days of therapy with ≥1 non-broad-spectrum antimicrobial administered, mean (SD) | 1.71 (4.09) | 1.72 (4.71) | 1.75 (4.49) | 1.72 (4.35) | 3.87 (6.68) | 4.49 (8.99) | 4.28 (7.89) | 4.10 (7.52) |
| Time from admission to first non-broad-spectrum antimicrobial administered, days, mean (SD) | 1.62 (3.59) | 1.68 (3.91) | 1.66 (3.40) | 1.65 (3.63) | 2.20 (4.89) | 2.49 (5.81) | 2.37 (4.66) | 2.30 (5.04) |

The pre-restriction period was June 26, 2023 to June 25, 2024; the restriction period was June 26, 2024 to December 23, 2024; and the post-restriction period was December 24, 2024 to June 25, 2025. The total 30-day readmission outcome is comprised of all readmissions, including inpatient readmissions, medical observation unit revisits, and emergency department revisits. The total number and percentage of hospitalizations in the associated time period is shown for binary outcomes (in-hospital mortality or hospice discharge, ≥1 blood culture obtained, ≥1 positive blood culture, ≥1 antimicrobial administered, ≥1 broad-spectrum antimicrobial administered, and ≥1 non-broad-spectrum antimicrobial administered). The mean number of hospitalizations and standard deviation in the associated time period is shown for count and continuous outcomes (30-day inpatient readmission outcomes, length of stay, total numbers of blood cultures and positive blood cultures, number of calendar days with ≥1 blood culture or ≥1 positive blood culture, time from admission to first blood culture or first positive blood culture, and days of therapy with ≥1 antimicrobial administered including broad and non-broad-spectrum). All of these outcomes reflected outcomes associated with individual hospitalizations. The overall blood culture positivity rate was the exception, as detailed below.

^a^ The overall blood culture positivity rate reflects all 3 hospital sites in the aggregate rather than an averaged measure for individual hospitalizations, and thus is not shown for the hospitalizations with ≥1 blood culture obtained subgroup. As an exception to all other outcomes shown here, both the numerator and denominator are explicitly shown, with the numerator describing the total number of positive blood cultures and the denominator describing the total number of blood cultures obtained across all 3 hospital sites for that time period.

^b^ Outcomes were only calculated for the hospitalizations with ≥1 blood culture obtained subgroup.

#### **eTable 5.** Primary and Secondary Outcomes at the Hospital of the University of Pennsylvania Aggregated by Time Period.

|  | **All hospitalizations** | | | | **Hospitalizations with ≥1 blood culture obtained (≥1-BC hospitalizations)** | | | |
| --- | --- | --- | --- | --- | --- | --- | --- | --- |
| **Outcome** | **Pre-restriction (n = 37 639)** | **Restriction (n = 18 576)** | **Post-restriction (n = 19 697)** | **Overall (n = 75 912)** | **Pre-restriction (n = 11 149)** | **Restriction (n = 4 463)** | **Post-restriction (n = 5 159)** | **Overall (n = 20 771)** |
| Overall blood culture positivity, No./total (%)^a^ | 1 360/37 258 (3.7) | 555/8 493 (6.5) | 932/17 295 (5.4) | 2 847/63 046 (4.5) | NA | NA | NA | NA |
| In-hospital mortality or hospice discharge, No. (%) | 1 079 (2.9) | 420 (2.3) | 453 (2.3) | 1 952 (2.6) | 672 (6.0) | 228 (5.1) | 261 (5.1) | 1 161 (5.6) |
| Total 30-day revisits, mean (SD) | 0.35 (0.90) | 0.35 (0.89) | 0.37 (0.93) | 0.35 (0.90) | 0.35 (0.75) | 0.36 (0.91) | 0.34 (0.79) | 0.35 (0.80) |
| 30-day inpatient readmission, mean (SD) | 0.26 (0.76) | 0.25 (0.71) | 0.28 (0.79) | 0.26 (0.76) | 0.24 (0.54) | 0.24 (0.57) | 0.23 (0.53) | 0.24 (0.54) |
| 30-day medical observation unit revisits, mean (SD) | 0.03 (0.17) | 0.03 (0.18) | 0.03 (0.18) | 0.03 (0.17) | 0.03 (0.19) | 0.03 (0.20) | 0.03 (0.17) | 0.03 (0.19) |
| 30-day emergency department revisits, mean (SD) | 0.06 (0.36) | 0.07 (0.44) | 0.06 (0.36) | 0.06 (0.38) | 0.08 (0.41) | 0.09 (0.61) | 0.08 (0.48) | 0.08 (0.48) |
| Length of stay, days, mean (SD) | 7.13 (10.40) | 7.45 (10.93) | 7.57 (11.15) | 7.32 (10.73) | 13.04 (15.99) | 14.56 (18.08) | 14.85 (17.95) | 13.82 (16.97) |
| ≥1 blood culture obtained, No. (%) | 11 149 (29.6) | 4 463 (24.0) | 5 159 (26.2) | 20 771 (27.4) | 11 149 (100.0) | 4 463 (100.0) | 5 159 (100.0) | 20 771 (100.0) |
| Total number of blood cultures obtained, mean (SD)^b^ | NA | NA | NA | NA | 3.34 (3.63) | 1.90 (2.29) | 3.35 (3.50) | 3.04 (3.40) |
| Number of calendar days with ≥1 blood culture obtained, mean (SD)^b^ | NA | NA | NA | NA | 1.87 (1.99) | 1.79 (1.91) | 1.93 (2.02) | 1.87 (1.98) |
| Time from admission to first blood culture obtained, days, mean (SD)^b^ | NA | NA | NA | NA | 1.53 (3.88) | 2.19 (4.87) | 2.09 (4.89) | 1.81 (4.38) |
| ≥1 positive blood culture obtained, No. (%)^b^ | NA | NA | NA | NA | 1 041 (9.3) | 447 (10.0) | 675 (13.1) | 2 163 (10.4) |
| Total number of positive blood cultures obtained, mean (SD)^b^ | NA | NA | NA | NA | 0.12 (0.45) | 0.12 (0.43) | 0.18 (0.56) | 0.14 (0.48) |
| Number of calendar days with ≥1 positive blood culture obtained, mean (SD)^b^ | NA | NA | NA | NA | 0.11 (0.39) | 0.12 (0.39) | 0.17 (0.50) | 0.13 (0.42) |
| ≥1 antimicrobial administered, No. (%) | 21 007 (55.8) | 10 398 (56.0) | 11 082 (56.3) | 42 487 (56.0) | 9 917 (88.9) | 4 072 (91.2) | 4 678 (90.7) | 18 667 (89.9) |
| Days of therapy with ≥1 antimicrobial administered, mean (SD) | 3.61 (7.55) | 3.67 (8.36) | 3.76 (8.44) | 3.66 (7.99) | 8.83 (11.53) | 9.93 (14.28) | 9.92 (13.88) | 9.34 (12.78) |
| Time from admission to first antimicrobial administered, days, mean (SD) | 1.24 (2.67) | 1.33 (2.94) | 1.34 (2.82) | 1.29 (2.78) | 1.22 (3.12) | 1.34 (3.68) | 1.38 (3.38) | 1.29 (3.32) |
| ≥1 broad-spectrum antimicrobial administered, No. (%) | 13 956 (37.1) | 6 948 (37.4) | 7 391 (37.5) | 28 295 (37.3) | 8 714 (78.2) | 3 640 (81.6) | 4 187 (81.2) | 16 541 (79.6) |
| Days of therapy with ≥1 broad-spectrum antimicrobial administered, mean (SD) | 2.46 (6.20) | 2.47 (6.93) | 2.56 (6.93) | 2.49 (6.57) | 6.63 (9.69) | 7.40 (12.09) | 7.39 (11.58) | 6.98 (10.73) |
| Time from admission to first broad-spectrum antimicrobial administered, days, mean (SD) | 1.79 (3.74) | 1.93 (4.08) | 1.98 (4.13) | 1.87 (3.93) | 1.79 (4.16) | 2.01 (4.87) | 2.05 (4.67) | 1.90 (4.46) |
| ≥1 non-broad-spectrum antimicrobial administered, No. (%) | 16 081 (42.7) | 7 906 (42.6) | 8 387 (42.6) | 32 374 (42.6) | 7 406 (66.4) | 3 023 (67.7) | 3 428 (66.4) | 13 857 (66.7) |
| Days of therapy with ≥1 non-broad-spectrum antimicrobial administered, mean (SD) | 1.97 (4.94) | 1.99 (5.23) | 2.02 (5.45) | 1.99 (5.14) | 4.51 (7.87) | 5.03 (9.21) | 4.99 (9.36) | 4.74 (8.56) |
| Time from admission to first non-broad-spectrum antimicrobial administered, days, mean (SD) | 1.92 (4.24) | 2.08 (4.85) | 2.00 (3.96) | 1.98 (4.33) | 2.54 (5.57) | 2.84 (6.83) | 2.75 (5.24) | 2.66 (5.79) |

The pre-restriction period was June 26, 2023 to June 25, 2024; the restriction period was June 26, 2024 to December 23, 2024; and the post-restriction period was December 24, 2024 to June 25, 2025. The total 30-day readmission outcome is comprised of all readmissions, including inpatient readmissions, medical observation unit revisits, and emergency department revisits. The total number and percentage of hospitalizations in the associated time period is shown for binary outcomes (in-hospital mortality or hospice discharge, ≥1 blood culture obtained, ≥1 positive blood culture, ≥1 antimicrobial administered, ≥1 broad-spectrum antimicrobial administered, and ≥1 non-broad-spectrum antimicrobial administered). The mean number of hospitalizations and standard deviation in the associated time period is shown for count and continuous outcomes (30-day inpatient readmission outcomes, length of stay, total numbers of blood cultures and positive blood cultures, number of calendar days with ≥1 blood culture or ≥1 positive blood culture, time from admission to first blood culture or first positive blood culture, and days of therapy with ≥1 antimicrobial administered including broad and non-broad-spectrum). All of these outcomes reflected outcomes associated with individual hospitalizations. The overall blood culture positivity rate was the exception, as detailed below.

^a^ The overall blood culture positivity rate reflects the entirety of the Hospital of the University of Pennsylvania rather than an averaged measure for individual hospitalizations, and thus is not shown for the hospitalizations with ≥1 blood culture obtained subgroup. As an exception to all other outcomes shown here, both the numerator and denominator are explicitly shown, with the numerator describing the total number of positive blood cultures and the denominator describing the total number of blood cultures obtained at the Hospital of the University of Pennsylvania for that time period.

^b^ Outcomes were only calculated for the hospitalizations with ≥1 blood culture obtained subgroup.

#### **eTable 6.** Primary and Secondary Outcomes at Penn Presbyterian Medical Center Aggregated by Time Period.

|  | **All hospitalizations** | | | | **Hospitalizations with ≥1 blood culture obtained (≥1-BC hospitalizations)** | | | |
| --- | --- | --- | --- | --- | --- | --- | --- | --- |
| **Outcome** | **Pre-restriction (n = 17 142)** | **Restriction (n = 8 717)** | **Post-restriction (n = 9 113)** | **Overall (n = 34 972)** | **Pre-restriction (n = 5 078)** | **Restriction (n = 1 781)** | **Post-restriction (n = 2 335)** | **Overall (n = 9 194)** |
| Overall blood culture positivity, No./total (%)^a^ | 745/14 671 (5.1) | 285/3 039 (9.4) | 570/6 881 (8.3) | 1 600/24 591 (6.5) | NA | NA | NA | NA |
| In-hospital mortality or hospice discharge, No. (%) | 548 (3.2) | 202 (2.3) | 205 (2.2) | 955 (2.7) | 290 (5.7) | 92 (5.2) | 113 (4.8) | 495 (5.4) |
| Total 30-day revisits, mean (SD) | 0.36 (1.06) | 0.36 (1.00) | 0.41 (1.46) | 0.37 (1.17) | 0.35 (0.83) | 0.35 (0.82) | 0.35 (0.77) | 0.35 (0.81) |
| 30-day inpatient readmission, mean (SD) | 0.23 (0.86) | 0.22 (0.74) | 0.28 (1.30) | 0.24 (0.97) | 0.20 (0.49) | 0.20 (0.48) | 0.21 (0.52) | 0.20 (0.50) |
| 30-day medical observation unit revisits, mean (SD) | 0.03 (0.16) | 0.03 (0.19) | 0.02 (0.16) | 0.03 (0.17) | 0.03 (0.18) | 0.03 (0.20) | 0.02 (0.16) | 0.03 (0.18) |
| 30-day emergency department revisits, mean (SD) | 0.11 (0.53) | 0.11 (0.55) | 0.10 (0.59) | 0.11 (0.55) | 0.12 (0.57) | 0.12 (0.56) | 0.12 (0.44) | 0.12 (0.54) |
| Length of stay, days, mean (SD) | 6.49 (8.58) | 6.73 (9.95) | 6.84 (8.66) | 6.64 (8.96) | 10.76 (12.84) | 13.07 (16.32) | 12.22 (12.93) | 11.58 (13.63) |
| ≥1 blood culture obtained, No. (%) | 5 078 (29.6) | 1 781 (20.4) | 2 335 (25.6) | 9 194 (26.3) | 5 078 (100.0) | 1 781 (100.0) | 2 335 (100.0) | 9 194 (100.0) |
| Total number of blood cultures obtained, mean (SD)^b^ | NA | NA | NA | NA | 2.89 (2.74) | 1.71 (1.81) | 2.95 (2.49) | 2.67 (2.56) |
| Number of calendar days with ≥1 blood culture obtained, mean (SD)^b^ | NA | NA | NA | NA | 1.60 (1.49) | 1.60 (1.56) | 1.66 (1.43) | 1.62 (1.49) |
| Time from admission to first blood culture obtained, days, mean (SD)^b^ | NA | NA | NA | NA | 1.04 (2.96) | 1.74 (4.18) | 1.45 (3.55) | 1.28 (3.39) |
| ≥1 positive blood culture obtained, No. (%)^b^ | NA | NA | NA | NA | 617 (12.2) | 227 (12.7) | 433 (18.5) | 1 277 (13.9) |
| Total number of positive blood cultures obtained, mean (SD)^b^ | NA | NA | NA | NA | 0.15 (0.43) | 0.16 (0.49) | 0.24 (0.61) | 0.17 (0.50) |
| Number of calendar days with ≥1 positive blood culture obtained, mean (SD)^b^ | NA | NA | NA | NA | 0.13 (0.38) | 0.15 (0.42) | 0.22 (0.50) | 0.16 (0.42) |
| ≥1 antimicrobial administered, No. (%) | 10 436 (60.9) | 5 130 (58.9) | 5 521 (60.6) | 21 087 (60.3) | 4 366 (86.0) | 1 582 (88.8) | 2 069 (88.6) | 8 017 (87.2) |
| Days of therapy with ≥1 antimicrobial administered, mean (SD) | 2.96 (5.16) | 2.95 (6.18) | 3.05 (5.52) | 2.98 (5.53) | 6.40 (7.72) | 7.92 (11.27) | 7.20 (8.49) | 6.90 (8.73) |
| Time from admission to first antimicrobial administered, days, mean (SD) | 0.95 (1.93) | 1.04 (2.46) | 0.97 (2.05) | 0.98 (2.10) | 0.93 (2.15) | 1.15 (3.15) | 0.95 (2.10) | 0.98 (2.37) |
| ≥1 broad-spectrum antimicrobial administered, No. (%) | 7 021 (41.0) | 3 399 (39.0) | 3 688 (40.5) | 14 108 (40.3) | 3 623 (71.3) | 1 383 (77.7) | 1 773 (75.9) | 6 779 (73.7) |
| Days of therapy with ≥1 broad-spectrum antimicrobial administered, mean (SD) | 1.83 (4.15) | 1.81 (4.57) | 1.85 (4.42) | 1.83 (4.33) | 4.37 (6.38) | 5.60 (8.17) | 5.00 (6.99) | 4.77 (6.93) |
| Time from admission to first broad-spectrum antimicrobial administered, days, mean (SD) | 1.17 (2.62) | 1.28 (3.16) | 1.29 (3.07) | 1.23 (2.88) | 1.33 (3.00) | 1.56 (3.97) | 1.53 (3.63) | 1.43 (3.39) |
| ≥1 non-broad-spectrum antimicrobial administered, No. (%) | 8 488 (49.5) | 4 187 (48.0) | 4 476 (49.1) | 17 151 (49.0) | 3 268 (64.4) | 1 185 (66.5) | 1 554 (66.6) | 6 007 (65.3) |
| Days of therapy with ≥1 non-broad-spectrum antimicrobial administered, mean (SD) | 1.72 (3.29) | 1.73 (4.15) | 1.78 (3.66) | 1.74 (3.62) | 3.27 (5.07) | 3.96 (7.87) | 3.66 (5.93) | 3.50 (5.93) |
| Time from admission to first non-broad-spectrum antimicrobial administered, days, mean (SD) | 1.56 (3.00) | 1.56 (3.00) | 1.60 (3.07) | 1.57 (3.02) | 2.08 (4.12) | 2.36 (4.40) | 2.28 (4.28) | 2.18 (4.22) |

The pre-restriction period was June 26, 2023 to June 25, 2024; the restriction period was June 26, 2024 to December 23, 2024; and the post-restriction period was December 24, 2024 to June 25, 2025. The total 30-day readmission outcome is comprised of all readmissions, including inpatient readmissions, medical observation unit revisits, and emergency department revisits. The total number and percentage of hospitalizations in the associated time period is shown for binary outcomes (in-hospital mortality or hospice discharge, ≥1 blood culture obtained, ≥1 positive blood culture, ≥1 antimicrobial administered, ≥1 broad-spectrum antimicrobial administered, and ≥1 non-broad-spectrum antimicrobial administered). The mean number of hospitalizations and standard deviation in the associated time period is shown for count and continuous outcomes (30-day inpatient readmission outcomes, length of stay, total numbers of blood cultures and positive blood cultures, number of calendar days with ≥1 blood culture or ≥1 positive blood culture, time from admission to first blood culture or first positive blood culture, and days of therapy with ≥1 antimicrobial administered including broad and non-broad-spectrum). All of these outcomes reflected outcomes associated with individual hospitalizations. The overall blood culture positivity rate was the exception, as detailed below.

^a^ The overall blood culture positivity rate reflects the entirety of Penn Presbyterian Medical Center rather than an averaged measure for individual hospitalizations, and thus is not shown for the hospitalizations with ≥1 blood culture obtained subgroup. As an exception to all other outcomes shown here, both the numerator and denominator are explicitly shown, with the numerator describing the total number of positive blood cultures and the denominator describing the total number of blood cultures obtained at Penn Presbyterian Medical Center for that time period.

^b^ Outcomes were only calculated for the hospitalizations with ≥1 blood culture obtained subgroup.

#### **eTable 7.** Primary and Secondary Outcomes at Pennsylvania Hospital Aggregated by Time Period.

|  | **All hospitalizations** | | | | **Hospitalizations with ≥1 blood culture obtained (≥1-BC hospitalizations)** | | | |
| --- | --- | --- | --- | --- | --- | --- | --- | --- |
| **Outcome** | **Pre-restriction (n = 17 919)** | **Restriction (n = 9 039)** | **Post-restriction (n = 9 372)** | **Overall (n = 36 330)** | **Pre-restriction (n = 3 869)** | **Restriction (n = 1 314)** | **Post-restriction (n = 1 758)** | **Overall (n = 6 941)** |
| Overall blood culture positivity, No./total (%)^a^ | 859/9 735 (8.8) | 215/1 845 (11.7) | 411/3 871 (10.6) | 1 485/15 451 (9.6) | NA | NA | NA | NA |
| In-hospital mortality or hospice discharge, No. (%) | 175 (1.0) | 61 (0.7) | 66 (0.7) | 302 (0.8) | 120 (3.1) | 41 (3.1) | 46 (2.6) | 207 (3.0) |
| Total 30-day revisits, mean (SD) | 0.37 (1.11) | 0.34 (1.05) | 0.31 (0.96) | 0.35 (1.06) | 0.43 (0.99) | 0.43 (1.11) | 0.42 (1.14) | 0.43 (1.05) |
| 30-day inpatient readmission, mean (SD) | 0.18 (0.58) | 0.15 (0.44) | 0.15 (0.43) | 0.16 (0.51) | 0.21 (0.50) | 0.20 (0.49) | 0.21 (0.50) | 0.21 (0.50) |
| 30-day medical observation unit revisits, mean (SD) | 0.04 (0.24) | 0.04 (0.23) | 0.03 (0.19) | 0.04 (0.23) | 0.05 (0.23) | 0.06 (0.28) | 0.04 (0.21) | 0.05 (0.24) |
| 30-day emergency department revisits, mean (SD) | 0.15 (0.76) | 0.15 (0.77) | 0.13 (0.70) | 0.15 (0.75) | 0.17 (0.70) | 0.17 (0.81) | 0.17 (0.88) | 0.17 (0.77) |
| Length of stay, days, mean (SD) | 4.57 (5.76) | 4.79 (6.77) | 4.85 (6.14) | 4.70 (6.12) | 7.81 (9.99) | 9.88 (14.53) | 8.77 (10.46) | 8.45 (11.13) |
| ≥1 blood culture obtained, No. (%) | 3 869 (21.6) | 1 314 (14.5) | 1 758 (18.8) | 6 941 (19.1) | 3 869 (100.0) | 1 314 (100.0) | 1 758 (100.0) | 6 941 (100.0) |
| Total number of blood cultures obtained, mean (SD)^b^ | NA | NA | NA | NA | 2.52 (1.70) | 1.40 (1.06) | 2.20 (1.59) | 2.23 (1.62) |
| Number of calendar days with ≥1 blood culture obtained, mean (SD)^b^ | NA | NA | NA | NA | 1.41 (1.01) | 1.34 (0.87) | 1.36 (0.96) | 1.38 (0.97) |
| Time from admission to first blood culture obtained, days, mean (SD)^b^ | NA | NA | NA | NA | 0.63 (2.28) | 1.08 (2.81) | 0.80 (2.31) | 0.76 (2.40) |
| ≥1 positive blood culture obtained, No. (%)^b^ | NA | NA | NA | NA | 577 (14.9) | 179 (13.6) | 265 (15.1) | 1 021 (14.7) |
| Total number of positive blood cultures obtained, mean (SD)^b^ | NA | NA | NA | NA | 0.22 (0.60) | 0.16 (0.45) | 0.23 (0.66) | 0.21 (0.59) |
| Number of calendar days with ≥1 positive blood culture obtained, mean (SD)^b^ | NA | NA | NA | NA | 0.17 (0.44) | 0.15 (0.41) | 0.17 (0.46) | 0.17 (0.44) |
| ≥1 antimicrobial administered, No. (%) | 8 615 (48.1) | 4 371 (48.4) | 4 569 (48.8) | 17 555 (48.3) | 3 261 (84.3) | 1 181 (89.9) | 1 521 (86.5) | 5 963 (85.9) |
| Days of therapy with ≥1 antimicrobial administered, mean (SD) | 1.72 (3.46) | 1.81 (4.85) | 1.78 (3.58) | 1.76 (3.88) | 4.81 (5.79) | 6.17 (10.79) | 5.35 (6.26) | 5.20 (7.13) |
| Time from admission to first antimicrobial administered, days, mean (SD) | 0.79 (1.68) | 0.76 (1.58) | 0.78 (1.69) | 0.78 (1.66) | 0.72 (1.80) | 0.77 (1.89) | 0.74 (1.97) | 0.73 (1.86) |
| ≥1 broad-spectrum antimicrobial administered, No. (%) | 4 145 (23.1) | 2 109 (23.3) | 2 167 (23.1) | 8 421 (23.2) | 2 428 (62.8) | 953 (72.5) | 1 175 (66.8) | 4 556 (65.6) |
| Days of therapy with ≥1 broad-spectrum antimicrobial administered, mean (SD) | 0.81 (2.50) | 0.89 (2.77) | 0.88 (2.73) | 0.85 (2.63) | 2.85 (4.51) | 3.89 (5.62) | 3.40 (5.14) | 3.19 (4.92) |
| Time from admission to first broad-spectrum antimicrobial administered, days, mean (SD) | 1.15 (2.35) | 1.05 (2.14) | 1.12 (2.31) | 1.12 (2.29) | 1.18 (2.77) | 1.15 (2.78) | 1.11 (2.46) | 1.16 (2.69) |
| ≥1 non-broad-spectrum antimicrobial administered, No. (%) | 7 251 (40.5) | 3 556 (39.3) | 3 773 (40.3) | 14 580 (40.1) | 2 541 (65.7) | 867 (66.0) | 1 185 (67.4) | 4 593 (66.2) |
| Days of therapy with ≥1 non-broad-spectrum antimicrobial administered, mean (SD) | 1.16 (2.41) | 1.17 (3.99) | 1.17 (2.52) | 1.16 (2.91) | 2.80 (4.07) | 3.39 (9.49) | 3.02 (4.52) | 2.97 (5.61) |
| Time from admission to first non-broad-spectrum antimicrobial administered, days, mean (SD) | 1.03 (2.40) | 0.95 (1.87) | 1.00 (2.09) | 1.00 (2.20) | 1.35 (3.33) | 1.42 (2.64) | 1.39 (2.90) | 1.38 (3.10) |

The pre-restriction period was June 26, 2023 to June 25, 2024; the restriction period was June 26, 2024 to December 23, 2024; and the post-restriction period was December 24, 2024 to June 25, 2025. The total 30-day readmission outcome is comprised of all readmissions, including inpatient readmissions, medical observation unit revisits, and emergency department revisits. The total number and percentage of hospitalizations in the associated time period is shown for binary outcomes (in-hospital mortality or hospice discharge, ≥1 blood culture obtained, ≥1 positive blood culture, ≥1 antimicrobial administered, ≥1 broad-spectrum antimicrobial administered, and ≥1 non-broad-spectrum antimicrobial administered). The mean number of hospitalizations and standard deviation in the associated time period is shown for count and continuous outcomes (30-day inpatient readmission outcomes, length of stay, total numbers of blood cultures and positive blood cultures, number of calendar days with ≥1 blood culture or ≥1 positive blood culture, time from admission to first blood culture or first positive blood culture, and days of therapy with ≥1 antimicrobial administered including broad and non-broad-spectrum). All of these outcomes reflected outcomes associated with individual hospitalizations. The overall blood culture positivity rate was the exception, as detailed below.

^a^ The overall blood culture positivity rate reflects Pennsylvania Hospital rather than an averaged measure for individual hospitalizations, and thus is not shown for the hospitalizations with ≥1 blood culture obtained subgroup. As an exception to all other outcomes shown here, both the numerator and denominator are explicitly shown, with the numerator describing the total number of positive blood cultures and the denominator describing the total number of blood cultures obtained at Pennsylvania Hospital for that time period.

^b^ Outcomes were only calculated for the hospitalizations with ≥1 blood culture obtained subgroup.

#### **eTable 8.** Interpretation of Effect Estimates of Blood Culture Restriction on Primary and Secondary Outcomes During All Hospitalizations at All 3 Hospital Sites Combined.

| **Outcome** | **Effect estimate** | **OR or IRR** | **P value** | **Interpretation (odds/rate)** |
| --- | --- | --- | --- | --- |
| In-hospital mortality or hospice discharge | Slope before blood culture restriction | 0.987 (0.983-0.990) | P< .001 | –1.3%/week |
|  | Level change upon blood culture restriction | 0.998 (0.834-1.195) | P= .98 | –0.2% (instant) |
|  | Slope during blood culture restriction | 1.006 (0.994-1.017) | P= .33 | +0.6%/week |
|  | Level change at end of blood culture restriction | 1.017 (0.802-1.290) | P= .89 | +1.7% (instant) |
|  | Slope after blood culture restriction | 0.972 (0.958-0.987) | P< .001 | –2.8%/week |
| Total 30-day revisits | Slope before blood culture restriction | 0.999 (0.998-1.001) | P= .29 | –0.1%/week |
|  | Level change upon blood culture restriction | 1.011 (0.939-1.089) | P= .77 | +1.1% (instant) |
|  | Slope during blood culture restriction | 0.998 (0.994-1.002) | P= .40 | –0.2%/week |
|  | Level change at end of blood culture restriction | 0.936 (0.856-1.024) | P= .15 | –6.4% (instant) |
|  | Slope after blood culture restriction | 1.012 (1.004-1.019) | P= .002 | +1.2%/week |
| 30-day inpatient readmission | Slope before blood culture restriction | 0.998 (0.997-1.000) | P= .03 | –0.2%/week |
|  | Level change upon blood culture restriction | 0.999 (0.906-1.101) | P= .98 | –0.1% (instant) |
|  | Slope during blood culture restriction | 0.998 (0.993-1.003) | P= .39 | –0.2%/week |
|  | Level change at end of blood culture restriction | 0.998 (0.897-1.112) | P= .98 | –0.2% (instant) |
|  | Slope after blood culture restriction | 1.013 (1.004-1.022) | P= .004 | +1.3%/week |
| 30-day medical observation unit revisits | Slope before blood culture restriction | 1.001 (0.998-1.004) | P= .50 | +0.1%/week |
|  | Level change upon blood culture restriction | 1.247 (1.109-1.402) | P< .001 | +24.7% (instant) |
|  | Slope during blood culture restriction | 0.988 (0.981-0.994) | P< .001 | –1.2%/week |
|  | Level change at end of blood culture restriction | 0.932 (0.761-1.142) | P= .50 | –6.8% (instant) |
|  | Slope after blood culture restriction | 1.015 (1.003-1.028) | P= .02 | +1.5%/week |
| 30-day emergency department revisits | Slope before blood culture restriction | 1.001 (0.999-1.004) | P= .34 | +0.1%/week |
|  | Level change upon blood culture restriction | 0.969 (0.858-1.094) | P= .61 | –3.1% (instant) |
|  | Slope during blood culture restriction | 1.003 (0.998-1.008) | P= .29 | +0.3%/week |
|  | Level change at end of blood culture restriction | 0.804 (0.707-0.914) | P< .001 | –19.6% (instant) |
|  | Slope after blood culture restriction | 1.006 (0.997-1.015) | P= .17 | +0.6%/week |
| Length of stay, days | Slope before blood culture restriction | 1.000 (0.999-1.001) | P= .57 | 0.0%/week |
|  | Level change upon blood culture restriction | 0.985 (0.949-1.023) | P= .43 | –1.5% (instant) |
|  | Slope during blood culture restriction | 1.002 (1.000-1.004) | P= .055 | +0.2%/week |
|  | Level change at end of blood culture restriction | 1.015 (0.964-1.069) | P= .58 | +1.5% (instant) |
|  | Slope after blood culture restriction | 0.993 (0.989-0.996) | P< .001 | –0.7%/week |
| Overall blood culture positivity | Slope before blood culture restriction | 1.002 (0.999-1.005) | P= .13 | +0.2%/week |
|  | Level change upon blood culture restriction | 1.362 (1.138-1.631) | P< .001 | +36.2% (instant) |
|  | Slope during blood culture restriction | 1.011 (1.001-1.021) | P= .03 | +1.1%/week |
|  | Level change at end of blood culture restriction | 0.730 (0.617-0.864) | P< .001 | –27.0% (instant) |
|  | Slope after blood culture restriction | 0.992 (0.980-1.004) | P= .19 | –0.8%/week |
| ≥1 blood culture obtained | Slope before blood culture restriction | 0.999 (0.998-1.001) | P= .42 | –0.1%/week |
|  | Level change upon blood culture restriction | 0.623 (0.563-0.689) | P< .001 | –37.7% (instant) |
|  | Slope during blood culture restriction | 1.009 (1.003-1.015) | P= .002 | +0.9%/week |
|  | Level change at end of blood culture restriction | 1.072 (0.957-1.201) | P= .23 | +7.2% (instant) |
|  | Slope after blood culture restriction | 0.987 (0.980-0.995) | P= .002 | –1.3%/week |
| ≥1 antimicrobial administered | Slope before blood culture restriction | 1.001 (1.000-1.003) | P= .08 | +0.1%/week |
|  | Level change upon blood culture restriction | 0.912 (0.865-0.962) | P< .001 | –8.8% (instant) |
|  | Slope during blood culture restriction | 1.003 (1.000-1.006) | P= .02 | +0.3%/week |
|  | Level change at end of blood culture restriction | 1.029 (0.955-1.109) | P= .45 | +2.9% (instant) |
|  | Slope after blood culture restriction | 0.993 (0.988-0.999) | P= .02 | –0.7%/week |
| Time from admission to first antimicrobial administered | Slope before blood culture restriction | 1.000 (0.998-1.001) | P= .73 | 0.0%/week |
|  | Level change upon blood culture restriction | 1.080 (1.001-1.165) | P= .046 | +8.0% (instant) |
|  | Slope during blood culture restriction | 0.999 (0.995-1.002) | P= .46 | –0.1%/week |
|  | Level change at end of blood culture restriction | 0.975 (0.905-1.049) | P= .49 | –2.5% (instant) |
|  | Slope after blood culture restriction | 1.002 (0.997-1.008) | P= .40 | +0.2%/week |
| Days of therapy with ≥1 antimicrobial administered | Slope before blood culture restriction | 1.000 (0.999-1.001) | P= .90 | 0.0%/week |
|  | Level change upon blood culture restriction | 0.954 (0.904-1.007) | P= .09 | –4.6% (instant) |
|  | Slope during blood culture restriction | 1.002 (0.999-1.005) | P= .25 | +0.2%/week |
|  | Level change at end of blood culture restriction | 1.050 (0.975-1.130) | P= .20 | +5.0% (instant) |
|  | Slope after blood culture restriction | 0.990 (0.985-0.994) | P< .001 | –1.0%/week |
| ≥1 broad-spectrum antimicrobial administered | Slope before blood culture restriction | 1.000 (0.999-1.001) | P= .91 | 0.0%/week |
|  | Level change upon blood culture restriction | 0.945 (0.899-0.993) | P= .03 | –5.5% (instant) |
|  | Slope during blood culture restriction | 1.003 (1.000-1.005) | P= .02 | +0.3%/week |
|  | Level change at end of blood culture restriction | 1.003 (0.944-1.067) | P= .91 | +0.3% (instant) |
|  | Slope after blood culture restriction | 0.994 (0.990-0.999) | P= .01 | –0.6%/week |
| Time from admission to first broad-spectrum antimicrobial administered | Slope before blood culture restriction | 1.000 (0.998-1.002) | P= .75 | 0.0%/week |
|  | Level change upon blood culture restriction | 1.075 (0.989-1.167) | P= .09 | +7.5% (instant) |
|  | Slope during blood culture restriction | 1.000 (0.997-1.003) | P= .87 | 0.0%/week |
|  | Level change at end of blood culture restriction | 1.011 (0.946-1.081) | P= .74 | +1.1% (instant) |
|  | Slope after blood culture restriction | 0.997 (0.992-1.002) | P= .29 | –0.3%/week |
| Days of therapy with ≥1 broad-spectrum antimicrobial administered | Slope before blood culture restriction | 1.000 (0.998-1.001) | P= .61 | 0.0%/week |
|  | Level change upon blood culture restriction | 0.957 (0.893-1.026) | P= .22 | –4.3% (instant) |
|  | Slope during blood culture restriction | 1.002 (0.998-1.006) | P= .29 | +0.2%/week |
|  | Level change at end of blood culture restriction | 1.028 (0.940-1.124) | P= .55 | +2.8% (instant) |
|  | Slope after blood culture restriction | 0.989 (0.984-0.995) | P< .001 | –1.1%/week |
| ≥1 non-broad-spectrum antimicrobial administered | Slope before blood culture restriction | 1.000 (0.999-1.002) | P= .54 | 0.0%/week |
|  | Level change upon blood culture restriction | 0.935 (0.883-0.992) | P= .02 | –6.5% (instant) |
|  | Slope during blood culture restriction | 1.002 (0.999-1.005) | P= .28 | +0.2%/week |
|  | Level change at end of blood culture restriction | 1.065 (1.000-1.135) | P= .050 | +6.5% (instant) |
|  | Slope after blood culture restriction | 0.993 (0.988-0.997) | P= .002 | –0.7%/week |
| Time from admission to first non-broad-spectrum antimicrobial administered | Slope before blood culture restriction | 0.999 (0.997-1.001) | P= .17 | –0.1%/week |
|  | Level change upon blood culture restriction | 1.079 (0.996-1.170) | P= .06 | +7.9% (instant) |
|  | Slope during blood culture restriction | 0.997 (0.993-1.001) | P= .18 | –0.3%/week |
|  | Level change at end of blood culture restriction | 1.040 (0.944-1.146) | P= .43 | +4.0% (instant) |
|  | Slope after blood culture restriction | 0.998 (0.992-1.005) | P= .64 | –0.2%/week |
| Days of therapy with ≥1 non-broad-spectrum antimicrobial administered | Slope before blood culture restriction | 1.001 (0.999-1.002) | P= .40 | +0.1%/week |
|  | Level change upon blood culture restriction | 0.941 (0.887-0.998) | P= .04 | –5.9% (instant) |
|  | Slope during blood culture restriction | 1.001 (0.998-1.005) | P= .49 | +0.1%/week |
|  | Level change at end of blood culture restriction | 1.077 (0.996-1.163) | P= .06 | +7.7% (instant) |
|  | Slope after blood culture restriction | 0.990 (0.985-0.995) | P< .001 | –1.0%/week |

Abbreviations: OR, odds ratio; IRR, incidence rate ratio.

Effect estimates reflect either odds ratios for binary outcomes or incidence rate ratios for count or continuous outcomes. The “Interpretation” column is intended to facilitate interpretation of the clinical significance of effect estimates.

#### **eTable 9.** Interpretation of Effect Estimates of Blood Culture Restriction on Primary and Secondary Outcomes During Hospitalizations with ≥1 Blood Culture Obtained at All 3 Hospital Sites Combined.

| **Outcome** | **Effect estimate** | **OR or IRR** | **P value** | **Interpretation (odds/rate)** |
| --- | --- | --- | --- | --- |
| In-hospital mortality or hospice discharge | Slope before blood culture restriction | 0.985 (0.980-0.990) | P< .001 | –1.5%/week |
|  | Level change upon blood culture restriction | 1.376 (1.101-1.721) | P= .005 | +37.6% (instant) |
|  | Slope during blood culture restriction | 0.996 (0.984-1.009) | P= .58 | –0.4%/week |
|  | Level change at end of blood culture restriction | 1.058 (0.838-1.337) | P= .63 | +5.8% (instant) |
|  | Slope after blood culture restriction | 0.980 (0.964-0.997) | P= .02 | –2.0%/week |
| Total 30-day revisits | Slope before blood culture restriction | 1.000 (0.998-1.003) | P= .69 | 0.0%/week |
|  | Level change upon blood culture restriction | 0.944 (0.832-1.072) | P= .37 | –5.6% (instant) |
|  | Slope during blood culture restriction | 1.005 (0.998-1.012) | P= .14 | +0.5%/week |
|  | Level change at end of blood culture restriction | 0.845 (0.725-0.984) | P= .03 | –15.5% (instant) |
|  | Slope after blood culture restriction | 1.000 (0.990-1.011) | P= .95 | 0.0%/week |
| 30-day inpatient readmission | Slope before blood culture restriction | 0.999 (0.996-1.001) | P= .27 | –0.1%/week |
|  | Level change upon blood culture restriction | 0.987 (0.887-1.097) | P= .80 | –1.3% (instant) |
|  | Slope during blood culture restriction | 1.004 (0.997-1.011) | P= .30 | +0.4%/week |
|  | Level change at end of blood culture restriction | 0.956 (0.822-1.111) | P= .56 | –4.4% (instant) |
|  | Slope after blood culture restriction | 0.995 (0.985-1.005) | P= .35 | –0.5%/week |
| 30-day medical observation unit revisits | Slope before blood culture restriction | 1.003 (0.998-1.008) | P= .27 | +0.3%/week |
|  | Level change upon blood culture restriction | 1.139 (0.877-1.479) | P= .33 | +13.9% (instant) |
|  | Slope during blood culture restriction | 0.994 (0.982-1.007) | P= .36 | –0.6%/week |
|  | Level change at end of blood culture restriction | 0.653 (0.446-0.956) | P= .03 | –34.7% (instant) |
|  | Slope after blood culture restriction | 1.030 (1.006-1.054) | P= .01 | +3.0%/week |
| 30-day emergency department revisits | Slope before blood culture restriction | 1.004 (0.999-1.008) | P= .13 | +0.4%/week |
|  | Level change upon blood culture restriction | 0.807 (0.589-1.106) | P= .18 | –19.3% (instant) |
|  | Slope during blood culture restriction | 1.013 (0.995-1.031) | P= .16 | +1.3%/week |
|  | Level change at end of blood culture restriction | 0.697 (0.495-0.983) | P= .04 | –30.3% (instant) |
|  | Slope after blood culture restriction | 1.003 (0.978-1.028) | P= .83 | +0.3%/week |
| Length of stay, days | Slope before blood culture restriction | 1.000 (0.998-1.001) | P= .47 | 0.0%/week |
|  | Level change upon blood culture restriction | 1.149 (1.084-1.217) | P< .001 | +14.9% (instant) |
|  | Slope during blood culture restriction | 1.000 (0.997-1.002) | P= .80 | 0.0%/week |
|  | Level change at end of blood culture restriction | 1.026 (0.964-1.092) | P= .42 | +2.6% (instant) |
|  | Slope after blood culture restriction | 0.991 (0.987-0.995) | P< .001 | –0.9%/week |
| Total number of blood cultures obtained | Slope before blood culture restriction | 1.001 (1.000-1.002) | P= .07 | +0.1%/week |
|  | Level change upon blood culture restriction | 0.508 (0.462-0.557) | P< .001 | –49.2% (instant) |
|  | Slope during blood culture restriction | 1.009 (1.001-1.017) | P= .02 | +0.9%/week |
|  | Level change at end of blood culture restriction | 1.501 (1.316-1.711) | P< .001 | +50.1% (instant) |
|  | Slope after blood culture restriction | 0.990 (0.982-0.998) | P= .02 | –1.0%/week |
| Number of calendar days with ≥1 blood culture obtained | Slope before blood culture restriction | 1.001 (1.000-1.001) | P= .04 | +0.1%/week |
|  | Level change upon blood culture restriction | 0.932 (0.882-0.984) | P= .01 | –6.8% (instant) |
|  | Slope during blood culture restriction | 1.001 (0.998-1.004) | P= .71 | +0.1%/week |
|  | Level change at end of blood culture restriction | 1.069 (1.016-1.124) | P= .010 | +6.9% (instant) |
|  | Slope after blood culture restriction | 0.996 (0.992-1.000) | P= .04 | –0.4%/week |
| Time from admission to first blood culture obtained, days | Slope before blood culture restriction | 0.998 (0.995-1.000) | P= .051 | –0.2%/week |
|  | Level change upon blood culture restriction | 1.722 (1.481-2.002) | P< .001 | +72.2% (instant) |
|  | Slope during blood culture restriction | 0.996 (0.988-1.005) | P= .40 | –0.4%/week |
|  | Level change at end of blood culture restriction | 0.998 (0.853-1.168) | P= .98 | –0.2% (instant) |
|  | Slope after blood culture restriction | 0.990 (0.979-1.001) | P= .08 | –1.0%/week |
| ≥1 positive blood culture obtained | Slope before blood culture restriction | 1.002 (0.999-1.005) | P= .28 | +0.2%/week |
|  | Level change upon blood culture restriction | 0.774 (0.647-0.926) | P= .005 | –22.6% (instant) |
|  | Slope during blood culture restriction | 1.017 (1.007-1.027) | P< .001 | +1.7%/week |
|  | Level change at end of blood culture restriction | 1.033 (0.873-1.221) | P= .71 | +3.3% (instant) |
|  | Slope after blood culture restriction | 0.989 (0.978-1.001) | P= .07 | –1.1%/week |
| Total number of positive blood cultures obtained | Slope before blood culture restriction | 1.003 (1.000-1.006) | P= .06 | +0.3%/week |
|  | Level change upon blood culture restriction | 0.674 (0.562-0.809) | P< .001 | –32.6% (instant) |
|  | Slope during blood culture restriction | 1.020 (1.011-1.029) | P< .001 | +2.0%/week |
|  | Level change at end of blood culture restriction | 1.114 (0.958-1.297) | P= .16 | +11.4% (instant) |
|  | Slope after blood culture restriction | 0.982 (0.971-0.994) | P= .002 | –1.8%/week |
| Number of calendar days with ≥1 positive blood culture obtained | Slope before blood culture restriction | 1.002 (0.999-1.005) | P= .15 | +0.2%/week |
|  | Level change upon blood culture restriction | 0.752 (0.642-0.882) | P< .001 | –24.8% (instant) |
|  | Slope during blood culture restriction | 1.018 (1.010-1.026) | P< .001 | +1.8%/week |
|  | Level change at end of blood culture restriction | 1.105 (0.944-1.294) | P= .22 | +10.5% (instant) |
|  | Slope after blood culture restriction | 0.982 (0.972-0.992) | P< .001 | –1.8%/week |
| ≥1 antimicrobial administered | Slope before blood culture restriction | 1.002 (0.999-1.005) | P= .22 | +0.2%/week |
|  | Level change upon blood culture restriction | 1.249 (1.034-1.509) | P= .02 | +24.9% (instant) |
|  | Slope during blood culture restriction | 1.003 (0.992-1.013) | P= .59 | +0.3%/week |
|  | Level change at end of blood culture restriction | 0.853 (0.674-1.080) | P= .19 | –14.7% (instant) |
|  | Slope after blood culture restriction | 0.997 (0.981-1.013) | P= .72 | –0.3%/week |
| Time from admission to first antimicrobial administered | Slope before blood culture restriction | 0.998 (0.996-1.000) | P= .10 | –0.2%/week |
|  | Level change upon blood culture restriction | 1.215 (1.080-1.367) | P= .001 | +21.5% (instant) |
|  | Slope during blood culture restriction | 0.998 (0.991-1.005) | P= .57 | -0.2%/week |
|  | Level change at end of blood culture restriction | 0.984 (0.851-1.138) | P= .83 | –1.6% (instant) |
|  | Slope after blood culture restriction | 0.997 (0.988-1.005) | P= .44 | –0.3%/week |
| Days of therapy with ≥1 antimicrobial administered | Slope before blood culture restriction | 1.000 (0.999-1.002) | P= .91 | +0.0%/week |
|  | Level change upon blood culture restriction | 1.152 (1.065-1.247) | P< .001 | +15.2% (instant) |
|  | Slope during blood culture restriction | 0.998 (0.994-1.002) | P= .39 | –0.2%/week |
|  | Level change at end of blood culture restriction | 1.024 (0.935-1.120) | P= .61 | +2.4% (instant) |
|  | Slope after blood culture restriction | 0.994 (0.988-1.000) | P= .054 | –0.6%/week |
| ≥1 broad-spectrum antimicrobial administered | Slope before blood culture restriction | 1.001 (0.998-1.003) | P= .66 | +0.1%/week |
|  | Level change upon blood culture restriction | 1.360 (1.145-1.615) | P< .001 | +36.0% (instant) |
|  | Slope during blood culture restriction | 0.998 (0.989-1.006) | P= .60 | –0.2%/week |
|  | Level change at end of blood culture restriction | 0.900 (0.791-1.023) | P= .11 | –10.0% (instant) |
|  | Slope after blood culture restriction | 1.003 (0.992-1.015) | P= .55 | +0.3%/week |
| Time from admission to first broad-spectrum antimicrobial administered | Slope before blood culture restriction | 1.000 (0.997-1.002) | P= .81 | 0.0%/week |
|  | Level change upon blood culture restriction | 1.192 (1.040-1.365) | P= .01 | +19.2% (instant) |
|  | Slope during blood culture restriction | 0.996 (0.989-1.004) | P= .37 | –0.4%/week |
|  | Level change at end of blood culture restriction | 1.026 (0.885-1.189) | P= .74 | +2.6% (instant) |
|  | Slope after blood culture restriction | 1.000 (0.991-1.009) | P= .99 | 0.0%/week |
| Days of therapy with ≥1 broad-spectrum antimicrobial administered | Slope before blood culture restriction | 1.000 (0.998-1.002) | P= .84 | 0.0%/week |
|  | Level change upon blood culture restriction | 1.187 (1.079-1.306) | P< .001 | +18.7% (instant) |
|  | Slope during blood culture restriction | 0.998 (0.992-1.003) | P= .35 | –0.2%/week |
|  | Level change at end of blood culture restriction | 1.005 (0.917-1.102) | P= .91 | +0.5% (instant) |
|  | Slope after blood culture restriction | 0.995 (0.989-1.002) | P= .16 | –0.5%/week |
| ≥1 non-broad-spectrum antimicrobial administered | Slope before blood culture restriction | 0.999 (0.997-1.001) | P= .22 | –0.1%/week |
|  | Level change upon blood culture restriction | 1.074 (0.974-1.184) | P= .15 | +7.4% (instant) |
|  | Slope during blood culture restriction | 1.001 (0.995-1.007) | P= .70 | +0.1%/week |
|  | Level change at end of blood culture restriction | 1.040 (0.915-1.183) | P= .55 | +4.0% (instant) |
|  | Slope after blood culture restriction | 0.990 (0.983-0.998) | P= .01 | –1.0%/week |
| Time from admission to first non-broad-spectrum antimicrobial administered | Slope before blood culture restriction | 0.997 (0.995-0.999) | P= .006 | –0.3%/week |
|  | Level change upon blood culture restriction | 1.298 (1.133-1.487) | P< .001 | +29.8% (instant) |
|  | Slope during blood culture restriction | 0.993 (0.986-1.001) | P= .07 | –0.7%/week |
|  | Level change at end of blood culture restriction | 1.119 (0.968-1.293) | P= .13 | +11.9% (instant) |
|  | Slope after blood culture restriction | 0.995 (0.985-1.005) | P= .32 | –0.5%/week |
| Days of therapy with ≥1 non-broad-spectrum antimicrobial administered | Slope before blood culture restriction | 1.001 (0.999-1.003) | P= .53 | +0.1%/week |
|  | Level change upon blood culture restriction | 1.111 (1.006-1.227) | P= .04 | +11.1% (instant) |
|  | Slope during blood culture restriction | 0.998 (0.992-1.004) | P= .46 | –0.2%/week |
|  | Level change at end of blood culture restriction | 1.055 (0.938-1.187) | P= .37 | +5.5% (instant) |
|  | Slope after blood culture restriction | 0.994 (0.987-1.002) | P= .13 | –0.6%/week |

Abbreviations: OR, odds ratio; IRR, incidence rate ratio.

Effect estimates reflect either odds ratios for binary outcomes or incidence rate ratios for count or continuous outcomes. The “Interpretation” column is intended to facilitate interpretation of the clinical significance of effect estimates.

#### **eTable 10.** Effect Estimates of Blood Culture Restriction on Primary and Secondary Outcomes for Hospitalizations at the Hospital of the University of Pennsylvania.

|  | **(Intercept)** | **Slope before blood culture restriction** | | **Level change upon blood culture restriction** | | **Slope during blood culture restriction** | | **Level change at end of blood culture restriction** | | **Slope after blood culture restriction** | |
| --- | --- | --- | --- | --- | --- | --- | --- | --- | --- | --- | --- |
| **Outcome** |  | **OR or IRR** | **P value** | **OR or IRR** | **P value** | **OR or IRR** | **P value** | **OR or IRR** | **P value** | **OR or IRR** | **P value** |
| **All hospitalizations** | | | | | | | | | | | |
| In-hospital mortality or hospice discharge | 0.043 (0.039-0.049) | 0.985 (0.981-0.989) | P< .001 | 1.202 (0.984-1.468) | P= .07 | 1.000 (0.989-1.010) | P= .97 | 1.126 (0.894-1.417) | P= .31 | 0.974 (0.958-0.990) | P= .001 |
| Total 30-day revisits | 0.352 (0.328-0.376) | 1.000 (0.997-1.002) | P= .82 | 1.026 (0.912-1.154) | P= .67 | 0.998 (0.992-1.004) | P= .48 | 1.058 (0.940-1.190) | P= .35 | 1.004 (0.996-1.013) | P= .30 |
| 30-day inpatient readmission | 0.271 (0.250-0.295) | 0.999 (0.996-1.002) | P= .42 | 1.006 (0.879-1.152) | P= .93 | 0.999 (0.993-1.005) | P= .73 | 1.122 (0.976-1.289) | P= .11 | 1.000 (0.991-1.010) | P= .98 |
| 30-day medical observation unit revisits | 0.025 (0.022-0.029) | 1.000 (0.996-1.004) | P= .99 | 1.197 (1.005-1.426) | P= .04 | 0.994 (0.984-1.005) | P= .27 | 0.846 (0.633-1.131) | P= .26 | 1.016 (0.998-1.034) | P= .08 |
| 30-day emergency department revisits | 0.055 (0.050-0.062) | 1.004 (1.000-1.007) | P= .07 | 1.032 (0.837-1.273) | P= .77 | 0.996 (0.987-1.005) | P= .41 | 0.892 (0.711-1.120) | P= .33 | 1.018 (1.002-1.035) | P= .03 |
| Length of stay, days | 7.175 (6.939-7.419) | 1.001 (1.000-1.002) | P= .22 | 0.984 (0.930-1.042) | P= .59 | 1.002 (0.999-1.005) | P= .26 | 1.009 (0.941-1.081) | P= .81 | 0.993 (0.988-0.998) | P= .002 |
| Overall blood culture positivity^a^ | 0.036 (0.032-0.040) | 1.002 (0.999-1.006) | P= .20 | 1.484 (1.215-1.814) | P< .001 | 1.012 (1.001-1.023) | P= .04 | 0.744 (0.587-0.942) | P= .01 | 0.984 (0.968-1.001) | P= .06 |
| ≥1 blood culture obtained^b^ | 0.438 (0.415-0.462) | 0.999 (0.997-1.001) | P= .20 | 0.734 (0.668-0.805) | P< .001 | 1.004 (0.998-1.009) | P= .19 | 1.026 (0.924-1.139) | P= .64 | 0.996 (0.988-1.003) | P= .25 |
| ≥1 antimicrobial administered | 1.232 (1.156-1.314) | 1.001 (0.999-1.003) | P= .19 | 0.934 (0.880-0.992) | P= .03 | 1.003 (0.999-1.006) | P= .11 | 1.003 (0.919-1.094) | P= .95 | 0.995 (0.989-1.002) | P= .13 |
| Time from admission to first antimicrobial administered, days | 1.229 (1.170-1.291) | 1.001 (0.999-1.002) | P= .47 | 1.105 (1.021-1.196) | P= .01 | 0.997 (0.993-1.001) | P= .13 | 0.972 (0.874-1.081) | P= .60 | 1.007 (0.999-1.015) | P= .09 |
| Days of therapy with ≥1 antimicrobial administered | 3.728 (3.558-3.907) | 1.000 (0.999-1.001) | P= .99 | 0.981 (0.912-1.056) | P= .61 | 1.000 (0.996-1.005) | P= .86 | 1.062 (0.961-1.173) | P= .24 | 0.990 (0.984-0.996) | P= .002 |
| ≥1 broad-spectrum antimicrobial administered | 0.594 (0.569-0.621) | 1.000 (0.999-1.002) | P= .84 | 0.956 (0.898-1.017) | P= .15 | 1.003 (1.000-1.006) | P= .02 | 0.980 (0.910-1.055) | P= .59 | 0.993 (0.988-0.999) | P= .02 |
| Time from admission to first broad-spectrum antimicrobial administered, days | 1.787 (1.674-1.907) | 1.001 (0.998-1.003) | P= .62 | 1.082 (0.981-1.193) | P= .12 | 0.999 (0.995-1.003) | P= .65 | 0.990 (0.896-1.094) | P= .85 | 1.001 (0.993-1.008) | P= .88 |
| Days of therapy with ≥1 broad-spectrum antimicrobial administered | 2.557 (2.409-2.714) | 1.000 (0.998-1.002) | P= .91 | 0.963 (0.881-1.052) | P= .40 | 1.001 (0.996-1.007) | P= .67 | 1.039 (0.930-1.162) | P= .50 | 0.990 (0.983-0.997) | P= .005 |
| ≥1 non-broad-spectrum antimicrobial administered | 0.758 (0.710-0.810) | 1.000 (0.998-1.002) | P= .72 | 0.975 (0.916-1.037) | P= .41 | 1.001 (0.998-1.005) | P= .36 | 1.038 (0.971-1.109) | P= .27 | 0.993 (0.988-0.998) | P= .006 |
| Time from admission to first non-broad-spectrum antimicrobial administered, days | 1.928 (1.807-2.058) | 1.001 (0.998-1.003) | P= .54 | 1.113 (1.004-1.234) | P= .04 | 0.994 (0.988-0.999) | P= .03 | 1.061 (0.922-1.221) | P= .41 | 1.004 (0.995-1.014) | P= .37 |
| Days of therapy with ≥1 non-broad-spectrum antimicrobial administered | 2.026 (1.918-2.140) | 1.000 (0.998-1.002) | P= .89 | 0.970 (0.894-1.053) | P= .47 | 1.000 (0.994-1.006) | P= .96 | 1.083 (0.965-1.214) | P= .17 | 0.990 (0.983-0.997) | P= .004 |
| **Hospitalizations with ≥1 blood culture obtained (≥1-BC hospitalizations)** | | | | | | | | | | | |
| In-hospital mortality or hospice discharge | 0.094 (0.081-0.110) | 0.985 (0.980-0.990) | P< .001 | 1.583 (1.188-2.109) | P= .002 | 0.986 (0.970-1.002) | P= .09 | 1.262 (0.913-1.744) | P= .16 | 0.988 (0.965-1.012) | P= .34 |
| Total 30-day revisits | 0.348 (0.320-0.380) | 1.000 (0.997-1.003) | P= .77 | 1.043 (0.891-1.221) | P= .60 | 1.001 (0.993-1.010) | P= .78 | 0.850 (0.676-1.070) | P= .17 | 1.005 (0.990-1.021) | P= .50 |
| 30-day inpatient readmission | 0.252 (0.231-0.276) | 0.998 (0.995-1.001) | P= .13 | 0.995 (0.875-1.132) | P= .94 | 1.006 (0.997-1.015) | P= .22 | 0.887 (0.697-1.129) | P= .33 | 0.993 (0.978-1.009) | P= .39 |
| 30-day medical observation unit revisits | 0.028 (0.023-0.035) | 1.002 (0.996-1.009) | P= .52 | 0.891 (0.623-1.274) | P= .53 | 1.011 (0.993-1.030) | P= .22 | 0.465 (0.287-0.755) | P= .002 | 1.025 (0.994-1.057) | P= .12 |
| 30-day emergency department revisits | 0.068 (0.057-0.081) | 1.004 (0.998-1.010) | P= .20 | 1.241 (0.837-1.840) | P= .28 | 0.986 (0.965-1.006) | P= .17 | 0.923 (0.594-1.436) | P= .72 | 1.035 (1.000-1.070) | P= .048 |
| Length of stay, days | 13.503 (12.842-14.199) | 1.000 (0.998-1.001) | P= .88 | 1.081 (0.997-1.172) | P= .06 | 1.001 (0.997-1.005) | P= .61 | 1.063 (0.963-1.173) | P= .23 | 0.988 (0.981-0.994) | P< .001 |
| Total number of blood cultures obtained^c^ | 3.294 (3.187-3.404) | 1.001 (1.000-1.002) | P= .09 | 0.482 (0.435-0.534) | P< .001 | 1.012 (1.003-1.020) | P= .006 | 1.531 (1.312-1.786) | P< .001 | 0.984 (0.975-0.994) | P< .001 |
| Number of calendar days with ≥1 blood culture obtained^c^ | 1.840 (1.782-1.900) | 1.001 (1.000-1.002) | P= .02 | 0.887 (0.823-0.956) | P= .002 | 1.002 (0.998-1.006) | P= .39 | 1.102 (1.013-1.198) | P= .02 | 0.993 (0.988-0.999) | P= .02 |
| Time from admission to first blood culture obtained, days^c^ | 1.652 (1.504-1.813) | 0.998 (0.995-1.000) | P= .09 | 1.613 (1.354-1.921) | P< .001 | 0.998 (0.988-1.007) | P= .62 | 1.062 (0.873-1.292) | P= .55 | 0.986 (0.973-0.999) | P= .04 |
| ≥1 positive blood culture obtained^c^ | 0.101 (0.090-0.114) | 1.001 (0.997-1.005) | P= .60 | 0.813 (0.658-1.006) | P= .06 | 1.019 (1.007-1.030) | P= .001 | 1.066 (0.878-1.294) | P= .52 | 0.982 (0.968-0.996) | P= .01 |
| Total number of positive blood cultures obtained^c^ | 0.114 (0.102-0.127) | 1.003 (0.999-1.007) | P= .09 | 0.697 (0.558-0.871) | P= .002 | 1.024 (1.011-1.036) | P< .001 | 1.151 (0.901-1.470) | P= .26 | 0.969 (0.953-0.986) | P< .001 |
| Number of calendar days with ≥1 positive blood culture obtained^c^ | 0.107 (0.096-0.119) | 1.002 (0.999-1.006) | P= .24 | 0.761 (0.617-0.939) | P= .01 | 1.020 (1.008-1.032) | P< .001 | 1.210 (0.944-1.551) | P= .13 | 0.971 (0.955-0.988) | P< .001 |
| ≥1 antimicrobial administered | 7.535 (6.568-8.645) | 1.003 (0.999-1.007) | P= .16 | 1.204 (0.954-1.519) | P= .12 | 0.999 (0.986-1.012) | P= .87 | 0.960 (0.722-1.278) | P= .78 | 1.000 (0.981-1.020) | P= .97 |
| Time from admission to first antimicrobial administered, days | 1.241 (1.144-1.347) | 1.000 (0.997-1.003) | P= .92 | 1.161 (0.995-1.354) | P= .06 | 0.997 (0.987-1.007) | P= .59 | 1.033 (0.828-1.288) | P= .77 | 0.999 (0.986-1.013) | P= .93 |
| Days of therapy with ≥1 antimicrobial administered | 9.113 (8.640-9.613) | 1.000 (0.998-1.002) | P= .90 | 1.105 (1.009-1.211) | P= .03 | 0.999 (0.993-1.004) | P= .59 | 1.079 (0.959-1.215) | P= .21 | 0.991 (0.983-0.998) | P= .01 |
| ≥1 broad-spectrum antimicrobial administered | 3.463 (3.109-3.858) | 1.002 (0.998-1.005) | P= .31 | 1.176 (0.936-1.479) | P= .16 | 0.999 (0.988-1.011) | P= .89 | 0.991 (0.837-1.173) | P= .92 | 1.000 (0.985-1.015) | P= 1.00 |
| Time from admission to first broad-spectrum antimicrobial administered, days | 1.788 (1.645-1.942) | 1.000 (0.998-1.003) | P= .76 | 1.160 (0.991-1.358) | P= .06 | 0.998 (0.988-1.007) | P= .63 | 1.012 (0.840-1.218) | P= .90 | 1.000 (0.988-1.012) | P= .99 |
| Days of therapy with ≥1 broad-spectrum antimicrobial administered | 6.805 (6.384-7.254) | 1.000 (0.998-1.002) | P= .74 | 1.094 (0.976-1.226) | P= .12 | 0.998 (0.992-1.005) | P= .61 | 1.066 (0.945-1.202) | P= .30 | 0.992 (0.984-1.000) | P= .04 |
| ≥1 non-broad-spectrum antimicrobial administered | 2.214 (2.016-2.431) | 0.996 (0.993-0.999) | P= .006 | 1.181 (1.046-1.335) | P= .007 | 0.999 (0.992-1.005) | P= .64 | 1.044 (0.896-1.216) | P= .58 | 0.990 (0.979-1.001) | P= .08 |
| Time from admission to first non-broad-spectrum antimicrobial administered, days | 2.573 (2.365-2.799) | 1.000 (0.998-1.003) | P= .76 | 1.187 (0.994-1.417) | P= .06 | 0.992 (0.980-1.003) | P= .15 | 1.202 (0.955-1.513) | P= .12 | 0.998 (0.983-1.013) | P= .77 |
| Days of therapy with ≥1 non-broad-spectrum antimicrobial administered | 4.662 (4.345-5.003) | 1.000 (0.998-1.002) | P= .97 | 1.095 (0.979-1.226) | P= .11 | 0.998 (0.990-1.006) | P= .67 | 1.096 (0.930-1.293) | P= .27 | 0.990 (0.980-1.000) | P= .06 |

Abbreviations: OR, odds ratio; IRR, incidence rate ratio.

The pre-restriction period was June 26, 2023 to June 25, 2024; the restriction period was June 26, 2024 to December 23, 2024; and the post-restriction period was December 24, 2024 to June 25, 2025. The total 30-day readmission outcome is comprised of all readmissions, including inpatient readmissions, medical observation unit revisits, and emergency department revisits. Effects estimates for binary outcomes (in-hospital mortality or hospice discharge, ≥1 blood culture obtained, ≥1 positive blood culture, ≥1 antimicrobial administered, ≥1 broad-spectrum antimicrobial administered, and ≥1 non-broad-spectrum antimicrobial administered) were odds ratios. Effect estimates for count and continuous outcomes (30-day inpatient readmission outcomes, length of stay, total numbers of blood cultures and positive blood cultures, number of calendar days with ≥1 blood culture or ≥1 positive blood culture obtained, time from admission to first blood culture obtained, time from admission to first antimicrobial administered including broad and non-broad-spectrum, and days of therapy with ≥1 antimicrobial administered including broad and non-broad-spectrum), were incidence rate ratios.

^a^ Effect estimates for the overall culture positivity rate reflect the entirety of the Hospital of the University of Pennsylvania rather than averaged measures for individual hospitalizations, and thus are not shown for the hospitalizations with ≥1 blood culture obtained subgroup (≥1-BC hospitalizations).

^b^ Effect estimates were not calculated for ≥1-BC hospitalizations as this subgroup had ≥1 blood culture obtained by definition.

^c^ Effect estimates were only calculated for ≥1-BC hospitalizations.

#### **eTable 11.** Effect Estimates of Blood Culture Restriction on Primary and Secondary Outcomes for Hospitalizations at Penn Presbyterian Medical Center.

|  | **(Intercept)** | **Slope before blood culture restriction** | | **Level change upon blood culture restriction** | | **Slope during blood culture restriction** | | **Level change at end of blood culture restriction** | | **Slope after blood culture restriction** | |
| --- | --- | --- | --- | --- | --- | --- | --- | --- | --- | --- | --- |
| **Outcome** |  | **OR or IRR** | **P value** | **OR or IRR** | **P value** | **OR or IRR** | **P value** | **OR or IRR** | **P value** | **OR or IRR** | **P value** |
| **All hospitalizations** | | | | | | | | | | | |
| In-hospital mortality or hospice discharge | 0.045 (0.038-0.054) | 0.988 (0.983-0.993) | P< .001 | 0.684 (0.470-0.997) | P= .048 | 1.026 (1.000-1.053) | P= .051 | 0.682 (0.410-1.133) | P= .14 | 0.965 (0.933-0.998) | P= .04 |
| Total 30-day revisits | 0.406 (0.353-0.466) | 0.996 (0.992-1.000) | P= .03 | 1.173 (0.974-1.413) | P= .09 | 0.995 (0.985-1.005) | P= .34 | 0.775 (0.590-1.017) | P= .07 | 1.036 (1.013-1.061) | P= .002 |
| 30-day inpatient readmission | 0.279 (0.227-0.342) | 0.993 (0.988-0.998) | P= .008 | 1.202 (0.919-1.571) | P= .18 | 0.995 (0.980-1.010) | P= .51 | 0.772 (0.530-1.126) | P= .18 | 1.044 (1.012-1.076) | P= .006 |
| 30-day medical observation unit revisits | 0.023 (0.019-0.028) | 1.003 (0.997-1.009) | P= .36 | 1.469 (1.137-1.900) | P= .003 | 0.978 (0.962-0.995) | P= .010 | 1.179 (0.782-1.777) | P= .43 | 1.017 (0.991-1.045) | P= .21 |
| 30-day emergency department revisits | 0.105 (0.091-0.122) | 1.000 (0.996-1.005) | P= .86 | 1.041 (0.847-1.280) | P= .70 | 1.000 (0.990-1.009) | P= .95 | 0.731 (0.587-0.911) | P= .005 | 1.019 (1.001-1.037) | P= .04 |
| Length of stay, days | 6.725 (6.481-6.979) | 0.999 (0.998-1.001) | P= .37 | 0.988 (0.933-1.046) | P= .67 | 1.002 (1.000-1.005) | P= .10 | 1.047 (0.958-1.145) | P= .31 | 0.990 (0.984-0.996) | P= .001 |
| Overall blood culture positivity^a^ | 0.051 (0.042-0.062) | 1.002 (0.996-1.008) | P= .45 | 1.483 (1.067-2.061) | P= .02 | 1.017 (1.000-1.034) | P= .051 | 0.549 (0.405-0.746) | P< .001 | 1.002 (0.978-1.027) | P= .85 |
| ≥1 blood culture obtained^b^ | 0.418 (0.387-0.452) | 1.001 (0.998-1.003) | P= .71 | 0.527 (0.447-0.620) | P< .001 | 1.012 (1.002-1.021) | P= .01 | 1.230 (1.013-1.493) | P= .04 | 0.980 (0.967-0.992) | P= .002 |
| ≥1 antimicrobial administered | 1.534 (1.435-1.640) | 1.001 (0.999-1.003) | P= .51 | 0.875 (0.793-0.965) | P= .007 | 1.002 (0.997-1.007) | P= .35 | 1.194 (1.007-1.415) | P= .04 | 0.987 (0.976-0.997) | P= .01 |
| Time from admission to first antimicrobial administered, days | 0.956 (0.870-1.050) | 1.000 (0.997-1.003) | P= .97 | 1.019 (0.866-1.198) | P= .82 | 1.004 (0.996-1.012) | P= .37 | 0.887 (0.765-1.030) | P= .12 | 0.997 (0.985-1.008) | P= .58 |
| Days of therapy with ≥1 antimicrobial administered | 3.085 (2.929-3.250) | 0.999 (0.997-1.001) | P= .51 | 0.937 (0.840-1.045) | P= .24 | 1.003 (0.999-1.008) | P= .15 | 1.122 (1.010-1.246) | P= .03 | 0.984 (0.976-0.991) | P< .001 |
| ≥1 broad-spectrum antimicrobial administered | 0.694 (0.641-0.751) | 1.000 (0.998-1.003) | P= .85 | 0.900 (0.796-1.019) | P= .10 | 1.001 (0.996-1.007) | P= .66 | 1.165 (1.020-1.330) | P= .02 | 0.988 (0.979-0.998) | P= .02 |
| Time from admission to first broad-spectrum antimicrobial administered, days | 1.254 (1.108-1.419) | 0.998 (0.994-1.002) | P= .32 | 1.080 (0.910-1.282) | P= .38 | 1.004 (0.997-1.011) | P= .23 | 1.018 (0.822-1.261) | P= .87 | 0.985 (0.969-1.001) | P= .07 |
| Days of therapy with ≥1 broad-spectrum antimicrobial administered | 1.915 (1.798-2.040) | 0.999 (0.997-1.002) | P= .48 | 0.937 (0.836-1.049) | P= .26 | 1.004 (0.999-1.009) | P= .13 | 1.078 (0.960-1.209) | P= .20 | 0.984 (0.975-0.992) | P< .001 |
| ≥1 non-broad-spectrum antimicrobial administered | 0.979 (0.920-1.042) | 1.000 (0.998-1.002) | P= .87 | 0.952 (0.864-1.049) | P= .32 | 0.999 (0.994-1.004) | P= .62 | 1.194 (1.035-1.377) | P= .01 | 0.991 (0.982-1.001) | P= .07 |
| Time from admission to first non-broad-spectrum antimicrobial administered, days | 1.725 (1.585-1.878) | 0.997 (0.993-1.000) | P= .04 | 1.009 (0.869-1.171) | P= .91 | 1.005 (0.998-1.013) | P= .13 | 0.942 (0.786-1.130) | P= .52 | 0.990 (0.978-1.002) | P= .09 |
| Days of therapy with ≥1 non-broad-spectrum antimicrobial administered | 1.768 (1.675-1.867) | 1.000 (0.998-1.002) | P= .78 | 0.966 (0.840-1.110) | P= .62 | 1.001 (0.994-1.007) | P= .84 | 1.188 (1.057-1.334) | P= .004 | 0.987 (0.977-0.996) | P= .007 |
| **Hospitalizations with ≥1 blood culture obtained (≥1-BC hospitalizations)** | | | | | | | | | | | |
| In-hospital mortality or hospice discharge | 0.087 (0.065-0.116) | 0.986 (0.977-0.996) | P= .004 | 0.774 (0.442-1.355) | P= .37 | 1.034 (1.005-1.063) | P= .02 | 0.606 (0.374-0.982) | P= .04 | 0.959 (0.924-0.994) | P= .02 |
| Total 30-day revisits | 0.338 (0.288-0.397) | 1.002 (0.997-1.006) | P= .51 | 0.855 (0.682-1.072) | P= .17 | 1.007 (0.995-1.018) | P= .27 | 0.947 (0.730-1.230) | P= .68 | 0.996 (0.978-1.015) | P= .66 |
| 30-day inpatient readmission | 0.194 (0.163-0.231) | 1.001 (0.996-1.007) | P= .60 | 0.959 (0.762-1.206) | P= .72 | 0.996 (0.984-1.009) | P= .58 | 1.138 (0.842-1.539) | P= .40 | 1.008 (0.989-1.029) | P= .41 |
| 30-day medical observation unit revisits | 0.022 (0.017-0.030) | 1.009 (0.997-1.021) | P= .13 | 1.141 (0.604-2.155) | P= .68 | 0.982 (0.944-1.022) | P= .37 | 0.896 (0.358-2.243) | P= .81 | 1.034 (0.979-1.092) | P= .23 |
| 30-day emergency department revisits | 0.123 (0.093-0.163) | 1.000 (0.992-1.009) | P= .96 | 0.636 (0.414-0.978) | P= .04 | 1.030 (1.008-1.052) | P= .006 | 0.722 (0.458-1.137) | P= .16 | 0.966 (0.934-0.998) | P= .04 |
| Length of stay, days | 11.666 (11.061-12.304) | 0.998 (0.996-1.000) | P= .03 | 1.240 (1.109-1.386) | P< .001 | 0.999 (0.995-1.004) | P= .79 | 1.008 (0.931-1.091) | P= .85 | 0.992 (0.985-0.999) | P= .02 |
| Total number of blood cultures obtained^c^ | 2.902 (2.760-3.051) | 1.000 (0.999-1.002) | P= .93 | 0.534 (0.461-0.619) | P< .001 | 1.008 (0.998-1.018) | P= .11 | 1.518 (1.301-1.771) | P< .001 | 0.993 (0.982-1.004) | P= .18 |
| Number of calendar days with ≥1 blood culture obtained^c^ | 1.635 (1.563-1.710) | 1.000 (0.998-1.001) | P= .57 | 0.997 (0.892-1.114) | P= .95 | 1.000 (0.995-1.005) | P= .98 | 1.048 (0.987-1.113) | P= .13 | 0.998 (0.992-1.005) | P= .57 |
| Time from admission to first blood culture obtained, days^c^ | 1.077 (0.943-1.230) | 0.999 (0.994-1.005) | P= .82 | 1.458 (1.094-1.943) | P= .01 | 1.009 (0.995-1.023) | P= .20 | 0.813 (0.587-1.125) | P= .21 | 0.981 (0.960-1.003) | P= .09 |
| ≥1 positive blood culture obtained^c^ | 0.135 (0.112-0.164) | 1.001 (0.995-1.007) | P= .69 | 0.764 (0.533-1.097) | P= .14 | 1.024 (1.005-1.044) | P= .01 | 0.896 (0.652-1.231) | P= .50 | 0.992 (0.968-1.016) | P= .51 |
| Total number of positive blood cultures obtained^c^ | 0.141 (0.117-0.170) | 1.002 (0.996-1.008) | P= .47 | 0.761 (0.530-1.093) | P= .14 | 1.025 (1.006-1.043) | P= .009 | 0.866 (0.645-1.163) | P= .34 | 0.994 (0.971-1.018) | P= .61 |
| Number of calendar days with ≥1 positive blood culture obtained^c^ | 0.129 (0.108-0.153) | 1.002 (0.997-1.007) | P= .47 | 0.742 (0.533-1.034) | P= .08 | 1.026 (1.009-1.043) | P= .002 | 0.875 (0.660-1.161) | P= .36 | 0.989 (0.968-1.010) | P= .30 |
| ≥1 antimicrobial administered | 6.717 (5.768-7.821) | 0.997 (0.991-1.002) | P= .27 | 1.250 (0.933-1.674) | P= .14 | 1.010 (0.996-1.023) | P= .16 | 0.909 (0.642-1.287) | P= .59 | 0.980 (0.956-1.004) | P= .10 |
| Time from admission to first antimicrobial administered, days | 1.023 (0.886-1.180) | 0.997 (0.992-1.002) | P= .20 | 1.125 (0.836-1.513) | P= .44 | 1.010 (0.996-1.025) | P= .16 | 0.703 (0.553-0.893) | P= .004 | 0.990 (0.970-1.010) | P= .33 |
| Days of therapy with ≥1 antimicrobial administered | 6.836 (6.446-7.250) | 0.999 (0.996-1.001) | P= .27 | 1.228 (1.040-1.451) | P= .02 | 0.998 (0.991-1.005) | P= .58 | 1.036 (0.941-1.141) | P= .47 | 0.993 (0.984-1.003) | P= .19 |
| ≥1 broad-spectrum antimicrobial administered | 2.576 (2.243-2.958) | 0.999 (0.995-1.003) | P= .68 | 1.488 (1.195-1.854) | P< .001 | 0.996 (0.985-1.006) | P= .42 | 1.018 (0.811-1.278) | P= .88 | 0.998 (0.982-1.014) | P= .77 |
| Time from admission to first broad-spectrum antimicrobial administered, days | 1.470 (1.248-1.730) | 0.997 (0.991-1.003) | P= .27 | 1.214 (0.898-1.641) | P= .21 | 1.002 (0.988-1.017) | P= .75 | 1.040 (0.742-1.458) | P= .82 | 0.985 (0.962-1.009) | P= .22 |
| Days of therapy with ≥1 broad-spectrum antimicrobial administered | 4.692 (4.414-4.987) | 0.998 (0.996-1.001) | P= .22 | 1.294 (1.097-1.527) | P= .002 | 0.998 (0.992-1.005) | P= .63 | 0.969 (0.881-1.066) | P= .52 | 0.995 (0.985-1.005) | P= .32 |
| ≥1 non-broad-spectrum antimicrobial administered | 1.917 (1.713-2.146) | 0.998 (0.994-1.002) | P= .28 | 1.102 (0.872-1.393) | P= .42 | 1.004 (0.993-1.014) | P= .47 | 1.098 (0.875-1.379) | P= .42 | 0.981 (0.963-0.998) | P= .03 |
| Time from admission to first non-broad-spectrum antimicrobial administered, days | 2.672 (2.368-3.015) | 0.991 (0.986-0.995) | P< .001 | 1.369 (1.056-1.775) | P= .02 | 1.005 (0.992-1.017) | P= .47 | 0.886 (0.675-1.162) | P= .38 | 0.985 (0.965-1.006) | P= .16 |
| Days of therapy with ≥1 non-broad-spectrum antimicrobial administered | 3.434 (3.159-3.733) | 0.999 (0.996-1.003) | P= .72 | 1.190 (0.921-1.539) | P= .18 | 0.995 (0.983-1.007) | P= .45 | 1.167 (0.996-1.366) | P= .06 | 0.993 (0.978-1.009) | P= .39 |

Abbreviations: OR, odds ratio; IRR, incidence rate ratio.

The pre-restriction period was June 26, 2023 to June 25, 2024; the restriction period was June 26, 2024 to December 23, 2024; and the post-restriction period was December 24, 2024 to June 25, 2025. The total 30-day readmission outcome is comprised of all readmissions, including inpatient readmissions, medical observation unit revisits, and emergency department revisits. Effects estimates for binary outcomes (in-hospital mortality or hospice discharge, ≥1 blood culture obtained, ≥1 positive blood culture, ≥1 antimicrobial administered, ≥1 broad-spectrum antimicrobial administered, and ≥1 non-broad-spectrum antimicrobial administered) were odds ratios. Effect estimates for count and continuous outcomes (30-day inpatient readmission outcomes, length of stay, total numbers of blood cultures and positive blood cultures, number of calendar days with ≥1 blood culture or ≥1 positive blood culture obtained, time from admission to first blood culture obtained, time from admission to first antimicrobial administered including broad and non-broad-spectrum, and days of therapy with ≥1 antimicrobial administered including broad and non-broad-spectrum), were incidence rate ratios.

^a^ Effect estimates for the overall culture positivity rate reflect the entirety of Penn Presbyterian Medical Center rather than averaged measures for individual hospitalizations, and thus are not shown for the hospitalizations with ≥1 blood culture obtained subgroup (≥1-BC hospitalizations).

^b^ Effect estimates were not calculated for ≥1-BC hospitalizations as this subgroup had ≥1 blood culture obtained by definition.

^c^ Effect estimates were only calculated for ≥1-BC hospitalizations.

#### **eTable 12.** Effect Estimates of Blood Culture Restriction on Primary and Secondary Outcomes for Hospitalizations at Pennsylvania Hospital.

|  | **(Intercept)** | **Slope before blood culture restriction** | | **Level change upon blood culture restriction** | | **Slope during blood culture restriction** | | **Level change at end of blood culture restriction** | | **Slope after blood culture restriction** | |
| --- | --- | --- | --- | --- | --- | --- | --- | --- | --- | --- | --- |
| **Outcome** |  | **OR or IRR** | **P value** | **OR or IRR** | **P value** | **OR or IRR** | **P value** | **OR or IRR** | **P value** | **OR or IRR** | **P value** |
| **All hospitalizations** | | | | | | | | | | | |
| In-hospital mortality or hospice discharge | 0.011 (0.008-0.016) | 0.996 (0.983-1.009) | P= .52 | 0.894 (0.532-1.501) | P= .67 | 0.986 (0.966-1.006) | P= .16 | 1.918 (1.234-2.981) | P= .004 | 0.976 (0.941-1.011) | P= .17 |
| Total 30-day revisits | 0.355 (0.328-0.385) | 1.002 (0.999-1.004) | P= .14 | 0.853 (0.762-0.956) | P= .006 | 1.002 (0.996-1.008) | P= .50 | 0.879 (0.763-1.013) | P= .08 | 0.999 (0.989-1.009) | P= .84 |
| 30-day inpatient readmission | 0.161 (0.146-0.178) | 1.004 (1.001-1.007) | P= .01 | 0.772 (0.656-0.909) | P= .002 | 0.998 (0.990-1.007) | P= .65 | 0.991 (0.835-1.177) | P= .92 | 1.006 (0.994-1.019) | P= .34 |
| 30-day medical observation unit revisits | 0.041 (0.035-0.048) | 1.001 (0.995-1.006) | P= .76 | 1.189 (0.930-1.521) | P= .17 | 0.986 (0.972-1.001) | P= .07 | 0.913 (0.565-1.476) | P= .71 | 1.012 (0.981-1.045) | P= .44 |
| 30-day emergency department revisits | 0.153 (0.136-0.173) | 0.999 (0.995-1.003) | P= .72 | 0.871 (0.708-1.073) | P= .19 | 1.011 (1.001-1.021) | P= .03 | 0.770 (0.617-0.961) | P= .02 | 0.988 (0.973-1.003) | P= .10 |
| Length of stay, days | 4.591 (4.414-4.775) | 1.001 (0.999-1.002) | P= .41 | 0.975 (0.925-1.027) | P= .34 | 1.003 (1.000-1.006) | P= .06 | 1.008 (0.928-1.095) | P= .85 | 0.993 (0.987-0.998) | P= .009 |
| Overall blood culture positivity^a^ | 0.095 (0.081-0.111) | 1.001 (0.996-1.007) | P= .67 | 1.256 (0.780-2.023) | P= .35 | 0.999 (0.974-1.025) | P= .95 | 1.027 (0.691-1.527) | P= .90 | 0.998 (0.965-1.031) | P= .89 |
| ≥1 blood culture obtained^b^ | 0.282 (0.256-0.310) | 0.999 (0.996-1.003) | P= .77 | 0.463 (0.375-0.570) | P< .001 | 1.024 (1.013-1.034) | P< .001 | 1.030 (0.872-1.218) | P= .73 | 0.971 (0.957-0.985) | P< .001 |
| ≥1 antimicrobial administered | 0.878 (0.821-0.939) | 1.002 (1.000-1.005) | P= .08 | 0.888 (0.784-1.005) | P= .06 | 1.006 (1.000-1.011) | P= .06 | 0.955 (0.871-1.047) | P= .32 | 0.995 (0.988-1.002) | P= .19 |
| Time from admission to first antimicrobial administered, days | 0.867 (0.775-0.969) | 0.997 (0.993-1.000) | P= .07 | 1.051 (0.915-1.207) | P= .48 | 0.999 (0.993-1.005) | P= .68 | 1.167 (0.999-1.362) | P= .051 | 0.989 (0.978-1.001) | P= .06 |
| Days of therapy with ≥1 antimicrobial administered | 1.640 (1.531-1.756) | 1.003 (1.001-1.005) | P= .02 | 0.857 (0.765-0.959) | P= .007 | 1.006 (1.000-1.011) | P= .04 | 0.927 (0.843-1.020) | P= .12 | 0.994 (0.987-1.002) | P= .14 |
| ≥1 broad-spectrum antimicrobial administered | 0.298 (0.276-0.322) | 1.001 (0.998-1.003) | P= .65 | 0.945 (0.842-1.060) | P= .33 | 1.003 (0.997-1.009) | P= .28 | 0.901 (0.802-1.011) | P= .08 | 1.002 (0.993-1.010) | P= .73 |
| Time from admission to first broad-spectrum antimicrobial administered, days | 1.192 (1.027-1.384) | 0.999 (0.993-1.005) | P= .71 | 1.015 (0.790-1.305) | P= .91 | 0.994 (0.984-1.004) | P= .24 | 1.206 (1.000-1.455) | P= .050 | 1.000 (0.983-1.016) | P= .98 |
| Days of therapy with ≥1 broad-spectrum antimicrobial administered | 0.816 (0.743-0.897) | 1.001 (0.998-1.004) | P= .52 | 0.940 (0.806-1.096) | P= .43 | 1.006 (0.998-1.013) | P= .13 | 0.931 (0.807-1.075) | P= .33 | 0.993 (0.982-1.003) | P= .17 |
| ≥1 non-broad-spectrum antimicrobial administered | 0.636 (0.591-0.684) | 1.003 (1.000-1.005) | P= .06 | 0.832 (0.721-0.961) | P= .01 | 1.006 (0.998-1.013) | P= .13 | 1.005 (0.893-1.132) | P= .93 | 0.992 (0.983-1.001) | P= .10 |
| Time from admission to first non-broad-spectrum antimicrobial administered, days | 1.150 (1.016-1.301) | 0.996 (0.993-1.000) | P= .050 | 0.973 (0.855-1.107) | P= .67 | 1.001 (0.995-1.007) | P= .78 | 1.170 (0.985-1.389) | P= .07 | 0.986 (0.974-0.997) | P= .01 |
| Days of therapy with ≥1 non-broad-spectrum antimicrobial administered | 1.058 (0.982-1.140) | 1.004 (1.001-1.007) | P= .008 | 0.804 (0.692-0.934) | P= .004 | 1.006 (0.999-1.013) | P= .07 | 0.935 (0.842-1.038) | P= .20 | 0.995 (0.987-1.004) | P= .31 |
| **Hospitalizations with ≥1 blood culture obtained (≥1-BC hospitalizations)** | | | | | | | | | | | |
| In-hospital mortality or hospice discharge | 0.042 (0.029-0.062) | 0.989 (0.976-1.002) | P= .10 | 1.555 (0.801-3.019) | P= .19 | 0.987 (0.958-1.016) | P= .37 | 1.544 (0.904-2.636) | P= .11 | 0.966 (0.926-1.007) | P= .10 |
| Total 30-day revisits | 0.412 (0.359-0.474) | 1.001 (0.996-1.006) | P= .66 | 0.849 (0.631-1.142) | P= .28 | 1.013 (0.992-1.034) | P= .22 | 0.727 (0.480-1.101) | P= .13 | 0.996 (0.969-1.023) | P= .77 |
| 30-day inpatient readmission | 0.217 (0.188-0.252) | 0.998 (0.993-1.004) | P= .50 | 0.925 (0.693-1.236) | P= .60 | 1.008 (0.992-1.024) | P= .33 | 1.014 (0.731-1.407) | P= .93 | 0.982 (0.960-1.005) | P= .13 |
| 30-day medical observation unit revisits | 0.049 (0.036-0.066) | 0.999 (0.989-1.009) | P= .85 | 2.003 (1.277-3.142) | P= .002 | 0.966 (0.942-0.991) | P= .009 | 0.970 (0.500-1.879) | P= .93 | 1.043 (0.997-1.090) | P= .06 |
| 30-day emergency department revisits | 0.148 (0.115-0.190) | 1.005 (0.997-1.014) | P= .22 | 0.556 (0.337-0.918) | P= .02 | 1.035 (1.004-1.067) | P= .03 | 0.439 (0.232-0.830) | P= .01 | 0.998 (0.957-1.041) | P= .92 |
| Length of stay, days | 7.687 (7.007-8.433) | 1.002 (0.999-1.005) | P= .17 | 1.112 (0.961-1.287) | P= .16 | 1.000 (0.991-1.009) | P= .97 | 0.942 (0.787-1.127) | P= .51 | 0.995 (0.983-1.008) | P= .46 |
| Total number of blood cultures obtained^c^ | 2.413 (2.296-2.535) | 1.002 (1.000-1.004) | P= .06 | 0.520 (0.452-0.598) | P< .001 | 1.002 (0.994-1.011) | P= .62 | 1.400 (1.221-1.606) | P< .001 | 1.006 (0.996-1.016) | P= .27 |
| Number of calendar days with ≥1 blood culture obtained^c^ | 1.382 (1.317-1.451) | 1.001 (0.999-1.003) | P= .28 | 0.914 (0.850-0.983) | P= .02 | 1.001 (0.996-1.005) | P= .78 | 1.009 (0.912-1.116) | P= .86 | 0.998 (0.992-1.005) | P= .62 |
| Time from admission to first blood culture obtained, days^c^ | 0.702 (0.569-0.866) | 0.997 (0.988-1.005) | P= .45 | 2.528 (1.503-4.254) | P< .001 | 0.974 (0.951-0.998) | P= .04 | 1.170 (0.836-1.638) | P= .36 | 1.014 (0.983-1.045) | P= .39 |
| ≥1 positive blood culture obtained^c^ | 0.167 (0.144-0.192) | 1.002 (0.997-1.008) | P= .41 | 0.818 (0.548-1.220) | P= .32 | 0.998 (0.976-1.020) | P= .87 | 1.121 (0.734-1.711) | P= .60 | 1.011 (0.979-1.044) | P= .50 |
| Total number of positive blood cultures obtained^c^ | 0.208 (0.177-0.245) | 1.003 (0.997-1.009) | P= .35 | 0.639 (0.442-0.923) | P= .02 | 1.001 (0.983-1.020) | P= .88 | 1.434 (0.981-2.096) | P= .06 | 1.004 (0.975-1.033) | P= .80 |
| Number of calendar days with ≥1 positive blood culture obtained^c^ | 0.164 (0.143-0.189) | 1.001 (0.996-1.007) | P= .61 | 0.841 (0.594-1.192) | P= .33 | 0.999 (0.980-1.018) | P= .89 | 1.195 (0.797-1.793) | P= .39 | 1.004 (0.975-1.033) | P= .81 |
| ≥1 antimicrobial administered | 4.537 (3.767-5.463) | 1.007 (1.000-1.013) | P= .04 | 1.239 (0.906-1.694) | P= .18 | 1.007 (0.986-1.028) | P= .52 | 0.626 (0.416-0.943) | P= .02 | 1.005 (0.979-1.031) | P= .71 |
| Time from admission to first antimicrobial administered, days | 0.846 (0.687-1.042) | 0.994 (0.988-1.001) | P= .07 | 1.391 (1.074-1.802) | P= .01 | 0.990 (0.978-1.001) | P= .09 | 1.456 (0.992-2.137) | P= .055 | 0.978 (0.951-1.006) | P= .12 |
| Days of therapy with ≥1 antimicrobial administered | 4.470 (4.064-4.916) | 1.004 (1.001-1.008) | P= .02 | 0.982 (0.860-1.122) | P= .79 | 1.005 (0.998-1.011) | P= .15 | 0.822 (0.705-0.958) | P= .01 | 1.000 (0.989-1.010) | P= .94 |
| ≥1 broad-spectrum antimicrobial administered | 1.647 (1.434-1.891) | 1.001 (0.996-1.006) | P= .64 | 1.437 (1.094-1.888) | P= .009 | 1.002 (0.989-1.016) | P= .73 | 0.671 (0.534-0.843) | P< .001 | 1.007 (0.988-1.027) | P= .45 |
| Time from admission to first broad-spectrum antimicrobial administered, days | 1.146 (0.938-1.401) | 1.001 (0.993-1.010) | P= .74 | 1.124 (0.771-1.638) | P= .54 | 0.988 (0.974-1.002) | P= .09 | 1.086 (0.835-1.413) | P= .54 | 1.013 (0.989-1.038) | P= .28 |
| Days of therapy with ≥1 broad-spectrum antimicrobial administered | 2.799 (2.522-3.105) | 1.002 (0.998-1.006) | P= .29 | 1.154 (0.975-1.366) | P= .09 | 1.003 (0.995-1.011) | P= .43 | 0.841 (0.706-1.002) | P= .053 | 0.998 (0.986-1.010) | P= .74 |
| ≥1 non-broad-spectrum antimicrobial administered | 1.528 (1.316-1.773) | 1.009 (1.003-1.014) | P= .002 | 0.751 (0.550-1.027) | P= .07 | 1.006 (0.989-1.024) | P= .48 | 0.951 (0.707-1.278) | P= .74 | 1.002 (0.981-1.024) | P= .83 |
| Time from admission to first non-broad-spectrum antimicrobial administered, days | 1.632 (1.282-2.078) | 0.994 (0.987-1.000) | P= .048 | 1.306 (0.998-1.709) | P= .052 | 0.994 (0.980-1.008) | P= .40 | 1.270 (0.874-1.847) | P= .21 | 0.983 (0.958-1.009) | P= .19 |
| Days of therapy with ≥1 non-broad-spectrum antimicrobial administered | 2.424 (2.101-2.796) | 1.007 (1.001-1.013) | P= .03 | 0.861 (0.665-1.115) | P= .26 | 1.006 (0.996-1.017) | P= .25 | 0.803 (0.669-0.965) | P= .02 | 1.003 (0.988-1.018) | P= .71 |

Abbreviations: OR, odds ratio; IRR, incidence rate ratio.

The pre-restriction period was June 26, 2023 to June 25, 2024; the restriction period was June 26, 2024 to December 23, 2024; and the post-restriction period was December 24, 2024 to June 25, 2025. The total 30-day readmission outcome is comprised of all readmissions, including inpatient readmissions, medical observation unit revisits, and emergency department revisits. Effects estimates for binary outcomes (in-hospital mortality or hospice discharge, ≥1 blood culture obtained, ≥1 positive blood culture, ≥1 antimicrobial administered, ≥1 broad-spectrum antimicrobial administered, and ≥1 non-broad-spectrum antimicrobial administered) were odds ratios. Effect estimates for count and continuous outcomes (30-day inpatient readmission outcomes, length of stay, total numbers of blood cultures and positive blood cultures, number of calendar days with ≥1 blood culture or ≥1 positive blood culture obtained, time from admission to first blood culture obtained, time from admission to first antimicrobial administered including broad and non-broad-spectrum, and days of therapy with ≥1 antimicrobial administered including broad and non-broad-spectrum), were incidence rate ratios.

^a^ Effect estimates for the overall culture positivity rate reflect the entirety of Pennsylvania Hospital rather than averaged measures for individual hospitalizations, and thus are not shown for the hospitalizations with ≥1 blood culture obtained subgroup (≥1-BC hospitalizations).

^b^ Effect estimates were not calculated for ≥1-BC hospitalizations as this subgroup had ≥1 blood culture obtained by definition.

^c^ Effect estimates were only calculated for ≥1-BC hospitalizations.

#### **eTable 13.** Effect Estimates of Blood Culture Restriction on Primary and Secondary Outcomes for Hospitalizations at All 3 Hospital Sites Combined with Washout Period of 1 Week.

|  | **(Intercept)** | **Slope before blood culture restriction** | | **Level change upon blood culture restriction** | | **Slope during blood culture restriction** | | **Level change at end of blood culture restriction** | | **Slope after blood culture restriction** | |
| --- | --- | --- | --- | --- | --- | --- | --- | --- | --- | --- | --- |
| **Outcome** |  | **OR or IRR** | **P value** | **OR or IRR** | **P value** | **OR or IRR** | **P value** | **OR or IRR** | **P value** | **OR or IRR** | **P value** |
| **All hospitalizations** | | | | | | | | | | | |
| In-hospital mortality or hospice discharge | 0.036 (0.032-0.040) | 0.987 (0.983-0.990) | P< .001 | 0.942 (0.787-1.127) | P= .51 | 1.009 (0.996-1.023) | P= .17 | 0.942 (0.737-1.205) | P= .64 | 0.971 (0.956-0.987) | P< .001 |
| Total 30-day revisits | 0.364 (0.348-0.381) | 0.999 (0.998-1.001) | P= .37 | 0.990 (0.919-1.067) | P= .80 | 0.999 (0.995-1.004) | P= .79 | 0.918 (0.840-1.004) | P= .06 | 1.012 (1.004-1.019) | P= .002 |
| 30-day inpatient readmission | 0.246 (0.231-0.262) | 0.998 (0.997-1.000) | P= .04 | 0.978 (0.884-1.082) | P= .67 | 0.999 (0.994-1.005) | P= .75 | 0.982 (0.880-1.096) | P= .75 | 1.013 (1.003-1.022) | P= .009 |
| 30-day medical observation unit revisits | 0.028 (0.026-0.031) | 1.001 (0.998-1.005) | P= .33 | 1.197 (1.063-1.349) | P= .003 | 0.989 (0.981-0.997) | P= .006 | 0.885 (0.717-1.093) | P= .26 | 1.017 (1.003-1.031) | P= .02 |
| 30-day emergency department revisits | 0.090 (0.083-0.098) | 1.002 (0.999-1.004) | P= .30 | 0.955 (0.840-1.085) | P= .48 | 1.004 (0.997-1.010) | P= .26 | 0.791 (0.689-0.907) | P< .001 | 1.007 (0.997-1.017) | P= .17 |
| Length of stay, days | 6.465 (6.309-6.624) | 1.000 (0.999-1.001) | P= .75 | 0.983 (0.945-1.022) | P= .38 | 1.003 (1.000-1.005) | P= .04 | 0.993 (0.941-1.047) | P= .79 | 0.993 (0.989-0.996) | P< .001 |
| Overall blood culture positivity^a^ | 0.048 (0.043-0.052) | 1.002 (0.999-1.006) | P= .15 | 1.298 (1.112-1.516) | P< .001 | 1.015 (1.005-1.025) | P= .004 | 0.713 (0.593-0.856) | P< .001 | 0.989 (0.977-1.002) | P= .09 |
| ≥1 blood culture obtained^b^ | 0.395 (0.377-0.413) | 0.999 (0.997-1.001) | P= .25 | 0.622 (0.562-0.688) | P< .001 | 1.010 (1.004-1.017) | P= .002 | 1.038 (0.929-1.160) | P= .51 | 0.988 (0.980-0.996) | P= .003 |
| ≥1 antimicrobial administered | 1.193 (1.142-1.246) | 1.001 (1.000-1.003) | P= .06 | 0.909 (0.860-0.961) | P< .001 | 1.003 (1.000-1.006) | P= .047 | 1.033 (0.954-1.119) | P= .43 | 0.994 (0.988-1.000) | P= .04 |
| Time from admission to first antimicrobial administered, days | 1.085 (1.041-1.132) | 1.000 (0.998-1.001) | P= .64 | 1.059 (0.983-1.141) | P= .13 | 1.001 (0.997-1.004) | P= .76 | 0.943 (0.878-1.014) | P= .11 | 1.001 (0.995-1.006) | P= .76 |
| Days of therapy with ≥1 antimicrobial administered | 3.089 (2.983-3.198) | 1.000 (0.999-1.001) | P= .83 | 0.958 (0.905-1.015) | P= .15 | 1.002 (0.998-1.006) | P= .23 | 1.021 (0.945-1.104) | P= .60 | 0.990 (0.985-0.995) | P< .001 |
| ≥1 broad-spectrum antimicrobial administered | 0.534 (0.516-0.553) | 1.000 (0.999-1.001) | P= .84 | 0.948 (0.902-0.998) | P= .04 | 1.003 (1.000-1.006) | P= .03 | 0.991 (0.928-1.057) | P= .78 | 0.994 (0.989-1.000) | P= .03 |
| Time from admission to first broad-spectrum antimicrobial administered, days | 1.538 (1.449-1.632) | 1.000 (0.998-1.002) | P= .93 | 1.060 (0.972-1.156) | P= .19 | 1.000 (0.997-1.003) | P= .99 | 0.997 (0.928-1.072) | P= .94 | 0.998 (0.992-1.004) | P= .46 |
| Days of therapy with ≥1 broad-spectrum antimicrobial administered | 2.001 (1.909-2.099) | 0.999 (0.998-1.001) | P= .36 | 0.967 (0.900-1.039) | P= .36 | 1.003 (0.998-1.008) | P= .32 | 1.002 (0.911-1.101) | P= .97 | 0.990 (0.983-0.996) | P= .002 |
| ≥1 non-broad-spectrum antimicrobial administered | 0.770 (0.736-0.807) | 1.001 (0.999-1.002) | P= .38 | 0.928 (0.873-0.987) | P= .02 | 1.002 (0.998-1.005) | P= .36 | 1.070 (1.000-1.144) | P= .048 | 0.992 (0.987-0.998) | P= .005 |
| Time from admission to first non-broad-spectrum antimicrobial administered, days | 1.720 (1.639-1.805) | 0.998 (0.997-1.000) | P= .07 | 1.078 (0.995-1.169) | P= .07 | 0.998 (0.994-1.003) | P= .48 | 1.009 (0.913-1.116) | P= .85 | 0.997 (0.990-1.004) | P= .46 |
| Days of therapy with ≥1 non-broad-spectrum antimicrobial administered | 1.739 (1.673-1.807) | 1.000 (0.999-1.002) | P= .58 | 0.937 (0.881-0.996) | P= .04 | 1.002 (0.998-1.007) | P= .34 | 1.046 (0.962-1.136) | P= .29 | 0.990 (0.984-0.996) | P< .001 |
| **Hospitalizations with ≥1 blood culture obtained (≥1-BC hospitalizations)** | | | | | | | | | | | |
| In-hospital mortality or hospice discharge | 0.083 (0.071-0.098) | 0.985 (0.980-0.990) | P< .001 | 1.340 (1.052-1.707) | P= .02 | 0.999 (0.985-1.014) | P= .89 | 0.982 (0.768-1.256) | P= .88 | 0.979 (0.961-0.997) | P= .02 |
| Total 30-day revisits | 0.358 (0.332-0.385) | 1.000 (0.998-1.003) | P= .72 | 0.967 (0.848-1.103) | P= .62 | 1.003 (0.996-1.011) | P= .38 | 0.844 (0.734-0.972) | P= .02 | 1.005 (0.994-1.015) | P= .38 |
| 30-day inpatient readmission | 0.233 (0.215-0.252) | 0.998 (0.996-1.001) | P= .20 | 1.020 (0.913-1.138) | P= .73 | 1.002 (0.994-1.010) | P= .63 | 0.954 (0.813-1.120) | P= .57 | 0.998 (0.987-1.009) | P= .71 |
| 30-day medical observation unit revisits | 0.030 (0.026-0.035) | 1.004 (0.999-1.009) | P= .15 | 1.120 (0.849-1.478) | P= .42 | 0.993 (0.978-1.009) | P= .37 | 0.661 (0.433-1.006) | P= .054 | 1.033 (1.005-1.062) | P= .02 |
| 30-day emergency department revisits | 0.096 (0.083-0.110) | 1.004 (0.999-1.009) | P= .13 | 0.823 (0.592-1.144) | P= .25 | 1.010 (0.990-1.030) | P= .32 | 0.695 (0.497-0.971) | P= .03 | 1.011 (0.985-1.038) | P= .43 |
| Length of stay, days | 11.973 (11.525-12.438) | 1.000 (0.998-1.001) | P= .47 | 1.148 (1.077-1.224) | P< .001 | 1.000 (0.996-1.003) | P= .86 | 1.016 (0.947-1.090) | P= .66 | 0.991 (0.986-0.996) | P< .001 |
| Total number of blood cultures obtained^c^ | 3.020 (2.951-3.091) | 1.001 (1.000-1.002) | P= .01 | 0.508 (0.460-0.562) | P< .001 | 1.009 (0.999-1.018) | P= .08 | 1.536 (1.326-1.779) | P< .001 | 0.991 (0.981-1.000) | P= .06 |
| Number of calendar days with ≥1 blood culture obtained^c^ | 1.705 (1.667-1.743) | 1.001 (1.000-1.001) | P= .06 | 0.931 (0.877-0.989) | P= .02 | 1.001 (0.997-1.005) | P= .67 | 1.064 (1.005-1.126) | P= .03 | 0.996 (0.991-1.000) | P= .06 |
| Time from admission to first blood culture obtained, days^c^ | 1.333 (1.226-1.450) | 0.998 (0.995-1.000) | P= .07 | 1.659 (1.422-1.936) | P< .001 | 0.998 (0.988-1.008) | P= .67 | 0.963 (0.810-1.146) | P= .67 | 0.989 (0.976-1.002) | P= .10 |
| ≥1 positive blood culture obtained^c^ | 0.121 (0.110-0.133) | 1.002 (0.999-1.005) | P= .27 | 0.744 (0.624-0.887) | P< .001 | 1.020 (1.009-1.031) | P< .001 | 1.008 (0.843-1.205) | P= .93 | 0.988 (0.975-1.000) | P= .050 |
| Total number of positive blood cultures obtained^c^ | 0.138 (0.125-0.152) | 1.003 (1.000-1.007) | P= .054 | 0.645 (0.550-0.755) | P< .001 | 1.023 (1.015-1.032) | P< .001 | 1.114 (0.946-1.312) | P= .20 | 0.980 (0.969-0.991) | P< .001 |
| Number of calendar days with ≥1 positive blood culture obtained^c^ | 0.123 (0.112-0.134) | 1.002 (0.999-1.005) | P= .17 | 0.733 (0.628-0.855) | P< .001 | 1.021 (1.012-1.029) | P< .001 | 1.098 (0.921-1.309) | P= .30 | 0.980 (0.969-0.991) | P< .001 |
| ≥1 antimicrobial administered | 6.586 (5.983-7.251) | 1.002 (0.999-1.005) | P= .24 | 1.276 (1.047-1.556) | P= .02 | 1.000 (0.990-1.010) | P= .95 | 0.891 (0.714-1.112) | P= .31 | 1.001 (0.984-1.019) | P= .87 |
| Time from admission to first antimicrobial administered, days | 6.586 (5.983-7.251) | 1.002 (0.999-1.005) | P= .24 | 1.276 (1.047-1.556) | P= .02 | 1.000 (0.990-1.010) | P= .95 | 0.891 (0.714-1.112) | P= .31 | 1.001 (0.984-1.019) | P= .87 |
| Days of therapy with ≥1 antimicrobial administered | 7.701 (7.375-8.041) | 1.000 (0.998-1.002) | P= .98 | 1.158 (1.064-1.261) | P< .001 | 0.998 (0.993-1.003) | P= .48 | 1.010 (0.914-1.117) | P= .84 | 0.994 (0.987-1.001) | P= .09 |
| ≥1 broad-spectrum antimicrobial administered | 7.701 (7.375-8.041) | 1.000 (0.998-1.002) | P= .98 | 1.158 (1.064-1.261) | P< .001 | 0.998 (0.993-1.003) | P= .48 | 1.010 (0.914-1.117) | P= .84 | 0.994 (0.987-1.001) | P= .09 |
| Time from admission to first broad-spectrum antimicrobial administered, days | 2.782 (2.554-3.030) | 1.000 (0.997-1.003) | P= .95 | 1.400 (1.164-1.684) | P< .001 | 0.996 (0.986-1.007) | P= .48 | 0.912 (0.793-1.049) | P= .20 | 1.004 (0.992-1.018) | P= .50 |
| Days of therapy with ≥1 broad-spectrum antimicrobial administered | 1.114 (1.053-1.178) | 0.999 (0.996-1.001) | P= .17 | 1.186 (1.047-1.344) | P= .007 | 0.999 (0.991-1.007) | P= .78 | 0.964 (0.820-1.134) | P= .66 | 0.996 (0.986-1.007) | P= .50 |
| ≥1 non-broad-spectrum antimicrobial administered | 5.552 (5.265-5.856) | 1.000 (0.998-1.001) | P= .74 | 1.204 (1.088-1.332) | P< .001 | 0.997 (0.991-1.003) | P= .33 | 1.002 (0.904-1.110) | P= .98 | 0.996 (0.988-1.003) | P= .26 |
| Time from admission to first non-broad-spectrum antimicrobial administered, days | 1.988 (1.870-2.114) | 0.999 (0.997-1.001) | P= .29 | 1.060 (0.955-1.176) | P= .27 | 1.001 (0.995-1.008) | P= .72 | 1.040 (0.902-1.198) | P= .59 | 0.990 (0.981-0.999) | P= .03 |
| Days of therapy with ≥1 non-broad-spectrum antimicrobial administered | 1.592 (1.489-1.702) | 1.000 (0.998-1.003) | P= .84 | 1.162 (1.009-1.338) | P= .04 | 0.996 (0.987-1.005) | P= .42 | 1.039 (0.886-1.220) | P= .64 | 1.000 (0.989-1.011) | P= .99 |

Abbreviations: OR, odds ratio; IRR, incidence rate ratio.

The pre-restriction period was June 26, 2023 to June 25, 2024; the restriction period was June 26, 2024 to December 23, 2024; and the post-restriction period was December 24, 2024 to June 25, 2025. The total 30-day readmission outcome is comprised of all readmissions, including inpatient readmissions, medical observation unit revisits, and emergency department revisits. Effects estimates for binary outcomes (in-hospital mortality or hospice discharge, ≥1 blood culture obtained, ≥1 positive blood culture, ≥1 antimicrobial administered, ≥1 broad-spectrum antimicrobial administered, and ≥1 non-broad-spectrum antimicrobial administered) were odds ratios. Effect estimates for count and continuous outcomes (30-day inpatient readmission outcomes, length of stay, total numbers of blood cultures and positive blood cultures, number of calendar days with ≥1 blood culture or ≥1 positive blood culture obtained, time from admission to first blood culture obtained, time from admission to first antimicrobial administered including broad and non-broad-spectrum, and days of therapy with ≥1 antimicrobial administered including broad and non-broad-spectrum), were incidence rate ratios.

^a^ Effect estimates for the overall culture positivity rate reflect all 3 hospital sites in the aggregate rather than averaged measures for individual hospitalizations, and thus are not shown for the hospitalizations with ≥1 blood culture obtained subgroup (≥1-BC hospitalizations).

^b^ Effect estimates were not calculated for ≥1-BC hospitalizations as this subgroup had ≥1 blood culture obtained by definition.

^c^ Effect estimates were only calculated for ≥1-BC hospitalizations.

#### **eTable 14.** Effect Estimates of Blood Culture Restriction on Primary and Secondary Outcomes for Hospitalizations at All 3 Hospital Sites Combined with Washout Period of 2 Weeks.

|  | **(Intercept)** | **Slope before blood culture restriction** | | **Level change upon blood culture restriction** | | **Slope during blood culture restriction** | | **Level change at end of blood culture restriction** | | **Slope after blood culture restriction** | |
| --- | --- | --- | --- | --- | --- | --- | --- | --- | --- | --- | --- |
| **Outcome** |  | **OR or IRR** | **P value** | **OR or IRR** | **P value** | **OR or IRR** | **P value** | **OR or IRR** | **P value** | **OR or IRR** | **P value** |
| **All hospitalizations** | | | | | | | | | | | |
| In-hospital mortality or hospice discharge | 0.036 (0.032-0.040) | 0.987 (0.983-0.990) | P< .001 | 0.966 (0.811-1.150) | P= .70 | 1.004 (0.993-1.015) | P= .50 | 1.014 (0.822-1.251) | P= .90 | 0.978 (0.963-0.993) | P= .004 |
| Total 30-day revisits | 0.364 (0.348-0.381) | 0.999 (0.998-1.001) | P= .37 | 0.991 (0.912-1.077) | P= .83 | 0.999 (0.994-1.005) | P= .83 | 0.929 (0.841-1.026) | P= .14 | 1.012 (1.003-1.020) | P= .009 |
| 30-day inpatient readmission | 0.246 (0.231-0.262) | 0.998 (0.997-1.000) | P= .050 | 0.971 (0.868-1.086) | P= .61 | 1.000 (0.993-1.007) | P= .90 | 0.995 (0.881-1.123) | P= .93 | 1.012 (1.001-1.023) | P= .04 |
| 30-day medical observation unit revisits | 0.028 (0.026-0.031) | 1.001 (0.998-1.004) | P= .44 | 1.205 (1.064-1.364) | P= .003 | 0.989 (0.981-0.998) | P= .02 | 0.881 (0.704-1.103) | P= .27 | 1.015 (1.000-1.031) | P= .06 |
| 30-day emergency department revisits | 0.090 (0.083-0.098) | 1.001 (0.998-1.005) | P= .34 | 0.971 (0.845-1.116) | P= .68 | 1.002 (0.995-1.010) | P= .54 | 0.798 (0.686-0.927) | P= .003 | 1.009 (0.998-1.021) | P= .12 |
| Length of stay, days | 6.472 (6.313-6.636) | 1.000 (0.999-1.001) | P= .88 | 0.981 (0.940-1.024) | P= .38 | 1.004 (1.001-1.006) | P= .01 | 0.968 (0.920-1.019) | P= .22 | 0.992 (0.988-0.996) | P< .001 |
| Overall blood culture positivity^a^ | 0.048 (0.044-0.053) | 1.002 (0.999-1.005) | P= .30 | 1.323 (1.126-1.555) | P< .001 | 1.017 (1.005-1.029) | P= .007 | 0.710 (0.580-0.868) | P< .001 | 0.986 (0.972-1.001) | P= .07 |
| ≥1 blood culture obtained^b^ | 0.393 (0.376-0.411) | 0.999 (0.998-1.001) | P= .41 | 0.620 (0.557-0.691) | P< .001 | 1.011 (1.003-1.019) | P= .009 | 1.035 (0.914-1.173) | P= .58 | 0.988 (0.979-0.998) | P= .02 |
| ≥1 antimicrobial administered | 1.187 (1.137-1.239) | 1.002 (1.000-1.003) | P= .02 | 0.885 (0.841-0.931) | P< .001 | 1.005 (1.002-1.008) | P< .001 | 0.986 (0.919-1.058) | P= .69 | 0.994 (0.988-0.999) | P= .03 |
| Time from admission to first antimicrobial administered, days | 1.095 (1.053-1.138) | 0.999 (0.998-1.001) | P= .22 | 1.060 (0.986-1.138) | P= .11 | 1.002 (0.998-1.006) | P= .38 | 0.927 (0.857-1.003) | P= .06 | 1.000 (0.993-1.006) | P= .89 |
| Days of therapy with ≥1 antimicrobial administered | 3.087 (2.979-3.198) | 1.000 (0.999-1.001) | P= .89 | 0.949 (0.890-1.012) | P= .11 | 1.004 (0.999-1.008) | P= .12 | 0.980 (0.912-1.053) | P= .59 | 0.990 (0.985-0.995) | P< .001 |
| ≥1 broad-spectrum antimicrobial administered | 0.532 (0.514-0.551) | 1.000 (0.999-1.001) | P= .89 | 0.928 (0.886-0.973) | P= .002 | 1.004 (1.002-1.007) | P< .001 | 0.956 (0.903-1.012) | P= .12 | 0.995 (0.990-0.999) | P= .03 |
| Time from admission to first broad-spectrum antimicrobial administered, days | 1.548 (1.459-1.642) | 1.000 (0.997-1.002) | P= .67 | 1.061 (0.969-1.163) | P= .20 | 1.001 (0.997-1.005) | P= .68 | 0.985 (0.909-1.068) | P= .72 | 0.997 (0.990-1.003) | P= .33 |
| Days of therapy with ≥1 broad-spectrum antimicrobial administered | 2.003 (1.909-2.103) | 0.999 (0.998-1.001) | P= .35 | 0.961 (0.886-1.041) | P= .33 | 1.004 (0.998-1.010) | P= .22 | 0.963 (0.876-1.058) | P= .43 | 0.990 (0.983-0.997) | P= .004 |
| ≥1 non-broad-spectrum antimicrobial administered | 0.766 (0.732-0.802) | 1.001 (1.000-1.002) | P= .18 | 0.903 (0.849-0.960) | P= .001 | 1.004 (0.999-1.008) | P= .09 | 1.031 (0.965-1.102) | P= .37 | 0.992 (0.986-0.997) | P= .004 |
| Time from admission to first non-broad-spectrum antimicrobial administered, days | 1.732 (1.653-1.816) | 0.998 (0.996-1.000) | P= .02 | 1.088 (1.005-1.178) | P= .04 | 0.998 (0.993-1.003) | P= .38 | 1.008 (0.907-1.121) | P= .88 | 0.998 (0.990-1.006) | P= .64 |
| Days of therapy with ≥1 non-broad-spectrum antimicrobial administered | 1.733 (1.667-1.802) | 1.001 (0.999-1.002) | P= .42 | 0.918 (0.859-0.982) | P= .01 | 1.004 (0.999-1.009) | P= .09 | 0.997 (0.926-1.074) | P= .94 | 0.989 (0.984-0.995) | P< .001 |
| **Hospitalizations with ≥1 blood culture obtained (≥1-BC hospitalizations)** | | | | | | | | | | | |
| In-hospital mortality or hospice discharge | 0.084 (0.072-0.099) | 0.984 (0.979-0.989) | P< .001 | 1.388 (1.067-1.806) | P= .01 | 0.995 (0.978-1.012) | P= .58 | 1.002 (0.773-1.301) | P= .99 | 0.984 (0.964-1.005) | P= .14 |
| Total 30-day revisits | 0.360 (0.335-0.388) | 1.000 (0.997-1.003) | P= .97 | 0.970 (0.843-1.116) | P= .67 | 1.006 (0.997-1.014) | P= .20 | 0.831 (0.722-0.957) | P= .01 | 1.002 (0.990-1.013) | P= .78 |
| 30-day inpatient readmission | 0.235 (0.217-0.254) | 0.998 (0.995-1.000) | P= .09 | 1.004 (0.900-1.121) | P= .94 | 1.006 (0.997-1.014) | P= .19 | 0.940 (0.815-1.084) | P= .39 | 0.992 (0.981-1.003) | P= .16 |
| 30-day medical observation unit revisits | 0.030 (0.025-0.035) | 1.004 (0.998-1.009) | P= .16 | 1.160 (0.884-1.524) | P= .28 | 0.992 (0.975-1.008) | P= .32 | 0.667 (0.423-1.054) | P= .08 | 1.034 (1.003-1.067) | P= .03 |
| 30-day emergency department revisits | 0.097 (0.084-0.111) | 1.003 (0.998-1.009) | P= .20 | 0.846 (0.594-1.206) | P= .36 | 1.011 (0.987-1.035) | P= .39 | 0.673 (0.470-0.965) | P= .03 | 1.013 (0.983-1.043) | P= .40 |
| Length of stay, days | 11.994 (11.536-12.470) | 0.999 (0.998-1.001) | P= .41 | 1.146 (1.068-1.229) | P< .001 | 1.000 (0.996-1.005) | P= .91 | 0.980 (0.916-1.049) | P= .56 | 0.991 (0.986-0.997) | P= .002 |
| Total number of blood cultures obtained^c^ | 3.019 (2.948-3.091) | 1.001 (1.000-1.002) | P= .01 | 0.508 (0.456-0.567) | P< .001 | 1.010 (0.999-1.021) | P= .09 | 1.514 (1.293-1.773) | P< .001 | 0.990 (0.979-1.001) | P= .08 |
| Number of calendar days with ≥1 blood culture obtained^c^ | 1.707 (1.668-1.746) | 1.001 (1.000-1.001) | P= .10 | 0.929 (0.870-0.993) | P= .03 | 1.002 (0.997-1.006) | P= .49 | 1.041 (0.982-1.105) | P= .18 | 0.995 (0.990-1.001) | P= .08 |
| Time from admission to first blood culture obtained, days^c^ | 1.345 (1.237-1.462) | 0.997 (0.995-1.000) | P= .03 | 1.631 (1.404-1.894) | P< .001 | 0.999 (0.988-1.010) | P= .86 | 0.920 (0.774-1.094) | P= .35 | 0.990 (0.976-1.004) | P= .15 |
| ≥1 positive blood culture obtained^c^ | 0.123 (0.112-0.135) | 1.001 (0.998-1.004) | P= .48 | 0.756 (0.624-0.916) | P= .004 | 1.023 (1.010-1.037) | P< .001 | 1.005 (0.831-1.216) | P= .96 | 0.984 (0.969-0.998) | P= .03 |
| Total number of positive blood cultures obtained^c^ | 0.139 (0.127-0.153) | 1.003 (0.999-1.006) | P= .11 | 0.655 (0.554-0.775) | P< .001 | 1.026 (1.017-1.036) | P< .001 | 1.093 (0.920-1.298) | P= .31 | 0.977 (0.964-0.989) | P< .001 |
| Number of calendar days with ≥1 positive blood culture obtained^c^ | 0.124 (0.114-0.135) | 1.001 (0.999-1.004) | P= .31 | 0.741 (0.629-0.872) | P< .001 | 1.024 (1.014-1.034) | P< .001 | 1.058 (0.881-1.270) | P= .55 | 0.977 (0.965-0.990) | P< .001 |
| ≥1 antimicrobial administered | 6.555 (5.944-7.228) | 1.002 (0.999-1.006) | P= .21 | 1.204 (1.014-1.431) | P= .03 | 1.003 (0.994-1.011) | P= .57 | 0.831 (0.678-1.018) | P= .07 | 1.003 (0.987-1.019) | P= .76 |
| Time from admission to first antimicrobial administered, days | 1.121 (1.060-1.186) | 0.998 (0.996-1.000) | P= .09 | 1.189 (1.049-1.349) | P= .007 | 0.998 (0.989-1.007) | P= .71 | 0.972 (0.819-1.153) | P= .74 | 0.997 (0.986-1.009) | P= .62 |
| Days of therapy with ≥1 antimicrobial administered | 7.714 (7.380-8.063) | 1.000 (0.998-1.002) | P= .89 | 1.145 (1.045-1.255) | P= .004 | 1.000 (0.994-1.006) | P= .90 | 0.956 (0.874-1.046) | P= .33 | 0.994 (0.987-1.001) | P= .10 |
| ≥1 broad-spectrum antimicrobial administered | 2.779 (2.547-3.031) | 1.000 (0.997-1.003) | P= .91 | 1.342 (1.125-1.601) | P= .001 | 0.999 (0.988-1.010) | P= .83 | 0.865 (0.755-0.990) | P= .04 | 1.004 (0.991-1.017) | P= .53 |
| Time from admission to first broad-spectrum antimicrobial administered, days | 1.605 (1.501-1.715) | 1.000 (0.997-1.002) | P= .88 | 1.160 (1.000-1.346) | P= .050 | 0.996 (0.986-1.007) | P= .49 | 1.055 (0.889-1.251) | P= .54 | 0.999 (0.987-1.011) | P= .85 |
| Days of therapy with ≥1 broad-spectrum antimicrobial administered | 5.574 (5.283-5.881) | 0.999 (0.998-1.001) | P= .57 | 1.193 (1.071-1.328) | P= .001 | 0.998 (0.992-1.005) | P= .66 | 0.952 (0.866-1.048) | P= .32 | 0.995 (0.987-1.003) | P= .24 |
| ≥1 non-broad-spectrum antimicrobial administered | 1.979 (1.861-2.104) | 0.999 (0.997-1.001) | P= .44 | 1.022 (0.913-1.143) | P= .71 | 1.004 (0.996-1.012) | P= .29 | 1.000 (0.859-1.165) | P= 1.00 | 0.987 (0.977-0.998) | P= .02 |
| Time from admission to first non-broad-spectrum antimicrobial administered, days | 2.446 (2.294-2.609) | 0.997 (0.994-0.999) | P= .006 | 1.309 (1.123-1.527) | P< .001 | 0.991 (0.982-1.000) | P= .06 | 1.111 (0.951-1.299) | P= .19 | 0.998 (0.985-1.010) | P= .71 |
| Days of therapy with ≥1 non-broad-spectrum antimicrobial administered | 3.955 (3.726-4.198) | 1.000 (0.998-1.003) | P= .71 | 1.079 (0.961-1.211) | P= .20 | 1.001 (0.993-1.010) | P= .78 | 0.966 (0.852-1.095) | P= .59 | 0.992 (0.982-1.002) | P= .11 |

Abbreviations: OR, odds ratio; IRR, incidence rate ratio.

The pre-restriction period was June 26, 2023 to June 25, 2024; the restriction period was June 26, 2024 to December 23, 2024; and the post-restriction period was December 24, 2024 to June 25, 2025. The total 30-day readmission outcome is comprised of all readmissions, including inpatient readmissions, medical observation unit revisits, and emergency department revisits. Effects estimates for binary outcomes (in-hospital mortality or hospice discharge, ≥1 blood culture obtained, ≥1 positive blood culture, ≥1 antimicrobial administered, ≥1 broad-spectrum antimicrobial administered, and ≥1 non-broad-spectrum antimicrobial administered) were odds ratios. Effect estimates for count and continuous outcomes (30-day inpatient readmission outcomes, length of stay, total numbers of blood cultures and positive blood cultures, number of calendar days with ≥1 blood culture or ≥1 positive blood culture obtained, time from admission to first blood culture obtained, time from admission to first antimicrobial administered including broad and non-broad-spectrum, and days of therapy with ≥1 antimicrobial administered including broad and non-broad-spectrum), were incidence rate ratios.

^a^ Effect estimates for the overall culture positivity rate reflect all 3 hospital sites in the aggregate rather than averaged measures for individual hospitalizations, and thus are not shown for the hospitalizations with ≥1 blood culture obtained subgroup (≥1-BC hospitalizations).

^b^ Effect estimates were not calculated for ≥1-BC hospitalizations as this subgroup had ≥1 blood culture obtained by definition.

^c^ Effect estimates were only calculated for ≥1-BC hospitalizations.

#### **eTable 15.** Effect Estimates of Blood Culture Restriction on Primary and Secondary Outcomes for Hospitalizations at All 3 Hospital Sites Combined, Excluding Hospitalizations Overlapping Restriction Cutoff Dates.

|  | **(Intercept)** | **Slope before blood culture restriction** | | **Level change upon blood culture restriction** | | **Slope during blood culture restriction** | | **Level change at end of blood culture restriction** | | **Slope after blood culture restriction** | |
| --- | --- | --- | --- | --- | --- | --- | --- | --- | --- | --- | --- |
| **Outcome** |  | **OR or IRR** | **P value** | **OR or IRR** | **P value** | **OR or IRR** | **P value** | **OR or IRR** | **P value** | **OR or IRR** | **P value** |
| **All hospitalizations** | | | | | | | | | | | |
| In-hospital mortality or hospice discharge | 0.037 (0.033-0.041) | 0.985 (0.981-0.988) | P< .001 | 1.121 (0.925-1.358) | P= .24 | 1.000 (0.989-1.011) | P= .99 | 1.138 (0.905-1.430) | P= .27 | 0.976 (0.962-0.990) | P< .001 |
| Total 30-day revisits | 0.365 (0.349-0.382) | 0.999 (0.998-1.001) | P= .24 | 1.020 (0.943-1.104) | P= .62 | 0.998 (0.993-1.003) | P= .45 | 0.935 (0.846-1.033) | P= .19 | 1.012 (1.004-1.019) | P= .003 |
| 30-day inpatient readmission | 0.247 (0.232-0.262) | 0.998 (0.996-1.000) | P= .02 | 1.017 (0.926-1.117) | P= .72 | 0.998 (0.993-1.003) | P= .34 | 0.996 (0.890-1.116) | P= .95 | 1.013 (1.004-1.022) | P= .004 |
| 30-day medical observation unit revisits | 0.028 (0.026-0.031) | 1.001 (0.998-1.004) | P= .44 | 1.261 (1.111-1.431) | P< .001 | 0.986 (0.978-0.995) | P= .001 | 0.954 (0.763-1.193) | P= .68 | 1.017 (1.004-1.031) | P= .01 |
| 30-day emergency department revisits | 0.090 (0.083-0.098) | 1.001 (0.999-1.004) | P= .35 | 0.954 (0.839-1.085) | P= .47 | 1.004 (0.997-1.010) | P= .28 | 0.798 (0.689-0.924) | P= .003 | 1.006 (0.996-1.015) | P= .28 |
| Length of stay, days | 6.808 (6.513-7.117) | 0.997 (0.995-0.999) | P= .002 | 1.192 (1.086-1.309) | P< .001 | 0.991 (0.984-0.999) | P= .02 | 1.272 (1.104-1.465) | P< .001 | 1.000 (0.992-1.008) | P= .97 |
| Overall blood culture positivity^a^ | 0.048 (0.044-0.053) | 1.002 (0.999-1.005) | P= .29 | 1.424 (1.186-1.710) | P< .001 | 1.010 (0.998-1.023) | P= .11 | 0.736 (0.592-0.916) | P= .006 | 0.992 (0.978-1.007) | P= .29 |
| ≥1 blood culture obtained^b^ | 0.406 (0.383-0.430) | 0.997 (0.995-1.000) | P= .03 | 0.699 (0.618-0.791) | P< .001 | 1.001 (0.990-1.011) | P= .90 | 1.276 (1.051-1.549) | P= .01 | 0.994 (0.982-1.005) | P= .29 |
| ≥1 antimicrobial administered | 1.218 (1.155-1.284) | 1.000 (0.998-1.002) | P= .92 | 0.977 (0.905-1.054) | P= .55 | 0.999 (0.994-1.003) | P= .59 | 1.123 (1.015-1.241) | P= .02 | 0.997 (0.990-1.003) | P= .32 |
| Time from admission to first antimicrobial administered, days | 1.124 (1.079-1.172) | 0.997 (0.996-0.999) | P= .001 | 1.243 (1.141-1.353) | P< .001 | 0.990 (0.984-0.997) | P= .003 | 1.150 (1.015-1.302) | P= .03 | 1.008 (1.001-1.016) | P= .03 |
| Days of therapy with ≥1 antimicrobial administered | 3.301 (3.121-3.493) | 0.995 (0.993-0.998) | P< .001 | 1.203 (1.075-1.346) | P= .001 | 0.989 (0.980-0.997) | P= .01 | 1.393 (1.178-1.648) | P< .001 | 0.998 (0.989-1.008) | P= .74 |
| ≥1 broad-spectrum antimicrobial administered | 0.548 (0.523-0.573) | 0.998 (0.996-1.000) | P= .10 | 1.049 (0.965-1.139) | P= .26 | 0.996 (0.991-1.002) | P= .20 | 1.129 (1.011-1.261) | P= .03 | 0.999 (0.992-1.006) | P= .71 |
| Time from admission to first broad-spectrum antimicrobial administered, days | 1.614 (1.520-1.715) | 0.997 (0.994-0.999) | P= .007 | 1.283 (1.151-1.430) | P< .001 | 0.987 (0.981-0.994) | P< .001 | 1.303 (1.144-1.483) | P< .001 | 1.007 (0.999-1.015) | P= .10 |
| Days of therapy with ≥1 broad-spectrum antimicrobial administered | 2.157 (2.016-2.309) | 0.994 (0.991-0.997) | P< .001 | 1.253 (1.093-1.436) | P= .001 | 0.986 (0.975-0.998) | P= .02 | 1.446 (1.168-1.791) | P< .001 | 1.000 (0.988-1.012) | P= .98 |
| ≥1 non-broad-spectrum antimicrobial administered | 0.786 (0.745-0.829) | 0.999 (0.998-1.001) | P= .60 | 0.986 (0.918-1.059) | P= .70 | 0.998 (0.994-1.002) | P= .40 | 1.145 (1.054-1.245) | P= .001 | 0.995 (0.990-1.001) | P= .08 |
| Time from admission to first non-broad-spectrum antimicrobial administered, days | 1.809 (1.721-1.903) | 0.995 (0.993-0.997) | P< .001 | 1.322 (1.207-1.448) | P< .001 | 0.987 (0.980-0.993) | P< .001 | 1.294 (1.120-1.496) | P< .001 | 1.005 (0.997-1.014) | P= .22 |
| Days of therapy with ≥1 non-broad-spectrum antimicrobial administered | 1.849 (1.749-1.954) | 0.996 (0.994-0.999) | P= .002 | 1.162 (1.053-1.282) | P= .003 | 0.990 (0.983-0.997) | P= .004 | 1.371 (1.202-1.564) | P< .001 | 0.997 (0.989-1.005) | P= .49 |
| **Hospitalizations with ≥1 blood culture obtained (≥1-BC hospitalizations)** | | | | | | | | | | | |
| In-hospital mortality or hospice discharge | 0.086 (0.074-0.101) | 0.982 (0.977-0.988) | P< .001 | 1.598 (1.256-2.035) | P< .001 | 0.990 (0.976-1.003) | P= .14 | 1.216 (0.945-1.564) | P= .13 | 0.984 (0.967-1.001) | P= .07 |
| Total 30-day revisits | 0.358 (0.332-0.385) | 1.000 (0.998-1.003) | P= .72 | 0.897 (0.802-1.004) | P= .06 | 1.009 (1.004-1.014) | P< .001 | 0.811 (0.705-0.932) | P= .003 | 0.997 (0.988-1.006) | P= .50 |
| 30-day inpatient readmission | 0.234 (0.216-0.253) | 0.998 (0.996-1.000) | P= .10 | 1.002 (0.895-1.121) | P= .98 | 1.004 (0.997-1.011) | P= .22 | 0.948 (0.819-1.097) | P= .47 | 0.994 (0.984-1.004) | P= .23 |
| 30-day medical observation unit revisits | 0.030 (0.025-0.035) | 1.004 (0.998-1.009) | P= .16 | 1.038 (0.791-1.361) | P= .79 | 0.999 (0.984-1.013) | P= .84 | 0.616 (0.412-0.921) | P= .02 | 1.027 (1.002-1.052) | P= .04 |
| 30-day emergency department revisits | 0.095 (0.082-0.109) | 1.005 (1.000-1.010) | P= .08 | 0.677 (0.536-0.854) | P< .001 | 1.022 (1.010-1.035) | P< .001 | 0.632 (0.455-0.877) | P= .006 | 0.995 (0.973-1.016) | P= .62 |
| Length of stay, days | 12.865 (12.228-13.536) | 0.995 (0.992-0.997) | P< .001 | 1.482 (1.322-1.660) | P< .001 | 0.984 (0.974-0.993) | P< .001 | 1.450 (1.208-1.739) | P< .001 | 1.002 (0.992-1.012) | P= .72 |
| Total number of blood cultures obtained^c^ | 3.110 (3.020-3.203) | 0.999 (0.998-1.000) | P= .10 | 0.547 (0.475-0.630) | P< .001 | 1.000 (0.985-1.016) | P= .98 | 1.873 (1.428-2.456) | P< .001 | 0.997 (0.982-1.013) | P= .73 |
| Number of calendar days with ≥1 blood culture obtained^c^ | 1.769 (1.717-1.822) | 0.998 (0.997-0.999) | P= .004 | 1.049 (0.977-1.126) | P= .18 | 0.993 (0.987-0.998) | P= .007 | 1.281 (1.166-1.407) | P< .001 | 1.001 (0.996-1.007) | P= .61 |
| Time from admission to first blood culture obtained, days^c^ | 1.409 (1.288-1.540) | 0.993 (0.990-0.997) | P< .001 | 2.271 (1.965-2.624) | P< .001 | 0.978 (0.968-0.987) | P< .001 | 1.440 (1.187-1.748) | P< .001 | 1.005 (0.993-1.017) | P= .43 |
| ≥1 positive blood culture obtained^c^ | 0.125 (0.114-0.138) | 1.000 (0.997-1.003) | P= .78 | 0.865 (0.713-1.049) | P= .14 | 1.009 (0.995-1.023) | P= .20 | 1.222 (0.963-1.551) | P= .10 | 0.995 (0.981-1.010) | P= .52 |
| Total number of positive blood cultures obtained^c^ | 0.143 (0.130-0.158) | 1.001 (0.997-1.004) | P= .76 | 0.759 (0.613-0.940) | P= .01 | 1.010 (0.995-1.025) | P= .18 | 1.403 (1.092-1.802) | P= .008 | 0.990 (0.974-1.006) | P= .22 |
| Number of calendar days with ≥1 positive blood culture obtained^c^ | 0.128 (0.117-0.140) | 0.999 (0.996-1.002) | P= .59 | 0.864 (0.720-1.037) | P= .12 | 1.007 (0.994-1.021) | P= .27 | 1.408 (1.102-1.799) | P= .006 | 0.989 (0.975-1.004) | P= .16 |
| ≥1 antimicrobial administered | 6.764 (6.119-7.478) | 1.000 (0.997-1.004) | P= .84 | 1.415 (1.169-1.714) | P< .001 | 0.994 (0.985-1.004) | P= .23 | 0.996 (0.796-1.246) | P= .97 | 1.004 (0.989-1.020) | P= .61 |
| Time from admission to first antimicrobial administered, days | 1.163 (1.097-1.233) | 0.995 (0.993-0.998) | P< .001 | 1.472 (1.297-1.670) | P< .001 | 0.982 (0.974-0.990) | P< .001 | 1.377 (1.168-1.624) | P< .001 | 1.010 (1.000-1.019) | P= .051 |
| Days of therapy with ≥1 antimicrobial administered | 8.302 (7.880-8.748) | 0.995 (0.993-0.997) | P< .001 | 1.472 (1.321-1.642) | P< .001 | 0.984 (0.975-0.993) | P< .001 | 1.407 (1.184-1.671) | P< .001 | 1.003 (0.993-1.013) | P= .51 |
| ≥1 broad-spectrum antimicrobial administered | 2.854 (2.618-3.112) | 0.999 (0.996-1.001) | P= .31 | 1.574 (1.358-1.825) | P< .001 | 0.990 (0.982-0.998) | P= .02 | 1.023 (0.881-1.187) | P= .77 | 1.009 (0.998-1.020) | P= .10 |
| Time from admission to first broad-spectrum antimicrobial administered, days | 1.682 (1.572-1.800) | 0.997 (0.994-0.999) | P= .008 | 1.452 (1.255-1.680) | P< .001 | 0.979 (0.972-0.987) | P< .001 | 1.488 (1.275-1.735) | P< .001 | 1.014 (1.004-1.024) | P= .004 |
| Days of therapy with ≥1 broad-spectrum antimicrobial administered | 6.003 (5.647-6.382) | 0.994 (0.992-0.997) | P< .001 | 1.532 (1.349-1.740) | P< .001 | 0.983 (0.972-0.993) | P= .002 | 1.404 (1.148-1.718) | P< .001 | 1.005 (0.993-1.017) | P= .41 |
| ≥1 non-broad-spectrum antimicrobial administered | 2.032 (1.904-2.168) | 0.998 (0.996-1.000) | P= .03 | 1.153 (1.034-1.285) | P= .01 | 0.996 (0.989-1.003) | P= .23 | 1.166 (1.003-1.355) | P= .046 | 0.994 (0.986-1.003) | P= .21 |
| Time from admission to first non-broad-spectrum antimicrobial administered, days | 2.582 (2.421-2.753) | 0.993 (0.990-0.995) | P< .001 | 1.638 (1.442-1.861) | P< .001 | 0.979 (0.971-0.988) | P< .001 | 1.528 (1.273-1.833) | P< .001 | 1.005 (0.994-1.016) | P= .37 |
| Days of therapy with ≥1 non-broad-spectrum antimicrobial administered | 4.263 (4.014-4.526) | 0.995 (0.993-0.998) | P< .001 | 1.434 (1.298-1.583) | P< .001 | 0.984 (0.977-0.990) | P< .001 | 1.443 (1.256-1.659) | P< .001 | 1.003 (0.995-1.012) | P= .48 |

Abbreviations: OR, odds ratio; IRR, incidence rate ratio.

The pre-restriction period was June 26, 2023 to June 25, 2024; the restriction period was June 26, 2024 to December 23, 2024; and the post-restriction period was December 24, 2024 to June 25, 2025. The total 30-day readmission outcome is comprised of all readmissions, including inpatient readmissions, medical observation unit revisits, and emergency department revisits. Effects estimates for binary outcomes (in-hospital mortality or hospice discharge, ≥1 blood culture obtained, ≥1 positive blood culture, ≥1 antimicrobial administered, ≥1 broad-spectrum antimicrobial administered, and ≥1 non-broad-spectrum antimicrobial administered) were odds ratios. Effect estimates for count and continuous outcomes (30-day inpatient readmission outcomes, length of stay, total numbers of blood cultures and positive blood cultures, number of calendar days with ≥1 blood culture or ≥1 positive blood culture obtained, time from admission to first blood culture obtained, time from admission to first antimicrobial administered including broad and non-broad-spectrum, and days of therapy with ≥1 antimicrobial administered including broad and non-broad-spectrum), were incidence rate ratios.

^a^ Effect estimates for the overall culture positivity rate reflect all 3 hospital sites in the aggregate rather than averaged measures for individual hospitalizations, and thus are not shown for the hospitalizations with ≥1 blood culture obtained subgroup (≥1-BC hospitalizations).

^b^ Effect estimates were not calculated for ≥1-BC hospitalizations as this subgroup had ≥1 blood culture obtained by definition.

^c^ Effect estimates were only calculated for ≥1-BC hospitalizations.
